# Updating the PRISMA reporting guideline for network meta-analysis: a scoping review

**DOI:** 10.1101/2025.02.06.25321746

**Authors:** Areti Angeliki Veroniki, Andrea C. Tricco, Daniella Rangira, Joanne E. McKenzie, Tianjing Li, Sharon E. Straus, Maureen Smith, Ferrán Catalá-López, Dianna Wolfe, Vera Nincic, Menelaos Konstantinidis, Juan Franco, David Tovey, Sai Surabi Thirugnanasampanthar, Jasmeen Dourka, Rachel Warren, George Wells, Adrienne Stevens, Brian Hutton

## Abstract

This scoping review represents the initial step in updating the Preferred Reporting Items for Systematic reviews and Meta-Analyses (PRISMA) extension for network meta-analysis (NMA) to improve the usability and relevance of systematic reviews with NMA for diverse audiences. The update will address gaps in reporting, such as documenting methods for assessing NMA homogeneity and transitivity, defining intervention nodes, and considering advances in statistical modeling. It will align with PRISMA 2020 and incorporate input from diverse knowledge users, including patients and the public. This scoping review included 61 studies, comprising 23 guidance documents and 38 overviews of reviews evaluating the completeness of reporting and methodological quality of published NMAs. We identified 37 additional NMA items for inclusion in the next step—the Delphi survey. Building on a 2014 scoping review by our team, we incorporated recent studies to inform the PRISMA-NMA update, adhering to established standards for guideline development.

**SUMMARY POINTS:**

1. Network meta-analyses (NMAs) are increasingly prominent in evidence synthesis, but their methodological transparency and reporting completeness remain inconsistent, which affects the reliability of results.
2. This scoping review highlights the need to update PRISMA-NMA to address evolving methodologies (e.g., component NMA) and reporting challenges to improve accessibility and utility of NMA findings.
3. The review underscores progress and gaps in NMA reporting since the 2015 PRISMA-NMA extension, focusing on synthesis methods, homogeneity, consistency and transitivity assessments, and network geometry.
4. Key recommendations, such as defining pre-specified nodes, specifying statistical methods, and addressing competing interests, were often overlooked in NMA reporting, but addressing these gaps could enhance transparency, reproducibility, and trust in NMA results.

## INTRODUCTION

Systematic reviews are fundamental to evidence-informed practice and policy, underscoring the need for their accurate, complete, and transparent reporting. The Preferred Reporting Items for Systematic reviews and Meta-Analyses (PRISMA) statement provides reporting^1^ recommendations to facilitate clarity, transparency, and reproducibility of systematic reviews. A number of extensions to PRISMA have been developed for particular review questions (e.g., diagnostic test accuracy), types of reviews (e.g., scoping reviews), or particular methods. An example of the latter is the PRISMA extension for network meta-analysis (NMA), published in 2015,^2^ which focuses on systematic reviews with NMAs. NMA allows the synthesis of evidence about the effect of multiple interventions in a single model, enabling comparisons of interventions that may plausibly be considered in specific contexts by incorporating indirect evidence alongside direct evidence. NMAs have revolutionized evidence synthesis and clinical decision-making across various fields^3–8^ over the past two decades, and have been adopted by agencies such as the National Institute for Health and Care Excellence (NICE), the Canada Drug Agency (CDA; formerly CADTH^9^), the National Health Service (NHS), and the World Health Organization (WHO) to inform health policies, cost-effectiveness analyses, and health technology assessments.^9–12^

The update of the 2009 PRISMA Statement in 2020,^1^ evidence of the need for improved reporting of systematic reviews with NMA and the continued evolution of NMA methodologies,^13–21^ provides the impetuses for updating PRISMA-NMA. The update of PRISMA-NMA aims to enhance the usability and relevance of systematic reviews that include NMAs for diverse audiences. Specifically, this update will address incompletely reported items (e.g., documentation of methods and findings for assessing homogeneity and similarity of intervention comparisons, and transparent synopsis of efforts to define intervention nodes), and incorporate advances in statistical modeling. Also, this update will provide an improved alignment with the updated PRISMA 2020 and involvement of diverse knowledge users— including patients and the public—throughout the update process to ensure new end-user perspectives are integrated.

As a first step in the process to update the PRISMA-NMA extension, we aimed to identify potential items for inclusion by undertaking a scoping review of studies evaluating the completeness of reporting and methodological quality of NMAs. We updated a 2014 scoping review performed by members of the research team^22^ to identify additional, more recent studies to inform the PRISMA-NMA extension update.^2^ The identified items will form the basis of the next step in the process – the Delphi survey (Figure 1). This process aligns with guidance for the development of reporting guidelines in health.^23^

**Figure 1.**
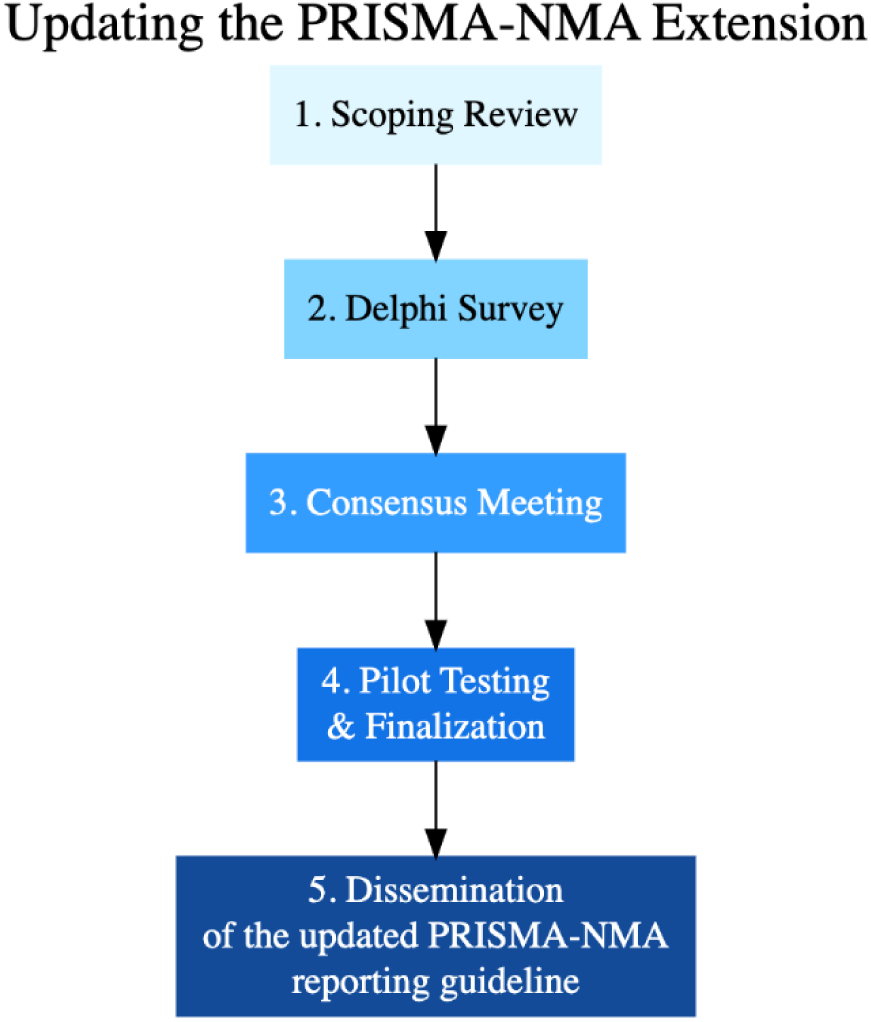
Flow diagram illustrating the individual steps involved in the updated PRISMA-NMA process.

## METHODS

### Protocol

The 2020 JBI (formerly Joanna Briggs Institute) methodological guidelines for scoping reviews were used to guide the methods of this scoping review^24^ and the PRISMA extension for scoping reviews was used to guide the reporting of this review.^25^ The methods for this study were drafted with input provided by evidence synthesis experts and knowledge users, including a patient partner (MS). We published our study protocol in the JBI^26^ and registered it with the Open Science Framework at https://osf.io/7bkwy. Any protocol deviations are mentioned in Appendix 1. A lay summary of this study, prepared by our patient partner (MS), is provided in Appendix 2.

### Literature search

We updated our previously developed literature search^22^ based on feedback from the steering committee composed of international researchers with at least ten years of experience in systematic reviews with NMA and experience in the development of reporting guidelines for studies in health (AAV, ACT, JEM, TL, SES, BH) and a patient partner with extensive experience in patient engagement in systematic reviews with NMA (MS). The literature search was developed by an experienced librarian and was peer-reviewed by another librarian using the Peer Review of Electronic Search Strategies (PRESS) Checklist (see Appendix 3 for literature search).^27^ Multiple databases were searched including MEDLINE, EMBASE, CINAHL, and *The Cochrane Library* from inception until December 11, 2023. We searched for unpublished literature based upon CDA Grey Matters guidance on December 13, 2023.^9^ We searched Google and a selection of organizational websites (Agency for Healthcare Research and Quality [AHRQ], Canadian Institute of Health Research [CIHR], CDA, EQUATOR, Guidelines International Network, IQWiG [Institute for Quality and Efficiency in Health Care] in Germany, JBI, NICE, and PRISMA). The literature search was supplemented by reviewing reference lists of the included studies, as well as by a key article^28^ identified by the steering committee.

Reference scanning was done using Citation Chaser.^29^ The search strategy was not limited by study design or language.

### Eligibility Criteria

We included any study design that offered reporting guidance or evaluated completeness of reporting pertaining to systematic reviews that include NMAs; studies assessing risk of bias and methodological quality relevant to NMA; and editorial guidelines or tutorials that described items related to reporting completeness of NMAs.^30^ Updates of studies were categorized as companion reports while the initially published paper was considered the main paper in the review. We excluded papers that were unrelated to NMA, did not focus on evaluating or providing reporting guidance, or were published before August 2013, as this was the last search date in the previous scoping review.^25^

### Study Selection

We used the systematic review software platform Synthesi.SR^31^ for screening. To ensure reliability, all reviewers completed a pilot test on 50 citations at level one (titles and abstracts) and 25 articles at level two (full-text papers) before screening, independently. After achieving high agreement at level one (88% in a single pilot test) and level two (84% after two pilot tests), team members (FCL, JD, MK, VN, RW, DW, AAV) independently screened all papers in duplicate (see Appendix 4 and 5 for screening guidance). Discrepancies were resolved by discussion with a third investigator (SST, AAV). We translated non-English articles using the DeepL Translate tool^32^ or with the help of team members. We summarised the search and selection process in a PRISMA 2020 flow diagram.^1^ The list of excluded study post-level two screening with reasoning is provided in Appendix 6.

### Data extraction

Data extraction was performed independently and in duplicate by team members (FCL, JD, MK, VN, RW, DW, AAV, and BH) using a standardized Excel form co-created by the steering committee and reviewers (see Appendix 7 for extraction guidance). We did not combine the results from the studies included in the previous scoping review,^22^ given that these already informed the development of the original PRISMA-NMA reporting guideline.^2^ We developed a new form, based on the previous review,^22^ with categories from which items were extracted: study characteristics (e.g., first author, year of publication, article source, study design [i.e. guidance document or overview of reviews]), study type (e.g., reporting or methodological quality in NMA), funding and funding source (e.g. public, industry, or mixed), reported objectives, inclusion criteria, number of NMAs included (in overviews of reviews [i.e., a study that assesses the reporting quality of a sample of NMAs]), timeframe of assessed literature, conflicts of interest, reporting/methodological guideline used, authors’ conclusions, and the authors’ recommendations as to which items should be reported. Prior to data extraction, we conducted a calibration exercise on a random sample of five included articles and modified the form according to feedback from the reviewers, steering committee, and our patient partner (MS). Data extraction began when we reached a high level of agreement (89% in a single pilot test). During this process our aim was to be overly inclusive and to capture as many details as possible, even if these were relevant only to the core PRISMA 2020.^1^ Results from in duplicate data extraction were combined and summarized for each paper by two team members (DR, SST) to comprehensively organize key findings. Then, two team members (DR, AAV) reviewed, combined identical items, and refined wording of the extracted items for clarity. The full list of items was organized according to the broader categories of PRISMA-NMA and PRISMA 2020 statement sections (i.e., Title, Abstract, Introduction, Methods, Results, Discussion, and Other information) and sub-categories (e.g., in the Methods category these were: Eligibility Criteria, Information Sources, Search Strategy, Selection Process, Data Collection, Data Items, Geometry of the Network, Study Risk of Bias Assessment, Effect Measures, Synthesis Methods, Assessment of Inconsistency, Reporting Bias Assessment, and Certainty Assessment). The final list of items was refined according to its relevance to the PRISMA-NMA or the core PRISMA 2020 reporting guideline, with additional input from the steering committee.

### Data synthesis and analysis

We present study characteristics and items suggested by authors across all the included studies (irrespective of type [i.e., overviews of reviews or guidance documents]) in tables. We present reporting items recommended as additional information to the PRISMA-NMA and/or PRISMA 2020 in tables. The frequency of items suggested in the overall sample of studies is depicted in bar-plots. We summarized the findings from overviews of reviews by providing details on the objectives, inclusion criteria, number of NMAs included in the overview, timeframe of the assessed literature, conflicts of interest, the reporting or methodological guidelines used for appraisal, and a summary of the authors’ conclusions. For guidance documents, we presented a summary of the objectives, conflicts of interest, and authors’ conclusions in tables.

## RESULTS

### Literature search

The literature search resulted in 2,815 citations identified through databases and registers, and 464 records identified through other methods including grey literature, reference scanning, and steering committee nomination (Figure 2). Prior to screening, 715 duplicates were removed, while 2,344 records were excluded at level one. At level two, we assessed 209 citations, of which 137 were excluded. In total, we included 72 papers: 62 primary studies and 10 companion reports (see Appendix 8 for final list of includes). Seven of the companion reports were extracted using the same process as for the primary studies to support the findings of their main papers.^33–39^ Of the remaining three companion reports, two were linked to the original PRISMA-NMA paper,^22, 40^ and one was a correction paper.^41^ The original PRISMA-NMA paper was included as a unique study; however, its data were not extracted or included in the final frequency counts as it is the study being updated in this review. Overall, 61 unique studies were extracted.

**Figure 2.**
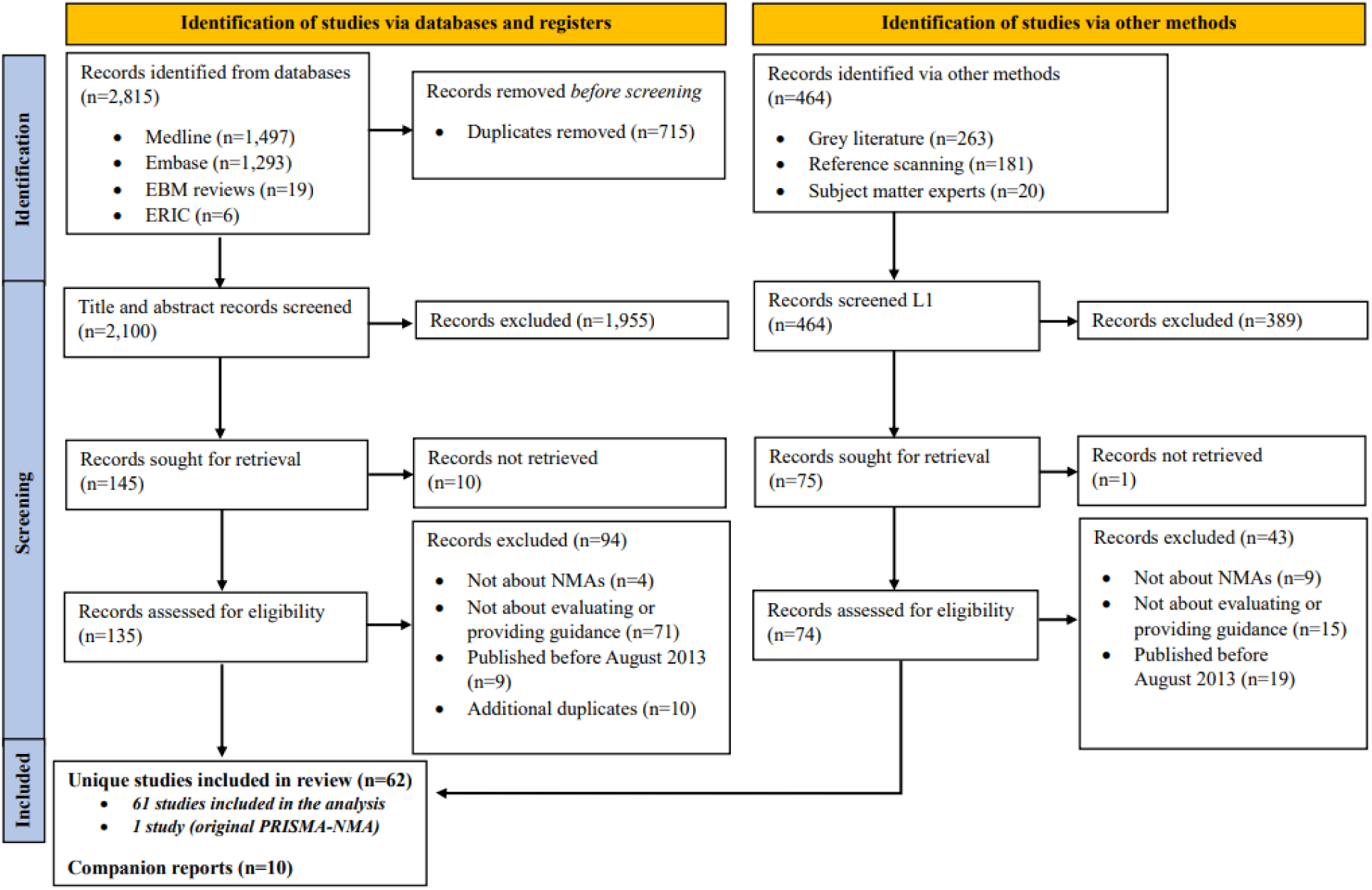
PRISMA Flow Diagram for study selection.

### Study characteristics

The study country of origin of the primary author, based on their affiliation, represented 15 countries (Table 1). The most commonly represented countries within the total 61 studies were: Canada (n = 14; 23.0%), followed by the UK (n=12; 19.7%), China (n=7; 11.6%), Switzerland (n=4; 6.6%), and Brazil (n=4; 6.6%). A breakdown of study types is shown in Table 1. Journals were the source of 51 (83.7%) studies, while the remaining studies were from grey literature sources including 2 (3.3%) textbooks, 6 (9.8%) reports, one (1.6%) presentation, and one (1.6%) thesis. Funding was reported in 34 (55.7%) of the 61 studies, and public funds were the most frequent source among the 34 studies reporting funding (n=27; 79.4%). Of the 61 included studies, 23 (37.7%) were guidance documents and 38 (62.3%) were overviews of reviews with NMA. Of the 23 guidance documents, 12 (52.2%) addressed reporting in NMA and 11 (47.8%) assessed methodological quality in NMA. Of the 38 overviews of reviews, 28 (73.7%) evaluated reporting completeness or quality in NMA and 10 (26.3%) evaluated quality or risk/source of bias in NMA methods.

**Table 1.** Characteristics of included studies (n=61)

| Characteristic | N (%) |
| --- | --- |
| *Country* |  |
| Australia | 1 (1.6%) |
| Brazil | 4 (6.6%) |
| Canada | 14 (23.0%) |
| China | 7 (11.6%) |
| France | 3 (4.9%) |
| Germany | 3 (4.9%) |
| Greece | 1 (1.6%) |
| Norway | 1 (1.6%) |
| Republic of Korea | 3 (4.9%) |
| South Africa | 1 (1.6%) |
| Spain | 2 (3.3%) |
| Switzerland | 4 (6.6%) |
| United Arab Emirates | 1 (1.6%) |
| United Kingdom | 12 (19.7%) |
| United States | 3 (4.9%) |
| Europe – Not Specified | 1 (1.6%) |
| *Source* |  |
| Journal | 51 (83.7%) |
| Grey lit - textbook | 2 (3.3%) |
| Grey lit - presentation | 1 (1.6%) |
| Grey lit - thesis | 1 (1.6%) |
| Grey lit - report | 6 (9.8%) |
| *Funding* |  |
| Yes | 34 (55.7%) |
| No | 14 (23.0%) |
| NR | 13 (21.3%) |
| *Source of Funding* |  |
| Publicly sponsored | 27 (44.2%) |
| Industry sponsored | 5 (8.2%) |
| Mixed | 2 (3.3%) |
| None | 14 (23.0%) |
| NR | 13 (21.3%) |
| *Study Type* |  |
| <u>Overviews of Reviews</u> |  |
| Evaluation of reporting completeness or quality in NMA | 28 (45.9%) |
| Evaluation of methods quality/risk of bias/source of bias in NMA | 10 (16.4%) |
| <u>Guidance Documents</u> |  |
| On reporting in NMA | 12 (19.7%) |
| On methodological quality in NMA | 11 (18.0%) |
| Total | 61 (100%) |

**Abbreviations** – NMA: Network Meta-analysis; NR: No Response

### Overview of reviews with NMA

The median number of NMAs included in each of the overviews was 62 (interquartile range [IQR: 25 to 200 NMAs]; Appendix 9: Table 2). The median timeframe assessed for NMAs in the literature was 51 years (IQR: 12.75 to 54.25 years). The longest timeframe spanned 58 years, from the assumed inception of databases in 1964 (based on MEDLINE^42^) to June 30, 2022 ^43^. In contrast, the shortest timeframe covered was 729 days, from January 1, 2018 to December 31, 2019.^44^ Conflicts of interest were reported in 31 (81.6%) overviews of reviews, with nine (23.7%) specifically disclosing conflicts of interest. Of the 38 overviews of reviews 16 (42.1%) used PRISMA-NMA, three (7.8%) PRISMA, two (5.2%) PRISMA-IPD^45^ [extension to individual participant data], nine (23.7%) A Measurement Tool to Assess Systematic Reviews (AMSTAR)^46^ (or AMSTAR 2 or R- AMSTAR), seven (18.4%) the International Society for Pharmacoeconomics and Outcomes Research (ISPOR)^47^ and seven (18.4%) national or international guidelines (e.g., CDA,^48^ NICE^11^). For example, Lee et *al*.^39^ revealed low-quality of conduct and reporting of NMAs published before 2017 with the most underreported items being network geometry, inconsistency assessment, and protocol registration. Gao et *al*.^49^ noted that the quality of NMAs published before 2018 was variable, including concerts regarding assessment of publication bias, evaluation of inconsistency, reporting of network geometry, and the structure of presentation. Similarly, Williams et *al*.^50^ highlighted that most NMAs published between 2008 and 2017 rarely assessed inconsistency or provided ranking plots and lacked presentation of effect estimates across all treatment comparisons with treatment rankings.

**Table 2.**
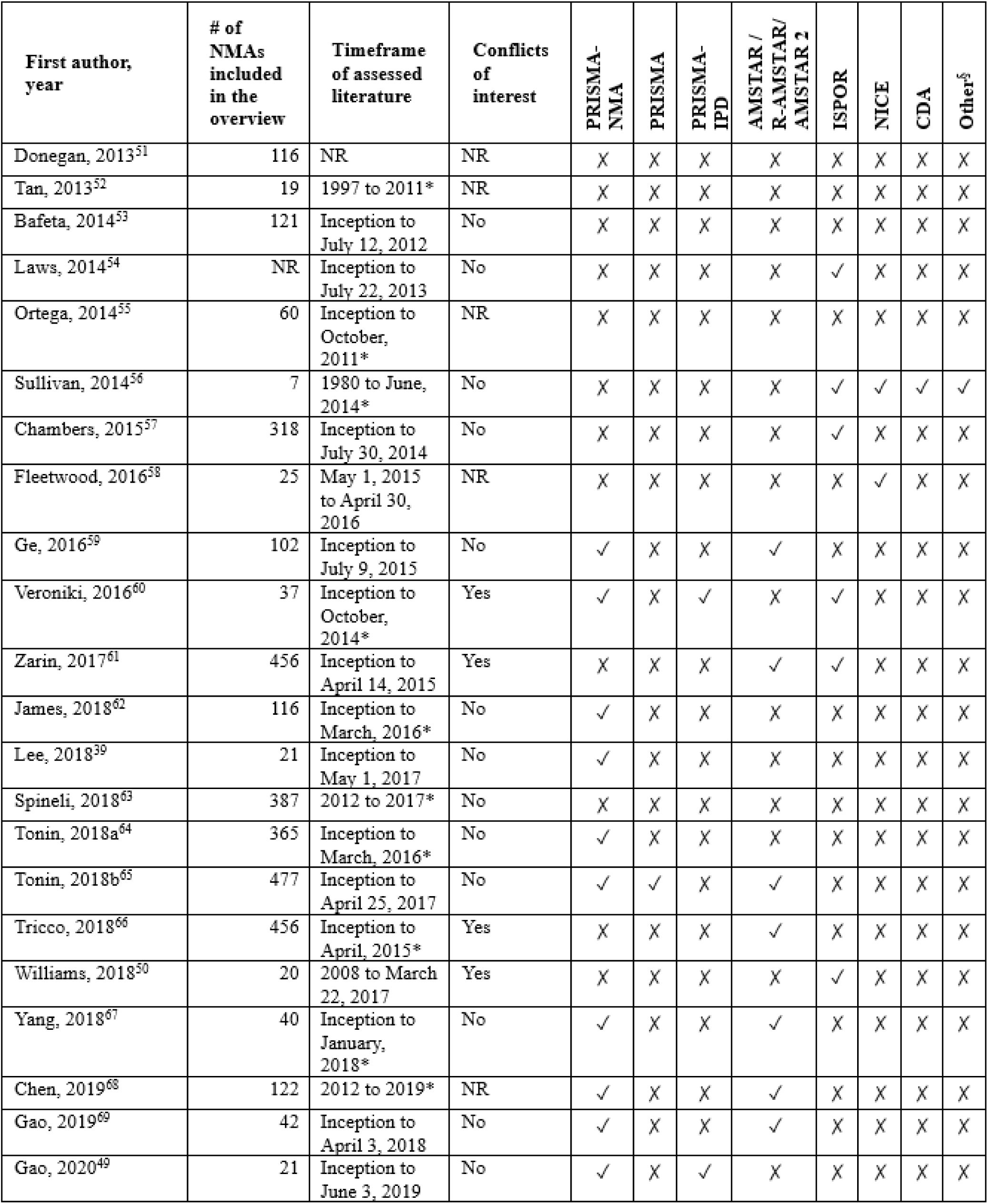
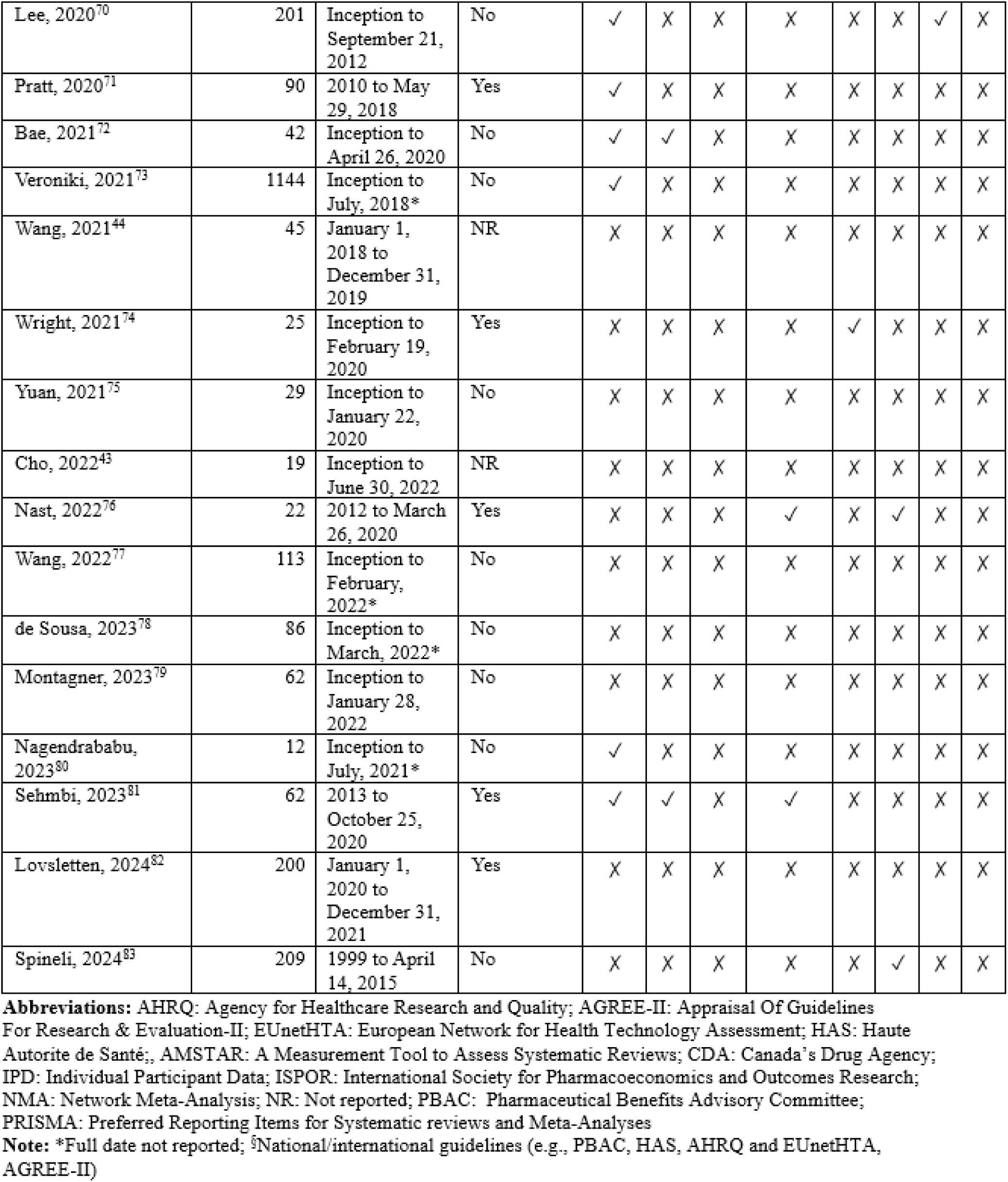
Characteristics of overviews of reviews with NMA (n=38)

### Guidance documents

The 23 guidance documents included in this scoping review aimed to provide tools and frameworks to enhance the reporting (n=12, 52.2%) and methodological quality (n=11, 47.8%) of NMAs (Appendix 10: Table 3). Chiocchia et *al*.^17^ presented the Risk of Bias due to Missing Evidence in Network Meta-Analysis (ROB-MEN) tool, which evaluates the risk of within-study and across-study bias due to missing evidence for each pairwise comparison in a NMA. Similarly, we identified studies presenting different approaches to evaluate credibility of NMA findings. Nikolakopoulou et *al*.^20^ presented the Confidence in Network Meta-Analysis (CINeMA) framework, which evaluates confidence in NMA results, Brignardello-Petersen et *al*.^21^ presented the Grading of Recommendations, Assessment, Development, and Evaluations (GRADE) approach extended to NMA to judge the certainty of NMA estimates, and Phillippo et *al*.^84^ presented the threshold analysis approach to assess confidence in NMA findings relevant to guideline recommendations and proposed methods to quantify the sensitivity of recommendations to plausible changes. Furthermore, the Risk of Bias in Network Meta-Analysis (RoB NMA) tool was introduced by Lunny et *al*.^85^ as a guidance to assess risk of bias in NMA and to identify potential limitations in NMA conduct that could lead to biased results and conclusions. Overall, conflicts of interest were reported in eight (34.8%) guidance documents, with three (13.0%) specifically disclosing conflicts of interest.

### PRISMA items

Across the 61 included studies, we identified 55 reporting items recommended as additional information to PRISMA-NMA or PRISMA 2020 (Appendix 11: Table 4). Among these 55 items, 37 were specific additional recommendations for PRISMA-NMA (Table 4). The frequencies of reporting for both the standard and additional guidance items beyond PRISMA-NMA or PRISMA 2020 in the 61 included studies are presented in Figure 3. The items reported by the fewest included studies were for the Availability of Data, Code, and Other Materials (n=4; 6.6%), Title (n = 14; 23.0%), Abstract (n = 15; 24.6%), and Certainty of evidence in results (n = 15; 24.6%) items, while the recommendations reported by most studies were for Synthesis methods (n = 48; 78.7%), Assessment of inconsistency (n = 45; 73.8%), Results of synthesis (n = 40; 65.6%), and Network geometry (n = 34; 55.7%). Among the 61 unique studies and seven companion reports that were extracted (n=68), additional items not specified in the PRISMA 2020 or the PRISMA-NMA checklists were recommended in 52 (76.5%) studies including defining pre-specified nodes and describing the node-making process,^14, 16, 21^ indicating whether statistical methods preserving within-study randomization were used,^50, 58, 64, 74, 86^ and in the presence of inconsistency, specifying whether both direct and indirect evidence were included in the NMA.^50, 58, 64, 74, 86^ Other frequent recommendations included describing the statistical model used to combine results, along with the associated algebra and analysis code,^52, 75, 77, 78^ detailing analysis methods to assess consistency and/or the transitivity assumption,^49, 54^ and reporting methods for evaluating the overall certainty of evidence (CINeMA, GRADE-NMA, GRADE for pairwise meta-analysis).^17, 20, 36, 44, 50, 59, 63, 87^ No additional reporting guidance was identified for the following PRISMA 2020 items: information sources, data collection process, conclusions, registration and protocol, support (funding), and competing interests. Similarly, no additional reporting guidance was identified for the presentation of network structure within the results category among the five NMA-related items.

**Figure 3.**
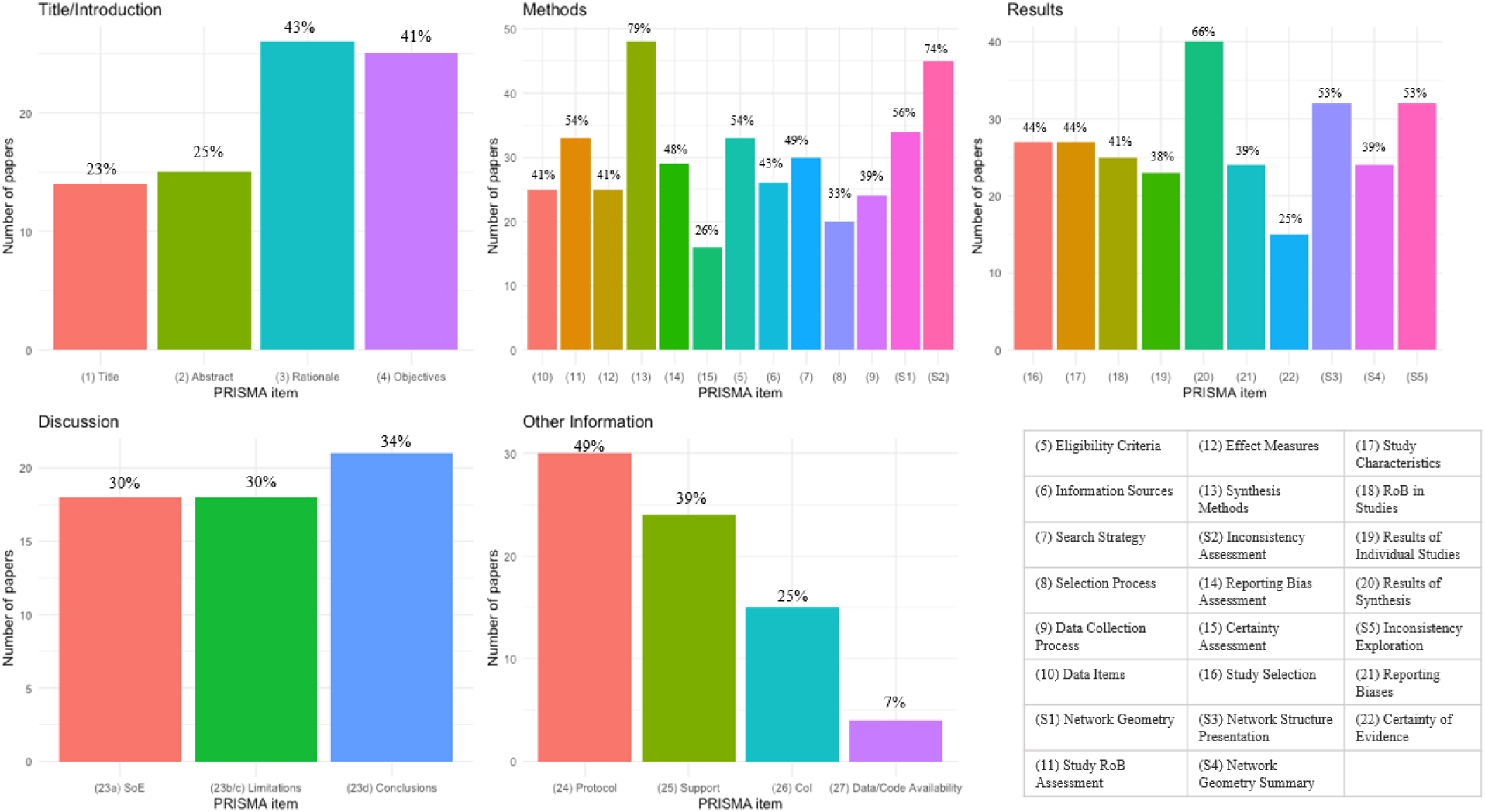
Frequencies of recommending each PRISMA item, considering both PRISMA-NMA and PRISMA 2020, in the included studies (n=61). **Abbreviations:** NMA: Network Meta-Analysis; PRISMA: Preferred Reporting Items for Systematic Reviews and Meta-Analyses; SoE: Summary of Evidence; CoI: Competing Interests; RoB: Risk of Bias.

**Table 4.**
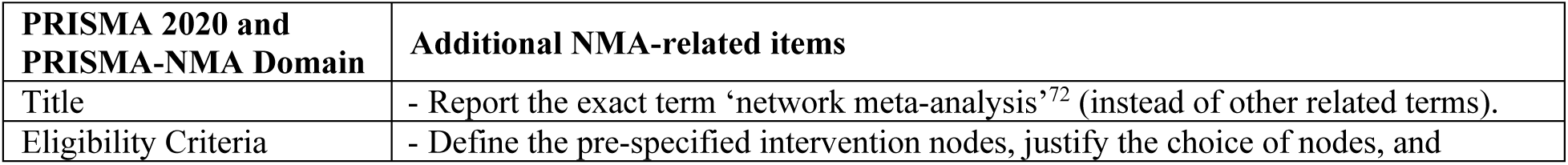
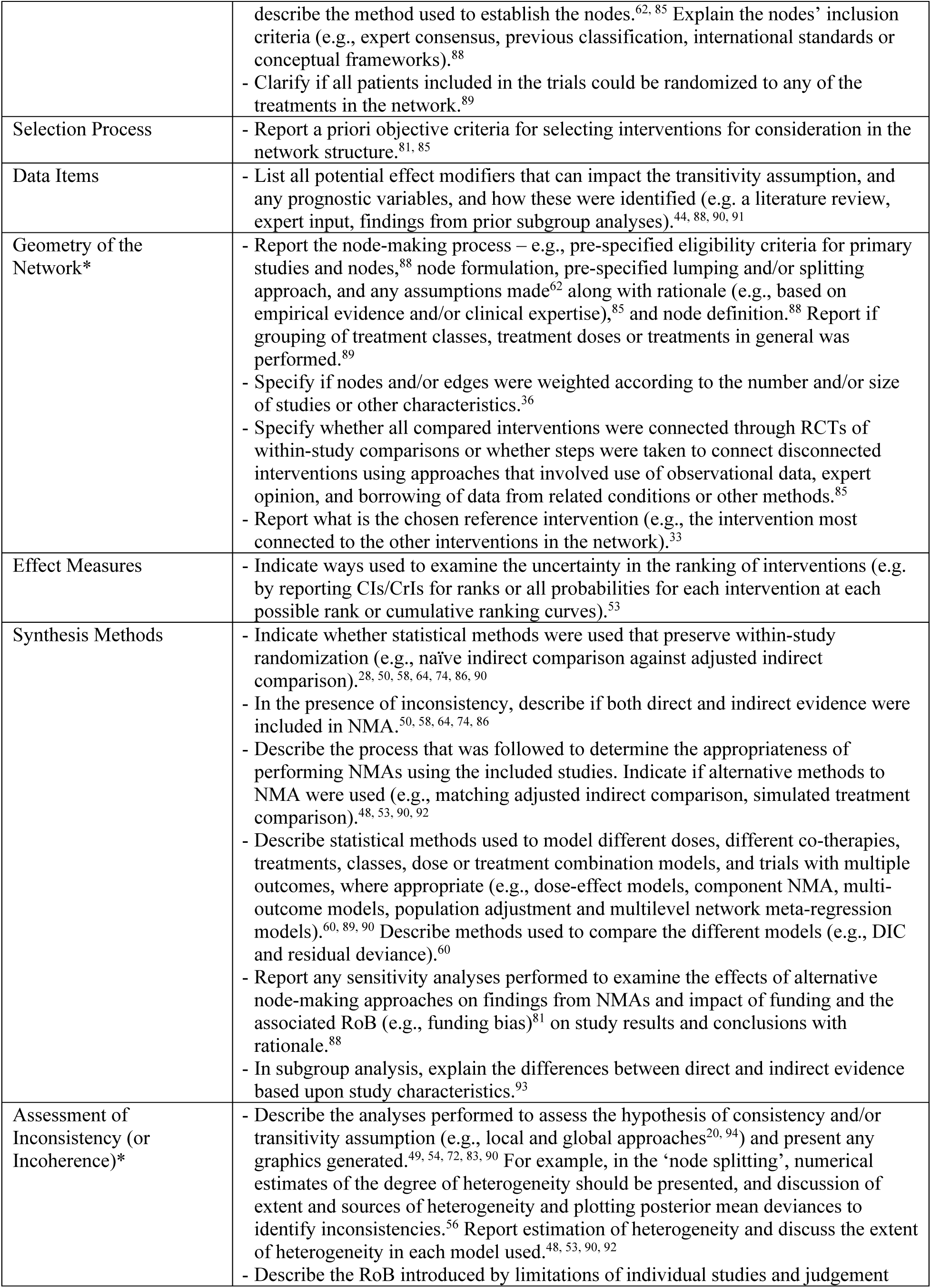
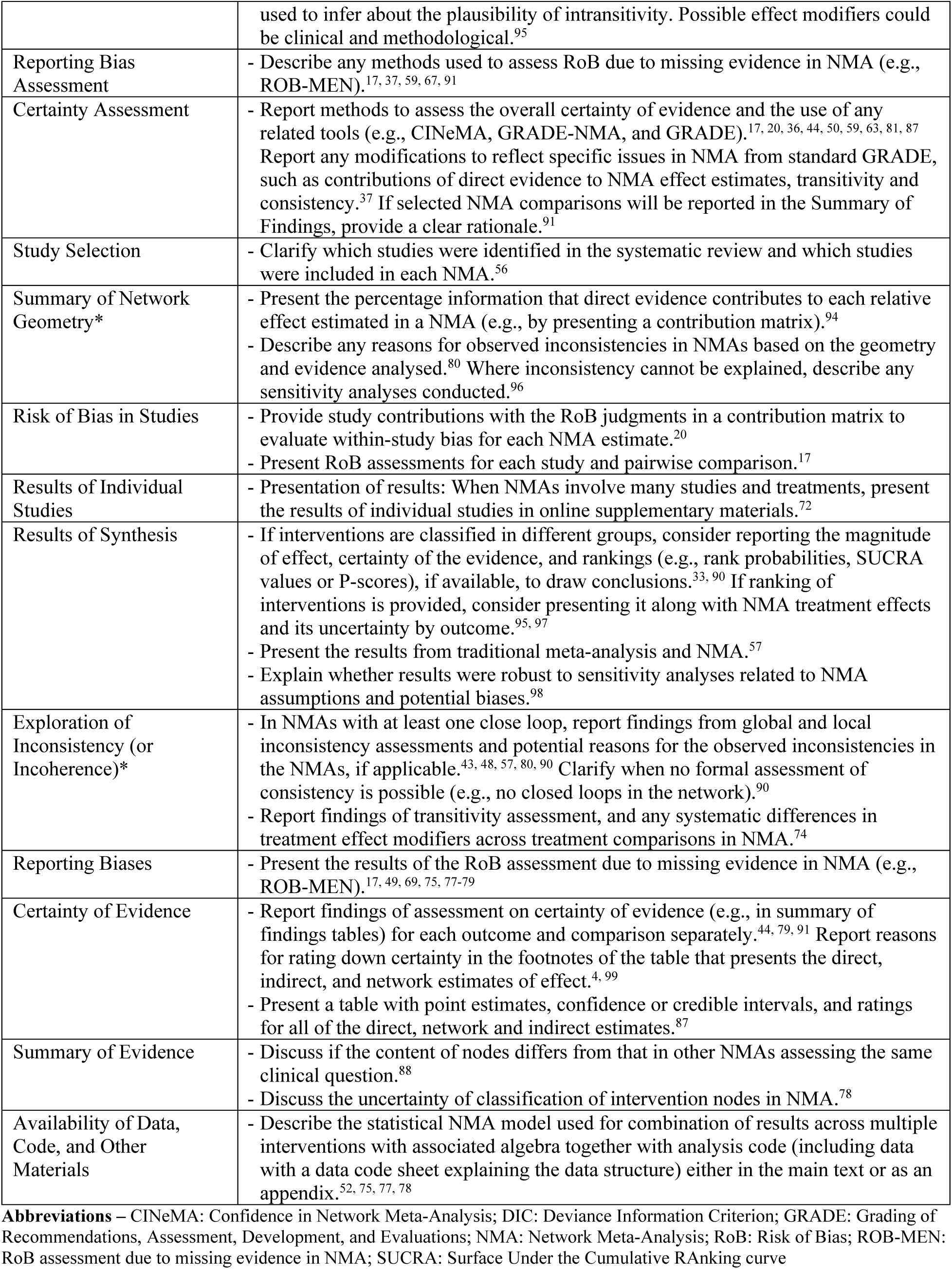
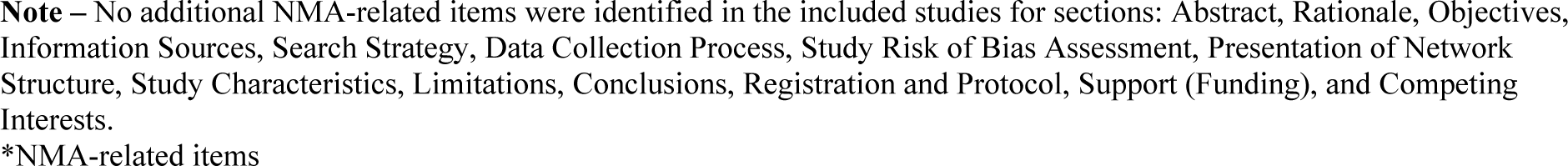
Additional items for PRISMA-NMA domains across included studies (n=61)

| PRISMA 2020 and PRISMA-NMA Domain | Additional NMA-related items |
| --- | --- |
| Title | - Report the exact term ‘network meta-analysis’<sup>72</sup> (instead of other related terms). |
| Eligibility Criteria | - Define the pre-specified intervention nodes, justify the choice of nodes, and |
|  | describe the method used to establish the nodes.<sup>62, 85</sup> Explain the nodes' inclusion criteria (e.g., expert consensus, previous classification, international standards or conceptual frameworks).<sup>88</sup><br>- Clarify if all patients included in the trials could be randomized to any of the treatments in the network.<sup>89</sup> |
| Selection Process | - Report a priori objective criteria for selecting interventions for consideration in the network structure.<sup>81, 85</sup> |
| Data Items | - List all potential effect modifiers that can impact the transitivity assumption, and any prognostic variables, and how these were identified (e.g. a literature review, expert input, findings from prior subgroup analyses).<sup>44, 88, 90, 91</sup> |
| Geometry of the Network* | - Report the node-making process – e.g., pre-specified eligibility criteria for primary studies and nodes,<sup>88</sup> node formulation, pre-specified lumping and/or splitting approach, and any assumptions made<sup>62</sup> along with rationale (e.g., based on empirical evidence and/or clinical expertise),<sup>85</sup> and node definition.<sup>88</sup> Report if grouping of treatment classes, treatment doses or treatments in general was performed.<sup>89</sup><br>- Specify if nodes and/or edges were weighted according to the number and/or size of studies or other characteristics.<sup>36</sup><br>- Specify whether all compared interventions were connected through RCTs of within-study comparisons or whether steps were taken to connect disconnected interventions using approaches that involved use of observational data, expert opinion, and borrowing of data from related conditions or other methods.<sup>85</sup><br>- Report what is the chosen reference intervention (e.g., the intervention most connected to the other interventions in the network).<sup>33</sup> |
| Effect Measures | - Indicate ways used to examine the uncertainty in the ranking of interventions (e.g. by reporting CIs/CrIs for ranks or all probabilities for each intervention at each possible rank or cumulative ranking curves).<sup>53</sup> |
| Synthesis Methods | - Indicate whether statistical methods were used that preserve within-study randomization (e.g., naïve indirect comparison against adjusted indirect comparison).<sup>28, 50, 58, 64, 74, 86, 90</sup><br>- In the presence of inconsistency, describe if both direct and indirect evidence were included in NMA.<sup>50, 58, 64, 74, 86</sup><br>- Describe the process that was followed to determine the appropriateness of performing NMAs using the included studies. Indicate if alternative methods to NMA were used (e.g., matching adjusted indirect comparison, simulated treatment comparison).<sup>48, 53, 90, 92</sup><br>- Describe statistical methods used to model different doses, different co-therapies, treatments, classes, dose or treatment combination models, and trials with multiple outcomes, where appropriate (e.g., dose-effect models, component NMA, multi-outcome models, population adjustment and multilevel network meta-regression models).<sup>60, 89, 90</sup> Describe methods used to compare the different models (e.g., DIC and residual deviance).<sup>60</sup><br>- Report any sensitivity analyses performed to examine the effects of alternative node-making approaches on findings from NMAs and impact of funding and the associated RoB (e.g., funding bias)<sup>81</sup> on study results and conclusions with rationale.<sup>88</sup><br>- In subgroup analysis, explain the differences between direct and indirect evidence based upon study characteristics.<sup>93</sup> |
| Assessment of Inconsistency (or Incoherence)* | - Describe the analyses performed to assess the hypothesis of consistency and/or transitivity assumption (e.g., local and global approaches<sup>20, 94</sup>) and present any graphics generated.<sup>49, 54, 72, 83, 90</sup> For example, in the 'node splitting', numerical estimates of the degree of heterogeneity should be presented, and discussion of extent and sources of heterogeneity and plotting posterior mean deviances to identify inconsistencies.<sup>56</sup> Report estimation of heterogeneity and discuss the extent of heterogeneity in each model used.<sup>48, 53, 90, 92</sup><br>- Describe the RoB introduced by limitations of individual studies and judgement |
|  | used to infer about the plausibility of intransitivity. Possible effect modifiers could be clinical and methodological.<sup>95</sup> |
| Reporting Bias Assessment | - Describe any methods used to assess RoB due to missing evidence in NMA (e.g., ROB-MEN).<sup>17, 37, 59, 67, 91</sup> |
| Certainty Assessment | - Report methods to assess the overall certainty of evidence and the use of any related tools (e.g., CINeMA, GRADE-NMA, and GRADE).<sup>17, 20, 36, 44, 50, 59, 63, 81, 87</sup><br>Report any modifications to reflect specific issues in NMA from standard GRADE, such as contributions of direct evidence to NMA effect estimates, transitivity and consistency.<sup>37</sup> If selected NMA comparisons will be reported in the Summary of Findings, provide a clear rationale.<sup>91</sup> |
| Study Selection | - Clarify which studies were identified in the systematic review and which studies were included in each NMA.<sup>56</sup> |
| Summary of Network Geometry* | - Present the percentage information that direct evidence contributes to each relative effect estimated in a NMA (e.g., by presenting a contribution matrix).<sup>94</sup><br>- Describe any reasons for observed inconsistencies in NMAs based on the geometry and evidence analysed.<sup>80</sup> Where inconsistency cannot be explained, describe any sensitivity analyses conducted.<sup>96</sup> |
| Risk of Bias in Studies | - Provide study contributions with the RoB judgments in a contribution matrix to evaluate within-study bias for each NMA estimate.<sup>20</sup><br>- Present RoB assessments for each study and pairwise comparison.<sup>17</sup> |
| Results of Individual Studies | - Presentation of results: When NMAs involve many studies and treatments, present the results of individual studies in online supplementary materials.<sup>72</sup> |
| Results of Synthesis | - If interventions are classified in different groups, consider reporting the magnitude of effect, certainty of the evidence, and rankings (e.g., rank probabilities, SUCRA values or P-scores), if available, to draw conclusions.<sup>33, 90</sup> If ranking of interventions is provided, consider presenting it along with NMA treatment effects and its uncertainty by outcome.<sup>95, 97</sup><br>- Present the results from traditional meta-analysis and NMA.<sup>57</sup><br>- Explain whether results were robust to sensitivity analyses related to NMA assumptions and potential biases.<sup>98</sup> |
| Exploration of Inconsistency (or Incoherence)* | - In NMAs with at least one close loop, report findings from global and local inconsistency assessments and potential reasons for the observed inconsistencies in the NMAs, if applicable.<sup>43, 48, 57, 80, 90</sup> Clarify when no formal assessment of consistency is possible (e.g., no closed loops in the network).<sup>90</sup><br>- Report findings of transitivity assessment, and any systematic differences in treatment effect modifiers across treatment comparisons in NMA.<sup>74</sup> |
| Reporting Biases | - Present the results of the RoB assessment due to missing evidence in NMA (e.g., ROB-MEN).<sup>17, 49, 69, 75, 77-79</sup> |
| Certainty of Evidence | - Report findings of assessment on certainty of evidence (e.g., in summary of findings tables) for each outcome and comparison separately.<sup>44, 79, 91</sup> Report reasons for rating down certainty in the footnotes of the table that presents the direct, indirect, and network estimates of effect.<sup>4, 99</sup><br>- Present a table with point estimates, confidence or credible intervals, and ratings for all of the direct, network and indirect estimates.<sup>87</sup> |
| Summary of Evidence | - Discuss if the content of nodes differs from that in other NMAs assessing the same clinical question.<sup>88</sup><br>- Discuss the uncertainty of classification of intervention nodes in NMA.<sup>78</sup> |
| Availability of Data, Code, and Other Materials | - Describe the statistical NMA model used for combination of results across multiple interventions with associated algebra together with analysis code (including data with a data code sheet explaining the data structure) either in the main text or as an appendix.<sup>52, 75, 77, 78</sup> |

**Abbreviations** – CINeMA: Confidence in Network Meta-Analysis; DIC: Deviance Information Criterion; GRADE: Grading of Recommendations, Assessment, Development, and Evaluations; NMA: Network Meta-Analysis; RoB: Risk of Bias; ROB-MEN: RoB assessment due to missing evidence in NMA; SUCRA: Surface Under the Cumulative Ranking curve
**Note** – No additional NMA-related items were identified in the included studies for sections: Abstract, Rationale, Objectives, Information Sources, Search Strategy, Data Collection Process, Study Risk of Bias Assessment, Presentation of Network Structure, Study Characteristics, Limitations, Conclusions, Registration and Protocol, Support (Funding), and Competing Interests. \*NMA-related items

## DISCUSSION

This scoping review highlights the progress and gaps in the reporting quality of NMAs since the introduction of the PRISMA-NMA extension in 2015.^2^ While NMAs have gained prominence in evidence synthesis, their methodological transparency and reporting completeness remain variable. Our findings show that the reporting of synthesis methods, methods to assess consistency and/or transitivity and their results, as well as of network geometry have received the most attention in overview of reviews with NMAs and NMA reporting guidance documents.

This likely reflects their critical importance to the reliability of NMA results. However, reporting of essential elements such as the title, abstract, and certainty of evidence were frequently overlooked in reporting which may reduce the accessibility and practical utility of NMA findings.

The findings from this review demonstrates a clear need to update PRISMA-NMA to address evolving methodologies and reporting challenges. Although this review primarily focuses on literature addressing reporting, rather than new methodological advancements, we recognize that evolving NMA methods (e.g., component NMA, individual participant NMA) may warrant attention in updated guidance. Recent approaches on evaluating certainty of evidence in NMA findings, such as the CINeMA framework,^20^ GRADE-NMA,^21^ and threshold analysis,^84^ as well as the RoB NMA tool to assess risk of bias in NMA,^17^ emphasize the growing complexity of NMAs and the demand for rigorous evaluation of confidence and methodological quality. However, the adoption of these approaches has not been universal, underscoring the need for additional reporting guidance to promote their integration into practice.

Importantly, this review identifies areas where PRISMA-NMA^2^ can be expanded to enhance its relevance, coverage, and usability. For example, defining pre-specified nodes,^36, 62, 85, 88^ specifying statistical methods that preserve within-study randomization,^50, 58, 64, 74, 86^ and describing methods used to explore the consistency and/or transitivity assumptions,^49, 54^ were frequently recommended in our included studies, but are not currently emphasized in PRISMA-NMA. Addressing these gaps in the updated checklist could improve the transparency and reproducibility of NMAs. Another significant finding is that the included articles placed little emphasis on reporting of competing interests (e.g., those related to the interventions assessed in the NMA). This omission could undermine trust in NMA findings. Strengthening the focus on this element in the updated PRISMA-NMA guideline could better align NMA practices with open science and transparency. Additionally, the included studies highlighted the importance of protocol transparency and data sharing,^44, 92^ aligning with the PRISMA 2020^1^ and EQUATOR^100^ guidance, as well as recent calls for open science promoting for reproducible research.

The strengths of our scoping review lie in its systematic approach and extensive engagement with knowledge users. By employing an integrated knowledge translation framework, we incorporated valuable insights throughout the research process, from refining the research question to interpreting the findings. A diverse range of knowledge users, including patient partners, journal editors, healthcare providers, policymakers, methodologists and statisticians, will be actively involved in all steps of updating the PRISMA-NMA guideline. This inclusive approach offers an opportunity to ensure the reporting guideline meets the needs of various end-users of evidence. Our review adhered to the 2020 JBI methodological guidelines for scoping reviews,^24^ with reviewers working in pairs and independently for screening and data extraction. We included both published and unpublished studies in any language, and the results were reported following the PRISMA extension for scoping reviews.^25^

Some limitations of this scoping review are worth noting. First, despite our systematic efforts to identify all relevant studies, it is possible that some studies were missed. However, our search strategy was developed by an experienced librarian, peer-reviewed by a second librarian, and all citations were screened in duplicate by our team. Also, we leveraged the expertise and experience of our authorship team to identify any further studies for inclusion. Second, we did not conduct a risk of bias assessment for the included studies. This is consistent with guidance for scoping reviews, which typically aim to map the available evidence rather than critically appraise individual sources of evidence.^24, 25^ Our primary objective was to compile a comprehensive list of potential checklist items to inform the update of the PRISMA-NMA reporting guideline, rather than to synthesize findings or evaluate the quality of the included evidence. Third, although we conducted the categorization of items in duplicate and had it reviewed by the steering committee, it is possible that some items could belong to a different category, which may affect the frequency calculations of each PRISMA item.

This study provides a basis for a Delphi panel survey and a subsequent online meeting of experts and knowledge users that will be held in 2025. The updated PRISMA-NMA should emphasize the principles of open science, transparency, and inclusivity by addressing existing reporting gaps and integrating perspectives from diverse knowledge users, including patients and the public. This approach can enhance trust, improve reproducibility, and strengthen the impact of NMAs supporting evidence-informed decision-making across healthcare and policy settings.

Lastly, this study may be of particular interest to methodologists, as it provides valuable insights into the process of updating a reporting guideline.

## CONCLUSIONS

This scoping review underscores the importance of updating the PRISMA-NMA reporting guideline to address gaps in reporting and align with advancements in NMA methodologies. Notable developments in areas such as synthesis methods, consistency and/or transitivity assessment, and critical aspects like certainty of evidence and transparency around competing interests have been overlooked in reporting. Expanding the PRISMA-NMA guideline to include additional guidance, such as reporting of node-making strategies and methods for assessing certainty of evidence, is essential for improving the usability and reliability of NMAs.

## Supplementary Files

**File name:** Supplementary Appendix Document.docx

**File format:** .docx

**Title of Data:** Appendices

**Description of Data:** Appendices for manuscript

**File name:** PRISMA-ScR Checklist.docx

**File format:** .docx

**Title of Data:** PRISMA-ScR checklist

**Description of Data:** PRISMA-ScR checklist for manuscript

## Data Sharing

The results and details of the studies included in this review can be found in the supplementary material.

## Supporting information

PRISMA Checklist

## Data Availability

All datasets supporting the conclusions of this article are included within the article.

## Acknowledgments

We thank Dr. Jessie McGowan for developing the literature searches. We thank Douglas M Salzwedel, MLIS (Information Specialist, Therapeutics Initiative, University of British Columbia) for peer reviewing the MEDLINE search strategy. We thank Neal Haddaway and Carole Lunny for providing feedback on our original grant proposal. We thank Yang Guo for translating and extraction the included Chinese-language study. We thank Brahmleen Kaur and Evan Mitton for their support in formatting the paper.

We acknowledge the traditional land of the Huron-Wendat, the Seneca, and most recently, the Mississaugas of the Credit River on which our research team operates in Toronto, Ontario, Canada. This land is still home to many Indigenous people from across Turtle Island and we are grateful to have the opportunity to work on this land.

## Dissemination

We plan to share findings in the following ways:

- Manuscript in a peer-reviewed journal
- Research brief
- National and international meetings/webinars
- Executive summary with infographics
- Websites (e.g., Knowledge Translation Program)
- Newsletters (e.g., Knowledge Translation Canada, Strategy for Patient-Oriented Research Evidence Alliance)
- Social media (e.g., BlueSky, LinkedIn)

## Ethics Approval

Not applicable.

## Patient and public engagement

Reporting completeness of a systematic review directly impacts patient care through evidence-based health and public health decision-making. Patients and the public can help promote reporting completeness and improve the use of systematic reviewers with network meta- analysis (NMA) in decision-making. Our team and steering committee involved a patient partner (MS) with expertise in patient engagement in reporting guidelines and systematic reviews with NMA, who provided their comments throughout the process of this scoping review. Our patient partner wrote a lay summary for this review (see Appendix 2).

## Transparency statement

The lead author (AAV) is the manuscript’s guarantor and affirms that it is an honest, accurate, and transparent account of the study being reported. No important aspects of the study have been omitted and any discrepancies from the study as originally planned have been explained.

## Open Access License

This is an open access article distributed in accordance with the Creative Commons Attribution Non Commercial (CC BY-4.0) license which permits others to distribute, remix, adapt, build upon this work non-commercially, and license their derivative works on different terms, provided the original work is properly cited and the use is non-commercial. See: http://creativecommons.org/licenses/by-nc/3.0/

## References

1. Page MJ, McKenzie JE, Bossuyt PM, Boutron I, Hoffmann TC, Mulrow CD, et al. The PRISMA 2020 statement: an updated guideline for reporting systematic reviews. BMJ. 2021;372:n71. doi: 10.1136/bmj.n71.

2. Hutton B, Salanti G, Caldwell DM, Chaimani A, Schmid CH, Cameron C, et al. The PRISMA extension statement for reporting of systematic reviews incorporating network meta-analyses of health care interventions: checklist and explanations. Ann Intern Med. 2015;162(11):777–84. doi: 10.7326/M14-2385.

3. Gnanavel S. Network meta-analysis in psychiatric research: Opportunities and caveats. Psychiatr Danub. 2018;30(3):367–9. doi: 10.24869/psyd.2018.367.

4. Al Khalifah R, Florez ID, Guyatt G, Thabane L. Network meta-analysis: users’ guide for pediatricians. BMC Pediatr. 2018;18(1):180. doi: 10.1186/s12887-018-1132-9.

5. Foote CJ, Chaudhry H, Bhandari M, Thabane L, Furukawa TA, Petrisor B, et al. Network Meta-analysis: Users’ Guide for Surgeons: Part I - Credibility. Clin Orthop Relat Res. 2015;473(7):2166–71. doi: 10.1007/s11999-015-4286-x.

6. Lee YH. Overview of Network Meta-analysis for a Rheumatologist. Journal of Rheumatic Diseases. 2016;23(1):1–15.

7. Watt J, Tricco AC, Straus S, Veroniki AA, Naglie G, Drucker AM. Research Techniques Made Simple: Network Meta-Analysis. J Invest Dermatol. 2019;139(1):4–12 e1. doi: 10.1016/j.jid.2018.10.028.

8. Phillips M, Chaudhary V. Understanding network meta-analysis methodology for the ophthalmologist. Curr Opin Ophthalmol. 2024;35(3):260–4. doi: 10.1097/ICU.0000000000001048.

9. Canadian Agency for Drugs and Technologies in Health. Grey Matters: A Practical Search Tool for Evidence-Based Medicine. Available from: [http://www.cadth.ca/resources/grey-matters].

10. National_Institutes_of_Health N. Systematic Review Standards & Organizations https://www.nihlibrary.nih.gov/services/systematic-review-service/systematic-review-standards-organizations.

11. NICE - National Institute for Health and Care Excellence. Available from: [https://www.nice.org.uk/ ].

12. WHO - World Health Organization. Available from: [https://www.who.int<u>/</u>].

13. Veroniki AA, Seitidis G, Nikolakopoulos S, Ballester M, Beltran J, Heijmans M, et al. Modeling Multicomponent Interventions in Network Meta-Analysis. Methods Mol Biol. 2022;2345:245–61. doi: 10.1007/978-1-0716-1566-9_15.

14. Rucker G, Petropoulou M, Schwarzer G. Network meta-analysis of multicomponent interventions. Biom J. 2020;62(3):808–21. doi: 10.1002/bimj.201800167.

15. Watt JA, Del Giovane C, Jackson D, Turner RM, Tricco AC, Mavridis D, et al. Incorporating dose effects in network meta-analysis. BMJ. 2022;376:e067003. doi: 10.1136/bmj-2021-067003.

16. Mavridis D, White IR. Dealing with missing outcome data in meta-analysis. Res Synth Methods. 2020;11(1):2–13. doi: 10.1002/jrsm.1349.

17. Chiocchia V, Nikolakopoulou A, Higgins JPT, Page MJ, Papakonstantinou T, Cipriani A, et al. ROB-MEN: a tool to assess risk of bias due to missing evidence in network meta-analysis. BMC Med. 2021;19(1):304. doi: 10.1186/s12916-021-02166-3.

18. Watt J, Del Giovane C. Network Meta-Analysis. Methods Mol Biol. 2022;2345:187-201. doi: 10.1007/978-1-0716-1566-9_12.

19. Brignardello-Petersen R, Tomlinson G, Florez I, Rind DM, Chu D, Morgan R, et al. Grading of recommendations assessment, development, and evaluation concept article 5: addressing intransitivity in a network meta-analysis. J Clin Epidemiol. 2023;160:151–9. doi: 10.1016/j.jclinepi.2023.06.010.

20. Nikolakopoulou A, Higgins JPT, Papakonstantinou T, Chaimani A, Del Giovane C, Egger M, et al. CINeMA: An approach for assessing confidence in the results of a network meta-analysis. PLoS Med. 2020;17(4):e1003082. doi: 10.1371/journal.pmed.1003082.

21. Brignardello-Petersen R, Florez ID, Izcovich A, Santesso N, Hazlewood G, Alhazanni W, et al. GRADE approach to drawing conclusions from a network meta-analysis using a minimally contextualised framework. BMJ. 2020;371:m3900. doi: 10.1136/bmj.m3900.

22. Hutton B, Salanti G, Chaimani A, Caldwell DM, Schmid C, Thorlund K, et al. The quality of reporting methods and results in network meta-analyses: an overview of reviews and suggestions for improvement. PLoS One. 2014;9(3):e92508. doi: 10.1371/journal.pone.0092508.

23. Moher D, Schulz KF, Simera I, Altman DG. Guidance for developers of health research reporting guidelines. PLoS Med. 2010;7(2):e1000217. doi: 10.1371/journal.pmed.1000217.

24. Peters MDJ, Marnie C, Tricco AC, Pollock D, Munn Z, Alexander L, et al. Updated methodological guidance for the conduct of scoping reviews. JBI Evid Synth. 2020;18(10):2119–26. doi: 10.11124/JBIES-20-00167.

25. Tricco AC, Lillie E, Zarin W, O’Brien KK, Colquhoun H, Levac D, et al. PRISMA Extension for Scoping Reviews (PRISMA-ScR): Checklist and Explanation. Ann Intern Med. 2018;169(7):467–73. doi: 10.7326/M18-0850.

26. Veroniki AA, Hutton B, Stevens A, McKenzie JE, Page MJ, Moher D, et al. Update to the PRISMA guidelines for network meta-analyses and scoping reviews and development of guidelines for rapid reviews: a scoping review protocol. JBI Evid Synth. 2025 doi: 10.11124/JBIES-24-00308.

27. PRESS - Peer Review of Electronic Search Strategies: 2015 Guideline Explanation and Elaboration (PRESS E&E). Ottawa: CADTH; 2016 Jan.

28. Lunny C, Veroniki AA, Hutton B, White I, Higgins J, Wright JM, et al. Knowledge user survey and Delphi process to inform development of a new risk of bias tool to assess systematic reviews with network meta-analysis (RoB NMA tool). BMJ Evid Based Med. 2022 doi: 10.1136/bmjebm-2022-111944.

29. Haddaway NR, Grainger MJ, Gray CT. Citationchaser: A tool for transparent and efficient forward and backward citation chasing in systematic searching. Res Synth Methods. 2022;13(4):533–45. doi: 10.1002/jrsm.1563.

30. Cope S, Zhang J, Saletan S, Smiechowski B, Jansen JP, Schmid P. A process for assessing the feasibility of a network meta-analysis: a case study of everolimus in combination with hormonal therapy versus chemotherapy for advanced breast cancer. BMC Med. 2014;12:93. doi: 10.1186/1741-7015-12-93.

31. Synthesi.SR Systematic Review Tool: St. Michael’s Hospital; 2006. The Joint Program in Knowledge Translation]. Available from: [http://knowledgetranslation.ca/sysrev/login.php].

32. DeepL SE. DeepL Translator 2017. Available from: [deepl.com/en/translator].

33. Brignardello-Petersen R, Izcovich A, Rochwerg B, Florez ID, Hazlewood G, Alhazanni W, et al. GRADE approach to drawing conclusions from a network meta-analysis using a partially contextualised framework. BMJ. 2020;371:m3907. doi: 10.1136/bmj.m3907.

34. Nagendrababu V, Faggion CM, Jr., Pulikkotil SJ, Alatta A, Dummer PMH. Methodological assessment and overall confidence in the results of systematic reviews with network meta-analyses in Endodontics. Int Endod J. 2022;55(5):393–404. doi: 10.1111/iej.13693.

35. Sehmbi H, Retter S, Shah UJ, Nguyen D, Martin J, Uppal V. Epidemiological, methodological, and statistical characteristics of network meta-analysis in anaesthesia: a systematic review. Br J Anaesth. 2023;130(3):272–86. doi: 10.1016/j.bja.2022.08.042.

36. Papakonstantinou T, Nikolakopoulou A, Higgins JPT, Egger M, Salanti G. CINeMA: Software for semiautomated assessment of the confidence in the results of network meta-analysis. Campbell Syst Rev. 2020;16(1):e1080. doi: 10.1002/cl2.1080.

37. Salanti G, Del Giovane C, Chaimani A, Caldwell DM, Higgins JP. Evaluating the quality of evidence from a network meta-analysis. PLoS One. 2014;9(7):e99682. doi: 10.1371/journal.pone.0099682.

38. Veroniki AA, Wong EKC, Lunny C, Martinez Molina JC, Florez ID, Tricco AC, et al. Does type of funding affect reporting in network meta-analysis? A scoping review of network meta-analyses. Syst Rev. 2023;12(1):81. doi: 10.1186/s13643-023-02235-z.

39. Lee DW, Shin IS. Critical quality evaluation of network meta-analyses in dental care. J Dent. 2018;75:7–11. doi: 10.1016/j.jdent.2018.05.010.

40. Hutton B, Catala-Lopez F, Moher D. [The PRISMA statement extension for systematic reviews incorporating network meta-analysis: PRISMA-NMA]. Med Clin (Barc). 2016;147(6):262–6. doi: 10.1016/j.medcli.2016.02.025.

41. Chambers JD, Naci H, Wouters OJ, Pyo J, Gunjal S, Kennedy IR, et al. Correction: An Assessment of the Methodological Quality of Published Network Meta-Analyses: A Systematic Review. PLoS One. 2015;10(7):e0131953. doi: 10.1371/journal.pone.0131953.

42. Medicine NLo. MEDLINE Overview 2024. Available from: [https://www.nlm.nih.gov/medline/medline_overview.html#:~:text=MEDLINE%20is%20the%20online%20counterpart,Medical%20Subject%20Headings%20(MeSH)].

43. Cho SH, Shin IS. Evaluation of the Reporting Standard Guidelines of Network Meta-Analyses in Physical Therapy: A Systematic Review. Healthcare (Basel). 2022;10(12) doi: 10.3390/healthcare10122371.

44. Wang R, Dwan K, Showell MG, van Wely M, Mol BW, Askie L, et al. Reporting of Cochrane systematic review protocols with network meta-analyses-A scoping review. Res Synth Methods. 2022;13(2):164–75. doi: 10.1002/jrsm.1531.

45. Stewart LA, Clarke M, Rovers M, Riley RD, Simmonds M, Stewart G, et al. Preferred Reporting Items for Systematic Review and Meta-Analyses of individual participant data: the PRISMA-IPD Statement. JAMA. 2015;313(16):1657–65. doi: 10.1001/jama.2015.3656.

46. Shea BJ, Bouter LM, Peterson J, Boers M, Andersson N, Ortiz Z, et al. External validation of a measurement tool to assess systematic reviews (AMSTAR). PLoS One. 2007;2(12):e1350. doi: 10.1371/journal.pone.0001350.

47. ISPOR: International Society for Pharmacoeconomics and Outcomes Research Available from: [https://www.ispor.org/home].

48. Richter T, Lee KM. Guidance Document on Reporting Indirect Comparisons. In: CADTH, editor. 2015.

49. Gao Y, Shi S, Li M, Luo X, Liu M, Yang K, et al. Statistical analyses and quality of individual participant data network meta-analyses were suboptimal: a cross-sectional study. BMC Med. 2020;18(1):120. doi: 10.1186/s12916-020-01591-0.

50. Williams T, Stein DJ, Ipser J. A systematic review of network meta-analyses for pharmacological treatment of common mental disorders. Evid Based Ment Health. 2018;21(1):7–11. doi: 10.1136/eb-2017-102718.

51. Donegan S, Williamson P, D’Alessandro U, Tudur Smith C. Assessing key assumptions of network meta-analysis: a review of methods. Res Synth Methods. 2013;4(4):291–323. doi: 10.1002/jrsm.1085.

52. Tan SH, Bujkiewicz S, Sutton A, Dequen P, Cooper N. Presentational approaches used in the UK for reporting evidence synthesis using indirect and mixed treatment comparisons. J Health Serv Res Policy. 2013;18(4):224–32. doi: 10.1177/1355819613498379.

53. Bafeta A, Trinquart L, Seror R, Ravaud P. Reporting of results from network meta-analyses: methodological systematic review. BMJ. 2014;348:g1741. doi: 10.1136/bmj.g1741.

54. Laws A, Kendall R, Hawkins N. A comparison of national guidelines for network meta-analysis. Value Health. 2014;17(5):642–54. doi: 10.1016/j.jval.2014.06.001.

55. Ortega A, Fraga MD, Alegre-del-Rey EJ, Puigventos-Latorre F, Porta A, Ventayol P, et al. A checklist for critical appraisal of indirect comparisons. Int J Clin Pract. 2014;68(10):1181–9. doi: 10.1111/ijcp.12487.

56. Sullivan SM, Coyle D, Wells G. What guidance are researchers given on how to present network meta-analyses to end-users such as policymakers and clinicians? A systematic review. PLoS One. 2014;9(12):e113277. doi: 10.1371/journal.pone.0113277.

57. Chambers JD, Naci H, Wouters OJ, Pyo J, Gunjal S, Kennedy IR, et al. An assessment of the methodological quality of published network meta-analyses: a systematic review. PLoS One. 2015;10(4):e0121715. doi: 10.1371/journal.pone.0121715.

58. Fleetwood K, Glanville J, McCool R, Wood H, Wilson K, Marshall C, et al. A Review of the Use of Network Meta-Analysis In Nice Single Technology Appraisals. Value in Health. 2016;19(7):348. doi: 10.1016/j.jval.2016.09.009.

59. Ge L, Tian JH, Li XX, Song F, Li L, Zhang J, et al. Epidemiology Characteristics, Methodological Assessment and Reporting of Statistical Analysis of Network Meta-Analyses in the Field of Cancer. Sci Rep. 2016;6:37208. doi: 10.1038/srep37208.

60. Veroniki AA, Straus SE, Soobiah C, Elliott MJ, Tricco AC. A scoping review of indirect comparison methods and applications using individual patient data. BMC Med Res Methodol. 2016;16:47. doi: 10.1186/s12874-016-0146-y.

61. Zarin W, Veroniki AA, Nincic V, Vafaei A, Reynen E, Motiwala SS, et al. Characteristics and knowledge synthesis approach for 456 network meta-analyses: a scoping review. BMC Med. 2017;15(1):3. doi: 10.1186/s12916-016-0764-6.

62. James A, Yavchitz A, Ravaud P, Boutron I. Node-making process in network meta-analysis of nonpharmacological treatment are poorly reported. J Clin Epidemiol. 2018;97:95–102. doi: 10.1016/j.jclinepi.2017.11.018.

63. Spineli LM, Yepes-Nunez JJ, Schunemann HJ. A systematic survey shows that reporting and handling of missing outcome data in networks of interventions is poor. BMC Med Res Methodol. 2018;18(1):115. doi: 10.1186/s12874-018-0576-9.

64. Tonin FS, Steimbach LM, Mendes AM, Borba HH, Pontarolo R, Fernandez-Llimos F. Mapping the characteristics of network meta-analyses on drug therapy: A systematic review. PLoS One. 2018;13(4):e0196644. doi: 10.1371/journal.pone.0196644.

65. Tonin FS, Borba HH, Leonart LP, Mendes AM, Steimbach LM, Pontarolo R, et al. Methodological quality assessment of network meta-analysis of drug interventions: implications from a systematic review. Int J Epidemiol. 2019;48(2):620–32. doi: 10.1093/ije/dyy197.

66. Tricco AC, Zarin W, Ghassemi M, Nincic V, Lillie E, Page MJ, et al. Same family, different species: methodological conduct and quality varies according to purpose for five types of knowledge synthesis. J Clin Epidemiol. 2018;96:133–42. doi: 10.1016/j.jclinepi.2017.10.014.

67. Yang F, Wang H, Zou J, Li X, Jin X, Cao Y, et al. Assessing the methodological and reporting quality of network meta-analyses in Chinese medicine. Medicine (Baltimore). 2018;97(47):e13052. doi: 10.1097/MD.0000000000013052.

68. Chen Y, Zeng XY, Liu DF, Tan XY, Cai XM, Yang FW, et al. [Critical quality evaluation and application value of network Meta-analyses in traditional Chinese medicine]. Zhongguo Zhong Yao Za Zhi. 2019;44(24):5322–8. doi: 10.19540/j.cnki.cjcmm.20191022.501.

69. Gao Y, Ge L, Ma X, Shen X, Liu M, Tian J. Improvement needed in the network geometry and inconsistency of Cochrane network meta-analyses: a cross-sectional survey. J Clin Epidemiol. 2019;113:214–27. doi: 10.1016/j.jclinepi.2019.05.022.

70. Lee A. Developing critical appraisal of systematic reviews reporting network meta-analysis: University of Oxford; 2020.

71. Pratt M, Wieland S, Ahmadzai N, Butler C, Wolfe D, Pussagoda K, et al. A scoping review of network meta-analyses assessing the efficacy and safety of complementary and alternative medicine interventions. Syst Rev. 2020;9(1):97. doi: 10.1186/s13643-020-01328-3.

72. Bae K, Shin IS. Critical evaluation of reporting quality of network meta-analyses assessing the effectiveness of acupuncture. Complement Ther Clin Pract. 2021;45:101459. doi: 10.1016/j.ctcp.2021.101459.

73. Veroniki AA, Tsokani S, Zevgiti S, Pagkalidou I, Kontouli KM, Ambarcioglu P, et al. Do reporting guidelines have an impact? Empirical assessment of changes in reporting before and after the PRISMA extension statement for network meta-analysis. Syst Rev. 2021;10(1):246. doi: 10.1186/s13643-021-01780-9.

74. Wright E, Yasmeen N, Malottki K, Sawyer LM, Borg E, Schwenke C, et al. Assessing the Quality and Coherence of Network Meta-Analyses of Biologics in Plaque Psoriasis: What Does All This Evidence Synthesis Tell Us? Dermatol Ther (Heidelb). 2021;11(1):181–220. doi: 10.1007/s13555-020-00463-y.

75. Yuan T, Xiong J, Wang X, Yang J, Jiang Y, Zhou X, et al. The Quality of Methodological and Reporting in Network Meta-Analysis of Acupuncture and Moxibustion: A Cross-Sectional Survey. Evid Based Complement Alternat Med. 2021;2021:2672173. doi: 10.1155/2021/2672173.

76. Nast A, Dressler C, Schuster C, Saure D, Augustin M, Reich K. Methods used for indirect comparisons of systemic treatments for psoriasis. A systematic review. Skin Health Dis. 2023;3(1):e112. doi: 10.1002/ski2.112.

77. Wang Y, Chen N, Guo K, Li Y, E F, Yang C, et al. Reporting and methodological quality of acupuncture network meta-analyses could be improved: an evidence mapping. J Clin Epidemiol. 2023;153:1–12. doi: 10.1016/j.jclinepi.2022.11.004.

78. de Sousa PG, Mainka FF, Tonin FS, Pontarolo R. Mapping the characteristics, methodological quality and standards of reporting of network meta-analyses on antithrombotic therapies: An overview. Int J Cardiol. 2023;386:125–33. doi: 10.1016/j.ijcard.2023.05.036.

79. Montagner AF, Angst PDM, Raggio DP, FH VDS, Tedesco TK. Methodological quality of network meta-analysis in dentistry: a meta-research. Braz Oral Res. 2023;37:e062. doi: 10.1590/1807-3107bor-2023.vol37.0062.

80. Nagendrababu V, Narasimhan S, Faggion CM, Jr., Dharmarajan L, Jacob PS, Gopinath VK, et al. Reporting quality of systematic reviews with network meta-analyses in Endodontics. Clin Oral Investig. 2023;27(7):3437–45. doi: 10.1007/s00784-023-04948-w.

81. Sehmbi H, Retter S, Shah UJ, Nguyen D, Martin J, Uppal V. Methodological and reporting quality assessment of network meta-analyses in anesthesiology: a systematic review and meta-epidemiological study. Can J Anaesth. 2023;70(9):1461–73. doi: 10.1007/s12630-023-02510-6.

82. Lovsletten PO, Wang X, Pitre T, Odegaard M, Veroniki AA, Lunny C, et al. A systematic survey of 200 systematic reviews with network meta-analysis (published 2020-2021) reveals that few reviews report structured evidence summaries. J Clin Epidemiol. 2024;173:111445. doi: 10.1016/j.jclinepi.2024.111445.

83. Spineli LM. An empirical study on 209 networks of treatments revealed intransitivity to be common and multiple statistical tests suboptimal to assess transitivity. BMC Med Res Methodol. 2024;24(1):301. doi: 10.1186/s12874-024-02436-7.

84. Phillippo DM, Dias S, Welton NJ, Caldwell DM, Taske N, Ades AE. Threshold Analysis as an Alternative to GRADE for Assessing Confidence in Guideline Recommendations Based on Network Meta-analyses. Ann Intern Med. 2019;170(8):538–46. doi: 10.7326/M18-3542.

85. Lunny C, Veroniki AA, Higgins JPT, Dias S, Hutton B, Wright JM, et al. Methodological review of NMA bias concepts provides groundwork for the development of a list of concepts for potential inclusion in a new risk of bias tool for network meta-analysis (RoB NMA Tool). Syst Rev. 2024;13(1):25. doi: 10.1186/s13643-023-02388-x.

86. Hummel N, Debray TPA, Didden EM, Efthimiou O, Egger M, Fletcher C, et al. Methodological guidance, recommendations and illustrative case studies for (network) meta-analysis and modelling to predict real-world effectiveness using individual participant and/or aggregate data. 2017 doi: 10.13140/RG.2.2.36349.36327.

87. Brignardello-Petersen R, Bonner A, Alexander PE, Siemieniuk RA, Furukawa TA, Rochwerg B, et al. Advances in the GRADE approach to rate the certainty in estimates from a network meta-analysis. J Clin Epidemiol. 2018;93:36–44. doi: 10.1016/j.jclinepi.2017.10.005.

88. Shi C, Westby M, Norman G, Dumville JC, Cullum N. Node-making processes in network meta-analysis of nonpharmacological interventions should be well planned and reported. J Clin Epidemiol. 2018;101:124–5. doi: 10.1016/j.jclinepi.2018.04.009.

89. Dias S, Ades AE, Welton NJ, Jansen JP, Sutton AJ. Network Meta-Analysis for Decision Making: Wiley; 2018.

90. Practical Guideline for Quantitative Evidence Synthesis: Direct and Indirect Comparisons. In: Commission E, editor. 2024.

91. Dwan K, Livingstone N. Editorial considerations in reviews with network meta-analysis 2020. Available from: [https://training.cochrane.org/resource/editorial-considerations-reviews-network-meta-analysis].

92. Chaimani A, Caldwell DM, Li T, Higgins JPT, Salanti G. Additional considerations are required when preparing a protocol for a systematic review with multiple interventions. J Clin Epidemiol. 2017;83:65–74. doi: 10.1016/j.jclinepi.2016.11.015.

93. Morton SC, Murad MH, O’Connor E, Lee CS, Booth M, Vandermeer BW, et al. Quantitative Synthesis-An Update. Methods Guide for Effectiveness and Comparative Effectiveness Reviews. AHRQ Methods for Effective Health Care. Rockville (MD)2008.

94. Chaimani A, Caldwell DM, Li T, Higgins JPT, Salanti G. Undertaking network meta-analyses. Cochrane Handbook for Systematic Reviews of Interventions. 2nd ed 2019.

95. Chaimani A, Salanti G, Leucht S, Geddes JR, Cipriani A. Common pitfalls and mistakes in the set-up, analysis and interpretation of results in network meta-analysis: what clinicians should look for in a published article. Evid Based Ment Health. 2017;20(3):88–94. doi: 10.1136/eb-2017-102753.

96. Welton NJ, Phillippo DM, Owen R, Jones HE, Dias S, Bujkiewicz S, et al. CHTE2020 SOURCES AND SYNTHESIS OF EVIDENCE; UPDATE TO EVIDENCE SYNTHESIS METHODS. 2020.

97. Jansen JP, Trikalinos T, Cappelleri JC, Daw J, Andes S, Eldessouki R, et al. Indirect treatment comparison/network meta-analysis study questionnaire to assess relevance and credibility to inform health care decision making: an ISPOR-AMCP-NPC Good Practice Task Force report. Value Health. 2014;17(2):157–73. doi: 10.1016/j.jval.2014.01.004.

98. Catala-Lopez F, Tobias A, Cameron C, Moher D, Hutton B. Network meta-analysis for comparing treatment effects of multiple interventions: an introduction. Rheumatol Int. 2014;34(11):1489–96. doi: 10.1007/s00296-014-2994-2.

99. Puhan MA, Schunemann HJ, Murad MH, Li T, Brignardello-Petersen R, Singh JA, et al. A GRADE Working Group approach for rating the quality of treatment effect estimates from network meta-analysis. BMJ. 2014;349:g5630. doi: 10.1136/bmj.g5630.

100. Moher D, Collins G, Hoffmann T, Glasziou P, Ravaud P, Bian ZX. Reporting on data sharing: executive position of the EQUATOR Network. BMJ. 2024;386:e079694. doi: 10.1136/bmj-2024-079694.

