## Appendices Document for "Updating the PRISMA reporting guideline for network meta-analysis: a scoping review"

### Appendix Supplementary Content

#### Appendix 1: Protocol amendments

##### Updating the PRISMA reporting guideline for network meta-analysis: a scoping review

| Category | Deviation | Notes |
|---|---|---|
| Exclusion criteria | Post-hoc exclusion criteria of excluding citations published before the 2014 scoping review search,[1] specifically published before the August 2013 time point. | Since this review builds on a 2014 scoping review by our team,[1] which guided the development of PRISMA-NMA,[2] we limited the inclusion to studies published after the search date of the previous review (August 2013). |

#### Appendix 2: Lay summary

This is a scoping review, a type of knowledge synthesis that uses a systematic approach to assess the extent of available evidence on a specific topic, to organise it into groups and to highlight gaps. This scoping review is the first step in a process to update a reporting guideline for systematic reviews (a type of knowledge synthesis) where a method known as a network meta-analysis is used: the Preferred Reporting Items for Systematic reviews and Meta-Analyses (PRISMA) extension for network meta-analysis (NMA) (PRISMA-NMA). It is an update to a 2014 scoping review to identify additional, more recent studies. These studies will then form the basis of a series of surveys (known as a Delphi study) to reach consensus on what should be included in the PRISMA-NMA reporting guideline. This update will align with current methodological evidence, include the perspectives of patients and the public, and offer authors clear guidance on what information should be reported and how they should do this. The project involves researchers, healthcare professionals, journal editors, and patients, caregivers and members of the public. A comprehensive search across multiple databases was conducted resulting in the inclusion of 61 studies, comprising 23 guidance documents and 38 overviews of reviews evaluating the completeness of reporting and the methodological quality of published NMAs. This resulted in an additional 37 NMA items for inclusion in the next step—the Delphi study. This scoping review underscores the importance of updating the PRISMA-NMA reporting guideline to address gaps in reporting and align with advancements in what to report and how best to do this.

#### Appendix 3: Database search strategies

**PRISMA**

**Search strategies**

**November 22, 2023**

**OVID databases included**:

- MEDLINE (Primary database)
- Cochrane Methodology Register
- Embase
- ERIC

**Ovid MEDLINE(R) ALL <1946 to November 21, 2023>**

**--------------------------------------------------------------------------------**

1 Network meta-analysis/ (5486)
2 ((network* or network-based or "mixed treatment" or mixed-treatment or "multiple treatment comparison" or mtc) adj2 (meta-analys#s or metaanalys#s or meta analys#s or "meta regression" or meta-regression)).tw,kf. (10051)
3 ((Indirect comparison* or indirect treatment* or bayesian) adj2 (meta-analys#s or metaanalys#s or meta analys#s or "meta regression" or meta-regression)).tw,kf. (2738)
4 (Indirect comparison* or indirect treatment* or mixed-treatment or mixed treatment or bayesian).tw,kf. and (Review Literature as Topic/ or meta-analysis as topic/ or systematic review as topic/ or *Matched-Pair Analysis/ or Technology Assessment, Biomedical/) (878)
5 ((multiparamet* adj2 evidence adj2 synthesis) or (multi-paramet* adj2 evidence adj2 synthesis)).tw,kf. (30)
*6 or/1-5 (11749)*
7 report*.ab. /freq=3 or report*.kf. (459722)
8 Publishing/ or Open Access Publishing/ or Periodicals as Topic/ or exp checklist/ or Publication Bias/ (89201)
9 Research Design/ and (mt or st).fs. (50941)
10 ((journal or periodical or publication or publish* or presentation) adj2 (report* or bias* or requirement* or adherence or compliance or guideline* or recommendation* or standard* or guidance or instruction* or checklist* or check list* or evaluat*)).tw,kf. (77693)
11 ((clear* or fully or adequately or inadequately or completely or incompletely or poor* or transparent* or method* or quality or element* or requirement* or guideline* or recommendation* or standard* or guidance or instruction* or assess* or apprais* or bias* or characteristic* or criteri* or critiqu* or evaluat* or quality or checklist* or check list* or score$1 or scoring or adherence or compliance or approach* or item* or measure or measures) adj2 (report* or conduct)).tw,kf. (228485)
*12 or/7-11 (811148)*
**13 6 and 12 (1497) – NMA – download this line**

**Embase Classic+Embase <1947 to 2023 November 21>**

**--------------------------------------------------------------------------------**

1 report*.ab. /freq=3 or report*.kf. (632076)
2 publishing/ or open access publishing/ or publication bias/ (66704)

3 ((journal or periodical or publication or publish* or presentation) adj (report* or bias* or requirement* or adherence or compliance or guideline* or recommendation* or standard* or guidance or instruction* or checklist* or check list* or evaluat*)).tw,kf. (59267)
4 ((clear* or fully or adequately or inadequately or completely or incompletely or poor* or transparent* or method* or quality or element* or requirement* or guideline* or recommendation* or standard* or guidance or instruction* or assess* or apprais* or bias* or characteristic* or criteri* or critiqu* or evaluat* or quality or checklist* or check list* or score$1 or scoring or adherence or compliance or approach* or item* or measure or measures) adj (report* or conduct)).tw,kf. (34595)
5 or/1-4 (760495)
6 network meta-analysis/ (7955)
7 ((network* or network-based or "mixed treatment" or mixed-treatment or "multiple treatment comparison" or mtc) adj2 (meta-analys#s or metaanalys#s or meta analys#s or "meta regression" or meta-regression)).tw,kf. (13957)
8 ((Indirect comparison* or indirect treatment* or bayesian) adj2 (meta-analys#s or metaanalys#s or meta analys#s or "meta regression" or meta-regression)).tw,kf. (3685)
9 6 or 7 or 8 (15570)
**10 5 and 9 (1293) – NMA – download this line**

**EBM Reviews - Cochrane Methodology Register <3rd Quarter 2012>**
--------------------------------------------------------------------------------
1 report*.ab. /freq=3 or report*.kf. (1597)
2 ((journal or periodical or publication or publish* or presentation) adj2 (report* or bias* or requirement* or adherence or compliance or guideline* or recommendation* or standard* or guidance or instruction* or checklist* or check list* or evaluat*)).tw,kf. (1409)
3 ((clear* or fully or adequately or inadequately or completely or incompletely or poor* or transparent* or method* or quality or element* or requirement* or guideline* or recommendation* or standard* or guidance or instruction* or assess* or apprais* or bias* or characteristic* or criteri* or critiqu* or evaluat* or quality or checklist* or check list* or score$1 or scoring or adherence or compliance or approach* or item* or measure or measures) adj2 (report* or conduct)).tw,kf. (1403)
4 or/1-3 (3031)
5 ((network* or network-based or "mixed treatment" or mixed-treatment or "multiple treatment comparison" or mtc) adj2 (meta-analys#s or metaanalys#s or meta analys#s or "meta regression" or meta-regression)).tw,kf. (26)
6 ((Indirect comparison* or indirect treatment* or bayesian) adj2 (meta-analys#s or metaanalys#s or meta analys#s or "meta regression" or meta-regression)).tw,kf. (177)
7 5 or 6 (185)
**8 4 and 7 (19)- NMA – download this line**

**ERIC <1965 to September 2023>**
--------------------------------------------------------------------------------
1 report*.ab. /freq=3 (39744)
2 ((journal or periodical or publication or publish* or presentation) adj2 (report* or bias* or requirement* or adherence or compliance or guideline* or recommendation* or standard* or guidance or instruction* or checklist* or check list* or evaluat*)).tw. (4800)
3 ((clear* or fully or adequately or inadequately or completely or incompletely or poor* or transparent* or method* or quality or element* or requirement* or guideline* or recommendation* or standard* or guidance or instruction* or assess* or apprais* or bias* or characteristic* or criteri* or critiqu* or evaluat* or quality or checklist* or check list* or score$1 or scoring or adherence or compliance or

approach* or item* or measure or measures) adj2 (report* or conduct)).tw. (28836)
4 or/1-3 (65747)
5 ((network* or network-based or "mixed treatment" or mixed-treatment or "multiple treatment comparison" or mtc) adj2 (meta-analys#s or metaanalys#s or meta analys#s or "meta regression" or meta-regression)).tw. (59)
6 ((Indirect comparison* or indirect treatment* or bayesian) adj2 (meta-analys#s or metaanalys#s or meta analys#s or "meta regression" or meta-regression)).tw. (33)
7 5 or 6 (84)
**8 4 and 7 (6) NMA – download this line**

#### Appendix 4: Level 1 screening guidelines

**PRISMA-NMA Update: L1 Screening Cheat Sheet**

**Website:** https://synthesi.sr/web/login
**Project Name (synthesi.SR):** PRISMA-NMA Update

**Research question**: What are the items that should be reported in network meta-analysis (NMA) in order to be consistent with current best evidence?

**Objective:** To update the PRISMA-NMA reporting guideline to reflect current evidence.

**<u>IMPORTANT NOTES:</u>**

- ***If you answer <u>NO</u> to any of <u>Questions 1-2</u>, the citation will be <u>EXCLUDED</u>***
- ***If you answer <u>YES</u> and/or <u>UNCLEAR</u> to any of Questions 1-2, the citation will be <u>INCLUDED.</u>***
    - ***In this case, please make sure to answer Questions 3 and 4, as they are <u>FLAGGING</u> questions.***

| ***Question 1: Does the study provide guidance on reporting or quality assessment of NMA? (*mandatory)** | |
|---|---|
| **Responses** | • **YES**<br>• **NO**<br>• **UNCLEAR** |
| **Notes** | • **INCLUDE (Yes/ Unclear)** if the study presents:<br> o Guidance/tutorial/guideline relevant to reporting in NMA<br> ▪ **Note:** These may include a *checklist*, *flow diagram* or *text* to guide authors in NMA reporting<br> o Evaluation of reporting completeness in NMA<br> o Evaluation of reporting quality in NMA<br> o Evaluation of sources of bias in NMA<br> o Evaluation of risk of bias in NMA<br> o Evaluation of methodological quality in NMA<br><br>• **INCLUDE** and **FLAG** (using protocol option in Q4):<br> o Protocols for relevant methodological studies<br> o E.g., protocol for a Delphi survey is considered for inclusion but protocols of applied NMAs that answer research PICO questions are to be excluded.<br>• **NOTE:** Studies exclusively focused on Diagnostic Test Accuracy (DTA) are not of interest. However, studies that mention the guidelines listed below are to be moved to L2 screening for a full and comprehensive review of eligibility:<br> o EQUATOR, PRISMA, CIHR, Agency for Healthcare Research and Quality [AHRQ], Guidelines International Network [GIN], National Institute for Health and Care Excellence [NICE], Canadian Agency for Drugs and Technologies in Health [CADTH], eunethta, Health Information and Quality Authority, Belgian Health Care Knowledge Centre [KCE], German Institute for Quality and Efficiency in Health Care [IQWIG]). |

| **Question 2: Does the study relate to human health (including psychology, education and sociology disciplines) and/or philosophy?** | |
|---|---|
| **Responses** | • **YES** |

| | |
|---|---|
| | • **NO**<br>• **UNCLEAR** |
| Notes | • **According to the World Health Organization, health is defined as 'a state of complete physical, mental and social well-being'.**<br><br>• **INCLUDE anything that relates to human health, including:**<br> ○ Public health<br> ○ Nutritional sciences<br> ○ Environmental sciences<br> ○ Social sciences<br> ○ Health service administration<br> ○ Health Promotion |

| **Question 3: Which of the following types does the study entail? Check all that apply. [Flagging Question]** | |
|---|---|
| Responses | • **Guidance/tutorial/guideline on reporting in NMA**<br>• **Guidance/tutorial/guideline on methodological quality in NMA**<br>• **Evaluation of reporting completeness or quality in NMA**<br>• **Evaluation of methods quality/risk of bias/source of bias in NMA**<br>• **Unclear/Unsure** |

| **Question 4: Is this potentially relevant, but in one of the following formats? Check all that apply. [Flagging Question]** | |
|---|---|
| Responses | • **Protocol**<br>• **Conference abstract**<br>• **Non-English**<br>• **Other (please make a note in the comments to explain)** |

#### Appendix 5: Level 2 screening guidelines

**PRISMA-NMA Update: L2 Screening Cheat Sheet**

**Website:** https://synthesi.sr/web/login
**Project Name (synthesi.SR):** PRISMA-NMA Update

**Research question**: What are the items that should be reported in network meta-analysis (NMA) in order to be consistent with current best evidence?

**Objective:** To update the PRISMA-NMA reporting guideline to reflect current evidence.

**IMPORTANT NOTES:**

- ***If you answer NO to any of Questions 1-3, the citation will be EXCLUDED***
- ***If you answer YES and/or UNCLEAR to any of Questions 1-3, the citation will be INCLUDED.***
    - ***In this case, please make sure to answer Questions 4 and 5, as they are FLAGGING questions.***
- ***If the article is not in English, please respond UNCLEAR to Questions 1-4 and indicate the appropriate non-English article option in Question 5.***

| ***Question 1: Is this study about NMAs? [*MANDATORY]** | |
|---|---|
| **Responses** | • **YES**<br>• **NO**<br>• **UNCLEAR** |
| **Notes** | • **INCLUDE (Yes/ Unclear)** if the study presents:<br>○ **Key Definition:** Network meta-analysis (NMA) is a technique for comparing three or more interventions simultaneously in a single analysis by combining both direct and indirect evidence across a network of studies.<br>○ **Include** any approaches that are about NMA or indirect comparisons, including matching-adjusted indirect comparison (MAIC), simulated treatment comparison (STC), mixed/multiple treatment comparison (MTC) and multiple treatment meta-analysis (MTM) are eligible.<br><br>• **NOTE:** NMAs of any type of data, such as, individual patient data (IPD) and/or aggregate data, are eligible. |

| ***Question 2: Is this study about evaluating or providing guidance on reporting or quality assessment of NMAs?** | |
|---|---|
| **Responses** | • **YES**<br>• **NO**<br>• **UNCLEAR**<br>• **DOES NOT MEET INCLUSION CRITERIA, BUT INCLUDES RELEVANT METHODOLOGY FOR NMAS** |

| | |
|---|---|
| **Notes** | • **INCLUDE (Yes/ Unclear)** if the study presents:<br> o Guidance/tutorial/guideline relevant to reporting in NMA<br> ▪ **Note:** These may include a *checklist*, *flow diagram* or *text* to guide authors in NMA reporting<br> ▪ **Include** editorial guidelines or tutorials that describe items related to reporting completeness (e.g., in the World Association of Medical Editors [WAME], International Committee of Medical Journal Editorial [ICMJE], and Council of Science Editors [3])<br> o Evaluation of reporting completeness in NMA<br> o Evaluation of reporting quality in NMA<br> o Evaluation of sources of bias in NMA<br> o Evaluation of risk of bias in NMA<br> o Evaluation of methodological quality in NMA<br><br>• **INCLUDE** and **FLAG** (using protocol option in Q5):<br> o Protocols for relevant methodological studies<br> o E.g., protocol for a Delphi survey is considered for inclusion but protocols of applied NMAs that answer research PICO questions are to be excluded.<br><br>• **EXCLUDE:**<br> o Commentaries on reporting guidelines and any publications describing manuscript formatting or journal guidance to authors<br><br>• **DOES NOT MEET INCLUSION CRITERIA, BUT INCLUDES RELEVANT METHODOLOGY FOR NMAS:**<br> o **NOTE:** Relevant methodology studies for NMAs will not be considered for inclusion in the present review. However, if deemed beneficial to consider in the Elaboration and Explanation (E&E) document, please select this option.<br> o **Example:** A study evaluating a method to deal with missing data and publication bias in a NMA is not eligible.<br> ▪ The reason is that the study is NOT actually providing guidance or evaluating the reporting, and/or quality of NMAs, rather they present/discuss methods to address these issues.<br><br>• **NOTE:** Studies exclusively focused on Diagnostic Test Accuracy (DTA) are not of interest.<br>• **NOTE:** Studies that mention relevant guidelines listed below are eligible:<br> o EQUATOR, PRISMA, CIHR, Agency for Healthcare Research and Quality [AHRQ], Guidelines International Network [GIN], National Institute for Health and Care Excellence [NICE], Canadian Agency for Drugs and Technologies in Health [CADTH], eunethta, Health Information and Quality Authority, Belgian Health Care Knowledge Centre [KCE], German Institute for Quality and Efficiency in Health Care [IQWIG]).<br>• **NOTE:** We are interested in guidance of reporting or quality assessment of NMAs of intervention. |

| | |
|---|---|
| | ○ **Example:** PRISMA-NMA checklist and ISPOR |

| **Question 3: Does the study relate to human health (including psychology, education and sociology disciplines) and/or philosophy?** | |
|---|---|
| **Responses** | • **YES**<br>• **NO**<br>• **UNCLEAR** |
| **Notes** | • **Key Definition:** According to the World Health Organization, health is defined as 'a state of complete physical, mental and social well-being'.<br><br>• **INCLUDE anything that relates to human health, including:**<br>○ Public health<br>○ Nutritional sciences<br>○ Environmental sciences<br>○ Social sciences<br>○ Health service administration<br>○ Health Promotion |

| **Question 4: Which of the following types does the study entail? Check all that apply. [Flagging Question]** | |
|---|---|
| **Responses** | • Guidance/tutorial/guideline on reporting in NMA<br>• Guidance/tutorial/guideline on methodological quality in NMA<br>• Evaluation of reporting completeness or quality in NMA<br>• Evaluation of methods quality/risk of bias/source of bias in NMA<br>• Unclear/Unsure |

| **Question 5: Is this potentially relevant, but in one of the following formats? Check all that apply. [Flagging Question]** | |
|---|---|
| **Responses** | • Protocol<br>• Conference abstract<br>• Non-English<br>• Systematic Review<br>• Other (please make a note in the comments to explain) |

#### Appendix 6: List of excluded studies post-level 2 screening

##### Excluded studies after completion of L2 screening with Reasons

| Study | Last Name of First Author | Reason for exclusion |
|---|---|---|
| ROB-MEN: a tool to assess risk of bias due to missing evidence in network meta-analysis | Chiocchia | Exclude-Duplicate |
| Do reporting guidelines have an impact? Empirical assessment of changes in reporting before and after the PRISMA extension statement for network meta-analysis | Veroniki | Exclude-Duplicate |
| Bibliographic study showed improving statistical methodology of network meta-analyses published between 1999 and 2015 | Petropoulou | Exclude Q2 |
| Bibliographic study showed improving statistical methodology of network meta-analyses published between 1999 and 2015 | Petropoulou | Exclude-Duplicate |
| A Comparison of National Guidelines for Network Meta-Analysis | Laws | Exclude-Duplicate |
| Reporting of Cochrane systematic review protocols with network meta-analyses-A scoping review | Wang | Exclude-Duplicate |
| Network meta-analysis: users' guide for pediatricians | Al Khalifah | Exclude Q2 |
| A meta-epidemiological survey of the reporting of effect modification in network meta-analyses | Kovic | Exclude-Duplicate |
| Reporting quality & transparency of published network meta-analysis. is it satisfactory? | Kopiec | Exclude-Not Retrievable |
| Methodological problems in the use of indirect comparisons for evaluating healthcare interventions: survey of published systematic reviews | Song | Exclude-Published before Aug2013 |
| A clinician's guide to network meta-analysis | Phillips | Exclude Q2 |
| Faulty connections: Can criticisms of network meta-analysis in nice submissions be avoided? | Martin | Exclude-Not Retrievable |
| Meta-analysis to support technology submissions to health technology assessment authorities: Criticisms by nice and evidence review groups in the UK | Batson | Exclude-Not Retrievable |
| Network meta-analysis for health technology submissions worldwide: A report checklist for network meta analysis best practices globally | Batson | Exclude-Not Retrievable |
| Are industry funded network meta-analyses lower quality? | Chambers | Exclude-Not Retrievable |
| Network meta-analyses: Methodological prerequisites and clinical usefulness | Christofilos | Exclude Q2 |
| PNS147 VISUALIZATION AND COMMUNICATION OF RESULTS FROM BAYESIAN NETWORK META-ANALYSIS: PAST, PRESENT, AND FUTURE | Eaton | Exclude-Not Retrievable |
| Are the searches of network meta-analysis comprehensive or well-reported? Poster | Tian | Exclude-Not Retrievable |

| | | |
|---|---|---|
| presentation at the 19th Cochrane Colloquium; 2011 Oct 19-22; Madrid, Spain [abstract] | | |
| Assessing the quality of report in network meta-analysis: A systematic review | Tonin | Exclude-Not Retrievable |
| Use of network meta-analysis in systematic reviews: a survey of authors | Lee | Exclude Q2 |
| Advance in the grade approach to rate the quality of evidence from a network meta-analysis | Wang | Exclude Q2 |
| Indirect Comparisons and Network Meta-Analyses | Kiefer | Exclude Q2 |
| A systematic review of the methodological quality of network meta-analyses | Chambers | Exclude-Not Retrievable |
| Indirect comparisons: a review of reporting and methodological quality | Donegan | Exclude-Published before Aug2013 |
| Methodological and reporting quality of indirect comparisons. Poster presentation at the 16th Cochrane Colloquium: Evidence in the era of globalisation; 2008 Oct 3-7; Freiburg, Germany [abstract] | Donegan | Exclude-Published before Aug2013 |
| Analysis of the systematic reviews process in reports of network meta-analyses: methodological systematic review | Bafeta | Exclude-Published before Aug2013 |
| Method's corner: Allergist's guide to network meta-analysis | Chu | Exclude Q2 |
| [Critical quality evaluation and application value of network Meta-analyses in traditional Chinese medicine] | Chen | Exclude-Not Translatable/Retrievable |
| Network Meta-analysis: Users' Guide for Surgeons: Part II - Certainty | Chaudhry | Exclude Q2 |
| Network Meta-analysis: Users' Guide for Surgeons: Part I - Credibility | Foote | Exclude Q2 |
| Use of Mixed Treatment Comparisons in Systematic Reviews | Coleman | Exclude-Published before Aug2013 |
| Characteristics of network meta-analyses presented at ICPE and ISPOR | Fusco | Exclude-Not Retrievable |
| Updated method guidelines for cochrane musculoskeletal group systematic reviews and metaanalyses | Ghogomu | Exclude Q2 |
| Network meta-analysis: an introduction for clinicians | Rouse | Exclude Q2 |
| What are the reporting and methodological qualities of network meta-analysis? Poster presentation at the 19th Cochrane Colloquium; 2011 Oct 19-22; Madrid, Spain [abstract] | Li | Exclude-Published before Aug2013 |
| Methods used to conduct and report Bayesian mixed treatment comparisons published in the medical literature: a systematic review | Sobieraj | Exclude-Published before Aug2013 |
| [Report quality evaluation of systematic review or Meta-analysis published in China Journal of Chinese Materia Medica] | Zhang | Exclude Q1 |
| Indirect comparisons for evaluation healthcare interventions: review of published systematic reviews and discussion of methodological problems. Oral presentation at the 16th | Song | Exclude-Published before Aug2013 |

| |
|---|
| Cochrane Colloquium: Evidence in the era of globalisation; 2008 Oct 3-7; Freiburg, Germany [abstract] |

#### Appendix 7: Data abstraction form guidelines

### PRISMA-NMA Extension Update

##### Data Abstraction Cheat Sheet – V6

**Research question**: What are the items that should be reported in network meta-analysis (NMA) in order to be consistent with current best evidence?

**Objective:** To update the PRISMA-NMA reporting guideline to reflect current evidence.

##### Overview

**NOTES**

Please enter not applicable (NA), not reported (NR), or "grey out" cells as needed instead of leaving cells blank.

**NOTE:** For all includes, if there are no new findings to data abstract, please state this so that we can clearly convey this in our report.

**Tracking changes of previously collected data:** We will follow the usual data verification approach. Feel free to delete or change information you do not agree with and change the font colour to red to indicate that the data has been changed.

**Comments field:** This field repeats in every tab. If you wish to leave overall comments about the study not captured elsewhere, please add all your comments here.

To enter a comment in a cell, right click and select "insert a comment."

**TAB 1. Study Characteristics**

| STUDY CHARACTERISTICS | |
|---|---|
| **Excel column** | **Description** |
| RefID | Please enter the RefID. |
| Reviewer | Please enter your initials.<br>**Example:** Areti-Angeliki Veroniki (AAV) |
| Last Name of First Author | Enter the last name of the first author.<br>**Example:** Smith |
| Publication Year | Enter the year the study was published.<br>**Example:** 2016 |
| Title | Enter the name of the study title as it appears in the full-text |
| Country | Enter the country as per corresponding author.<br>If the study doesn't mention the country but mentions a region, enter the region (e.g., Sub-Saharan Africa). Do not enter cities.<br>**Example:** USA, Canada, Netherlands |
| Source | Select the publication type.<br>☐ Journal article<br>☐ Report<br>☐ Other |
| Name of Publication Source | Enter the name of the publication source.<br><br>**Example:** BMJ, Cochrane Library<br>**Note:** If you selected "Other" for "Source", please specify the publication type and source here. |
| Funding | Please indicate if authors describe funding using the dropdown options.<br>☐ Yes<br>☐ No<br>☐ Unclear<br>☐ Not Reported<br><br>**Note**: Specify "No" if nothing is declared under the "Disclaimers", "Acknowledgment" or similar sections of the paper or if it is specifically stated that the study did not receive funding. Otherwise, select "Not reported". If there is still uncertainty, then select "Unclear". |
| Funding Source | Enter all sources of funding support for the research. Try to be succinct as possible.<br><br>**Example:** CIHR; Bayer. Enter 'None' for no funding source. |
| Funding Source Type | Select the category of the research sponsor type from the dropdown.<br>☐ Publicly Sponsored (research grants, government or agency commissioned)<br>☐ Industry Sponsored (pharma or any other industry) |

| | |
|---|---|
| | ☐ Public & Industry Sponsored<br>☐ Not Sponsored (if the paper specifically states that no funding was received)<br>☐ Not Reported<br><br>**Note:** If a paper reports internal funding, but the institution itself is an industry or mix of both public and industry then select "Public & Industry Sponsored".<br>**Tip:** If unclear, a quick Google search of the funding source name should help you choose the most appropriate source category. Funding from health insurers and private healthcare industries will fall under industry-sponsored. |
| Study design | Select the study design from the dropdown.<br>☐ Any type of review<br>☐ Guidance Document/Other<br><br>**Note:** Studies might not always explicitly state the design in which case select the best option based on the methods section. If unclear, please include any relevant information in the comment column at the end of the tab. If a study serves both as a review and a guidance document, please prioritize selecting "review" as the study design as to capture the most details.<br><br>This is important to distinguish as it will inform the subsequent tabs you complete. Please ensure to complete the last tab if you have checked off "any type of review". If you have selected "Guidance Document/Other", please proceed to completing the second tab. |
| Specify Other Study Design | If you selected "Guidance Document/Other" but believe it doesn't meet the criteria of a guidance document, please specify details on the study design here.<br><br>**Example**: Report, Case Study |
| Study Type | Please categorize the study type using the dropdown.<br>☐ Guidance/tutorial/guideline on reporting in NMA<br>☐ Guidance/tutorial/guideline on methodological quality in NMA<br>☐ Evaluation of reporting completeness or quality in NMA<br>☐ Evaluation of methods quality/risk of bias/source of bias in NMA |
| Additional Comments | Enter any relevant comments for the study that are not captured elsewhere here |

**TAB 2. Reporting_Method Qual_Guidance**

Our **main goal** is to extract all relevant information in accordance with the PRISMA guidelines for systematic reviews with network meta-analyses. Please abstract pertinent details concisely to avoid overlooking any important information. If you're uncertain about what to abstract, consult the supplementary **PRISMA NMA checklist (Supplement 1A)**[1] provided below for guidance on the type of information to include. It is anticipated that certain sections, such as introductions, abstracts, and select sections within the methods, will adhere to the core **PRISMA 2020 checklist (Supplement 1B)**[2], with no nuances specific to NMA. Any relevant points from PRISMA 2020 should be included in the 'Additional Comments' section. Only include the recommended NMA reporting in the columns provided.

| **Guidance/tutorial/guideline on reporting or methodological quality in NMA** | |
|---|---|
| **Excel column** | **Description** |
| RefID | Please enter the RefID. |
| Reviewer | Please enter your initials.<br><br>**Example:** Areti-Angeliki Veroniki (AAV) |
| Target Audience | Select the target audience using the dropdown.<br>☐ Researchers (including technology and information specialists)<br>☐ Healthcare and Allied Care Professionals (including managers, program planners, administrators)<br>☐ Government authorities and policy-makers<br>☐ Public Health Professionals (e.g., Epidemiologist, Health Promotion Specialists)<br>☐ Patients and Community Members (e.g., community partners, consumers, patients in the public, caregivers or carers)<br>☐ Educators<br>☐ Social and Community Outreach Worker<br>☐ Funding bodies<br>☐ >1 Target Audience<br>☐ Not Specified (no target audience specified by authors)<br><br>**TIP:** This information is usually found in the introduction or discussion sections.<br>**Note:** Please select the options only if the authors have specified the target audience(s). If no target audience was specified, please select "Not Specified". |
| Target Audience (Open Text) | If you selected ">1 Target Audience". Enter all applicable target audience alphabetically.<br><br>If no target audience was specified, please list all relevant target audiences alphabetically you think this may be relevant to.<br><br>**Example:** Not specified but likely for: Public Health Professionals, |

| | |
|---|---|
| | Researchers |
| **Specific to Study Designs of Any Type of Review** | |
| Inclusion Criteria | Enter the inclusion criteria of the review. |
| Timeframe of Assessed Literature | Enter when these NMAs were published. |
| # of NMAs Included | Enter the number of NMAs included in the review. |
| Conflicts of Interests | Enter if any conflicts of interest in the review was declared. |
| Reporting/Methodological Guideline Used (if any) | Enter if any reporting/methodological guideline was used, if any.<br><br>**Example**: PRISMA, ISPOR, GRADE, CINeMA |
| **Items Relevant to Title** | |
| Title | Enter suggested reporting information relevant to title. |
| Other | Enter any additional comments relevant to the title. |
| **Items Relevant to Abstract** | |
| Abstract | Enter suggested reporting information relevant to abstract. |
| Other | Enter any additional comments relevant to the abstract. |
| **Items Relevant to Introduction** | |
| Rationale | Enter suggested reporting information relevant to the rationale. |
| Objectives | Enter suggested reporting information relevant to the objectives. |
| Other | Enter any additional comments relevant to the introduction. |
| **Items Relevant to Methods** | |
| Protocol & Registration | Enter suggested reporting information relevant to the protocol and registration. |
| Eligibility Criteria | Enter suggested reporting information relevant to the eligibility criteria. |
| Information Source | Enter suggested reporting information relevant to the information source. |
| Search | Enter suggested reporting information to the search. |
| Data Collection Process | Enter suggested reporting relevant to the data collection process. |
| Data Items | Enter suggested reporting relevant to the data items. |
| Geometry of the Network | Enter suggested reporting relevant to the geometry of the network. |

| | |
|---|---|
| | **Definition of Network Geometry:** Used to refer to the architecture of the treatment comparisons involved in the studies of a network meta-analysis. This encompasses the treatments included in the comparisons, the distribution of studies across the comparisons, and the allocation of patients to each treatment.[2] |
| Risk of Bias within Individual Studies | Enter suggested reporting relevant to the risk of bias associated with the individual studies/trials included in the network.<br><br>**Note:** Risk of bias (ROB) within *individual studies* uses tools such as ROB2 and ROBINS-I tool. This is different from assessing ROB across studies. |
| Summary Measures | Enter suggested reporting relevant to the summary measures. |
| Planned Methods for Analysis | Enter suggested reporting relevant to the planned methods for analysis.<br><br>**Example:** Handling of multi-arm trials, selection of variance structure, heterogeneity and its assessment, selection of prior distributions in Bayesian analyses, assessment of model fit. |
| Assessment of Inconsistency | Enter suggested reporting relevant to the assessment of inconsistency.<br><br>**Definition of Inconsistency (or incoherence):** Inconsistency occurs when different sources of information within the network for a particular treatment effect disagree. We usually statistically assess consistency between direct and indirect evidence.[3] |
| Risk of Bias Across Studies | Enter suggested reporting relevant to the risk of bias across studies/ trials in the network.<br><br>**Note**: Risk of bias across studies can include using funnel plot or comparison-adjusted funnel plot to assess publication bias and small-study effects, Egger's test, Begg's test, to name a few. |
| Certainty of Evidence | Enter suggested reporting relevant to the certainty (confidence) of evidence. |
| Additional Analyses | Enter suggested reporting relevant to the additional analyses.<br><br>**Note:** Additional analyses can include sensitivity analysis, subgroup analysis, network meta-regression, individual participant data analyses. |
| Other | Enter any additional comments relevant to the methods.<br><br>For instance, if recommendations regarding the assessment and reporting of transitivity, assumptions of homogeneity, similarity, and consistency are present, or if there are considerations regarding visual representations of results, please include relevant details in this section. |
| **Items Relevant to Results** | |
| Study Selection | Enter suggested reporting relevant to the study selection. |

| | |
|---|---|
| Presentation of Network Structure | Enter suggested reporting relevant to the presentation of network structure.<br><br>**Note:** The presentation of network structure can also incorporate tables in addition to, or as a supplement to, network plots with information on treatment comparisons, studies, and participants.<br><br>**Note:** Generic figure of a network plot.[2]<br><br>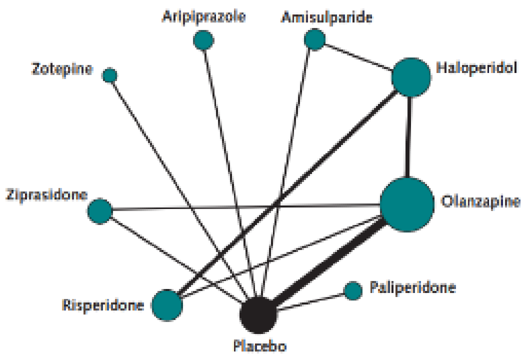<br> |
| Summary of Network Geometry | Enter suggested reporting relevant to the summary of network geometry<br><br>**Note:** Summary of network geometry refers to description of network geometry and evidence base of studies. |
| Study Characteristics | Enter suggested reporting relevant to the study characteristics.<br><br>**Note:** As in the PRISMA 2020 checklist, this refers to PICOS-related information, study time frame, sample size, patient demographics. This is often presented in tables and frequency plots. |
| Risk of Bias within Individual Studies | Enter suggested reporting relevant to the risk of bias within individual studies.<br><br>**Note:** Findings from risk of bias within individual studies can be presented in tables, "traffic light" plots, weighted bar plots of the distribution of risk-of-bias judgements, to name a few. |
| Results of Individual Studies | Enter suggested reporting relevant to the results of individual studies.<br><br>**Note:** Results of individual studies can be presented in tables or plots with individual study results. |
| Synthesis of Results | Enter suggested reporting relevant to the synthesis of results.<br><br>**Note:** Summaries of findings can be presented in various formats, such as tables of pairwise and network meta-analysis results, league tables, forest plots, interval plots, SUCRA plots, rank-heat plots, and kilim plots. |
| Exploration for Inconsistency | Enter suggested reporting relevant to the exploration for inconsistency.<br><br>**Definition of Inconsistency (or incoherence):** Inconsistency occurs when different sources of information within the network for a particular treatment effect disagree. We usually statistically assess consistency |

| | |
|---|---|
| | between direct and indirect evidence.[3]<br><br>**Note:** Presenting inconsistency results can include using loop-specific approach, Bucher method, node (or side) splitting method, design by treatment interaction method, Lu and Ades method, to name a few, |
| Risk of Bias Across Studies | Enter suggested reporting relevant to the risk of bias across studies.<br><br>**Note**: Risk of bias across studies can include using funnel plot or comparison-adjusted funnel plot to assess publication bias and small-study effects, Egger's test, Begg's test, to name a few. |
| Certainty of Evidence | Enter suggested reporting relevant to the certainty (confidence) of evidence. |
| Results of Additional Analyses | Enter suggested reporting relevant to the results of additional analyses.<br><br>**Note:** Additional analyses can include sensitivity analysis, subgroup analysis, network meta-regression, individual participant data analyses. |
| Other | Enter any additional comments relevant to the results.<br><br>**Example:** if studies provide information on suggested reporting for the findings from checks performed to assess the appropriateness of NMA assumptions, please include relevant details in this section. |
| **Items Relevant to Discussion** | |
| Summary of Evidence | Enter suggested reporting relevant to the summary of evidence. |
| Limitations | Enter suggested reporting relevant to the limitations. |
| Conclusions | Enter suggested reporting relevant to the conclusions. |
| Other | Enter any additional comments relevant to the discussion. |
| **Items Relevant to Funding** | |
| Funding | Enter suggested reporting relevant to funding. |
| Other | Enter any additional comments relevant to the funding. |
| Author's Conclusions | Note down author's conclusions from the study, if relevant. |
| Author's Additional Suggestions for Consideration in Reporting | Note any additional suggestions from the authors. |
| NMA-Related Concepts/Terms with Definitions Reported | Enter any NMA-related concepts/terms that the study had defined along with the page number.<br><br>**Note:** Some NMA-related concepts/terms include: inconsistency/incoherence, transitivity/similarity/indirectness, Risk of bias/within-study bias, Publication bias/small-study effects/across-study |

| | |
|---|---|
| | bias |
| Additional Comments | Enter any relevant comments for the study that are not captured elsewhere here. |

**Sample checklist 1A: PRISMA NMA Checklist of Items to Include When Reporting A Systematic Review Involving a Network Meta-analysis[1]**

| Section/Topic | Item # | Checklist Item | Reported on Page # |
|---|---|---|---|
| **TITLE** | | | |
| Title | 1 | Identify the report as a systematic review *incorporating a network meta-analysis (or related form of meta-analysis).* | |
| **ABSTRACT** | | | |
| Structured summary | 2 | Provide a structured summary including, as applicable:<br>**Background:** main objectives<br>**Methods:** data sources; study eligibility criteria, participants, and interventions; study appraisal; and *synthesis methods, such as network meta-analysis.*<br>**Results:** number of studies and participants identified; summary estimates with corresponding confidence/credible intervals; *treatment rankings may also be discussed. Authors may choose to summarize pairwise comparisons against a chosen treatment included in their analyses for brevity.*<br>**Discussion/Conclusions:** limitations; conclusions and implications of findings.<br>**Other:** primary source of funding; systematic review registration number with registry name. | |
| **INTRODUCTION** | | | |
| Rationale | 3 | Describe the rationale for the review in the context of what is already known*, including mention of why a network meta-analysis has been conducted.* | |
| Objectives | 4 | Provide an explicit statement of questions being addressed, with reference to participants, interventions, comparisons, outcomes, and study design (PICOS). | |
| **METHODS** | | | |
| Protocol and registration | 5 | Indicate whether a review protocol exists and if and where it can be accessed (e.g., Web address); and, if available, provide registration information, including registration number. | |
| Eligibility criteria | 6 | Specify study characteristics (e.g., PICOS, length of follow-up) and report characteristics (e.g., years considered, language, publication status) used as criteria for eligibility, giving rationale. *Clearly describe eligible treatments included in the treatment network, and note whether any have been clustered or merged into the same node (with justification).* | |
| Information sources | 7 | Describe all information sources (e.g., databases with dates of coverage, contact with study authors to identify additional studies) | |

| | | |
|---|---|---|
| | | in the search and date last searched. |
| Search | 8 | Present full electronic search strategy for at least one database, including any limits used, such that it could be repeated. |
| Study selection | 9 | State the process for selecting studies (i.e., screening, eligibility, included in systematic review, and, if applicable, included in the meta-analysis). |
| Data collection process | 10 | Describe method of data extraction from reports (e.g., piloted forms, independently, in duplicate) and any processes for obtaining and confirming data from investigators. |
| Data items | 11 | List and define all variables for which data were sought (e.g., PICOS, funding sources) and any assumptions and simplifications made. |
| **Geometry of the network** | **S1** | Describe methods used to explore the geometry of the treatment network under study and potential biases related to it. This should include how the evidence base has been graphically summarized for presentation, and what characteristics were compiled and used to describe the evidence base to readers. |
| Risk of bias within individual studies | 12 | Describe methods used for assessing risk of bias of individual studies (including specification of whether this was done at the study or outcome level), and how this information is to be used in any data synthesis. |
| Summary measures | 13 | State the principal summary measures (e.g., risk ratio, difference in means). *Also describe the use of additional summary measures assessed, such as treatment rankings and surface under the cumulative ranking curve (SUCRA) values, as well as modified approaches used to present summary findings from meta-analyses.* |
| Planned methods of analysis | 14 | Describe the methods of handling data and combining results of studies for each network meta-analysis. This should include, but not be limited to:<br>• *Handling of multi-arm trials;*<br>• *Selection of variance structure;*<br>• *Selection of prior distributions in Bayesian analyses; and*<br>• *Assessment of model fit.* |
| **Assessment of Inconsistency** | **S2** | Describe the statistical methods used to evaluate the agreement of direct and indirect evidence in the treatment network(s) studied. Describe efforts taken to address its presence when found. |
| Risk of bias across studies | 15 | Specify any assessment of risk of bias that may affect the cumulative evidence (e.g., publication bias, selective reporting within studies). |
| Additional analyses | 16 | Describe methods of additional analyses if done, indicating which were pre-specified. This may include, but not be limited to, the following:<br>• Sensitivity or subgroup analyses;<br>• Meta-regression analyses;<br>• *Alternative formulations of the treatment network; and*<br>• *Use of alternative prior distributions for Bayesian analyses (if applicable).* |
| **RESULTS†** | | |
| Study selection | 17 | Give numbers of studies screened, assessed for eligibility, and included in the review, with reasons for exclusions at each stage, |

| | | |
|---|---|---|
| | | ideally with a flow diagram. |
| **Presentation of network structure** | **S3** | Provide a network graph of the included studies to enable visualization of the geometry of the treatment network. |
| **Summary of network geometry** | **S4** | Provide a brief overview of characteristics of the treatment network. This may include commentary on the abundance of trials and randomized patients for the different interventions and pairwise comparisons in the network, gaps of evidence in the treatment network, and potential biases reflected by the network structure. |
| Study characteristics | 18 | For each study, present characteristics for which data were extracted (e.g., study size, PICOS, follow-up period) and provide the citations. |
| Risk of bias within studies | 19 | Present data on risk of bias of each study and, if available, any outcome level assessment. |
| Results of individual studies | 20 | For all outcomes considered (benefits or harms), present, for each study: 1) simple summary data for each intervention group, and 2) effect estimates and confidence intervals. *Modified approaches may be needed to deal with information from larger networks.* |
| Synthesis of results | 21 | Present results of each meta-analysis done, including confidence/credible intervals. *In larger networks, authors may focus on comparisons versus a particular comparator (e.g. placebo or standard care), with full findings presented in an appendix. League tables and forest plots may be considered to summarize pairwise comparisons.* If additional summary measures were explored (such as treatment rankings), these should also be presented. |
| **Exploration for inconsistency** | **S5** | Describe results from investigations of inconsistency. This may include such information as measures of model fit to compare consistency and inconsistency models, *P* values from statistical tests, or summary of inconsistency estimates from different parts of the treatment network. |
| Risk of bias across studies | 22 | Present results of any assessment of risk of bias across studies for the evidence base being studied. |
| Results of additional analyses | 23 | Give results of additional analyses, if done (e.g., sensitivity or subgroup analyses, meta-regression analyses*, alternative network geometries studied, alternative choice of prior distributions for Bayesian analyses,* and so forth). |
| **DISCUSSION** | | |
| Summary of evidence | 24 | Summarize the main findings, including the strength of evidence for each main outcome; consider their relevance to key groups (e.g., healthcare providers, users, and policy-makers). |
| Limitations | 25 | Discuss limitations at study and outcome level (e.g., risk of bias), and at review level (e.g., incomplete retrieval of identified research, reporting bias). *Comment on the validity of the assumptions, such as transitivity and consistency. Comment on any concerns regarding network geometry (e.g., avoidance of certain comparisons).* |
| Conclusions | 26 | Provide a general interpretation of the results in the context of other evidence, and implications for future research. |
| **FUNDING** | | |

| | | |
|---|---|---|
| Funding | 27 | Describe sources of funding for the systematic review and other support (e.g., supply of data); role of funders for the systematic review. *This should also include information regarding whether funding has been received from manufacturers of treatments in the network and/or whether some of the authors are content experts with professional conflicts of interest that could affect use of treatments in the network.* |

**Abbreviation:** PICOS: population, intervention, comparators, outcomes, study design.
* Text in italics indicates wording specific to reporting of network meta-analyses that has been added to guidance from the PRISMA statement.
† Authors may wish to plan for use of appendices to present all relevant information in full detail for items in this section.

#### Sample checklist 1B: PRISMA 2020 Checklist[2]

| Section and Topic | Item # | Checklist item | Location where item is reported |
|---|---|---|---|
| **TITLE** | | | |
| Title | 1 | Identify the report as a systematic review. | |
| **ABSTRACT** | | | |
| Abstract | 2 | See the PRISMA 2020 for Abstracts checklist. | |
| **INTRODUCTION** | | | |
| Rationale | 3 | Describe the rationale for the review in the context of existing knowledge. | |
| Objectives | 4 | Provide an explicit statement of the objective(s) or question(s) the review addresses. | |
| **METHODS** | | | |
| Eligibility criteria | 5 | Specify the inclusion and exclusion criteria for the review and how studies were grouped for the syntheses. | |
| Information sources | 6 | Specify all databases, registers, websites, organisations, reference lists and other sources searched or consulted to identify studies. Specify the date when each source was last searched or consulted. | |
| Search strategy | 7 | Present the full search strategies for all databases, registers and websites, including any filters and limits used. | |
| Selection process | 8 | Specify the methods used to decide whether a study met the inclusion criteria of the review, including how many reviewers screened each record and each report retrieved, whether they worked independently, and if applicable, details of automation tools used in the process. | |
| Data collection process | 9 | Specify the methods used to collect data from reports, including how many reviewers collected data from each report, whether they worked independently, any processes for obtaining or confirming data from study investigators, and if applicable, details of automation tools used in the process. | |
| Data items | 10a | List and define all outcomes for which data were sought. Specify whether all results that were compatible with each outcome domain in each study were sought (e.g. for all measures, time points, analyses), and if not, the methods used to decide which results to collect. | |
| | 10b | List and define all other variables for which data were sought (e.g. participant and intervention characteristics, funding sources). Describe any assumptions made about any missing or unclear information. | |
| Study risk of bias assessment | 11 | Specify the methods used to assess risk of bias in the included studies, including details of the tool(s) used, how many reviewers assessed each study and whether they worked independently, and if applicable, details of automation tools used in the process. | |
| Effect measures | 12 | Specify for each outcome the effect measure(s) (e.g. risk ratio, mean difference) used in the synthesis or presentation of results. | |
| Synthesis | 13a | Describe the processes used to decide which studies were eligible for each synthesis (e.g. tabulating the study intervention characteristics and | |

| Section and Topic | Item # | Checklist item | Location where item is reported |
|---|---|---|---|
| methods | | comparing against the planned groups for each synthesis (item #5)). | |
| | 13b | Describe any methods required to prepare the data for presentation or synthesis, such as handling of missing summary statistics, or data conversions. | |
| | 13c | Describe any methods used to tabulate or visually display results of individual studies and syntheses. | |
| | 13d | Describe any methods used to synthesize results and provide a rationale for the choice(s). If meta-analysis was performed, describe the model(s), method(s) to identify the presence and extent of statistical heterogeneity, and software package(s) used. | |
| | 13e | Describe any methods used to explore possible causes of heterogeneity among study results (e.g. subgroup analysis, meta-regression). | |
| | 13f | Describe any sensitivity analyses conducted to assess robustness of the synthesized results. | |
| Reporting bias assessment | 14 | Describe any methods used to assess risk of bias due to missing results in a synthesis (arising from reporting biases). | |
| Certainty assessment | 15 | Describe any methods used to assess certainty (or confidence) in the body of evidence for an outcome. | |
| **RESULTS** | | | |
| Study selection | 16a | Describe the results of the search and selection process, from the number of records identified in the search to the number of studies included in the review, ideally using a flow diagram. | |
| | 16b | Cite studies that might appear to meet the inclusion criteria, but which were excluded, and explain why they were excluded. | |
| Study characteristics | 17 | Cite each included study and present its characteristics. | |
| Risk of bias in studies | 18 | Present assessments of risk of bias for each included study. | |
| Results of individual studies | 19 | For all outcomes, present, for each study: (a) summary statistics for each group (where appropriate) and (b) an effect estimate and its precision (e.g. confidence/credible interval), ideally using structured tables or plots. | |
| Results of syntheses | 20a | For each synthesis, briefly summarise the characteristics and risk of bias among contributing studies. | |
| | 20b | Present results of all statistical syntheses conducted. If meta-analysis was done, present for each the summary estimate and its precision (e.g. confidence/credible interval) and measures of statistical heterogeneity. If comparing groups, describe the direction of the effect. | |
| | 20c | Present results of all investigations of possible causes of heterogeneity among study results. | |
| | 20d | Present results of all sensitivity analyses conducted to assess the robustness of the synthesized results. | |
| Reporting biases | 21 | Present assessments of risk of bias due to missing results (arising from | |

| Section and Topic | Item # | Checklist item | Location where item is reported |
|---|---|---|---|
| | | reporting biases) for each synthesis assessed. | |
| Certainty of evidence | 22 | Present assessments of certainty (or confidence) in the body of evidence for each outcome assessed. | |
| **DISCUSSION** | | | |
| Discussion | 23a | Provide a general interpretation of the results in the context of other evidence. | |
| | 23b | Discuss any limitations of the evidence included in the review. | |
| | 23c | Discuss any limitations of the review processes used. | |
| | 23d | Discuss implications of the results for practice, policy, and future research. | |
| **OTHER INFORMATION** | | | |
| Registration and protocol | 24a | Provide registration information for the review, including register name and registration number, or state that the review was not registered. | |
| | 24b | Indicate where the review protocol can be accessed, or state that a protocol was not prepared. | |
| | 24c | Describe and explain any amendments to information provided at registration or in the protocol. | |
| Support | 25 | Describe sources of financial or non-financial support for the review, and the role of the funders or sponsors in the review. | |
| Competing interests | 26 | Declare any competing interests of review authors. | |
| Availability of data, code and other materials | 27 | Report which of the following are publicly available and where they can be found: template data collection forms; data extracted from included studies; data used for all analyses; analytic code; any other materials used in the review. | |

**References**

1. Hutton B, Salanti G, Caldwell DM, Chaimani A, Schmid CH, Cameron C, Ioannidis JP, Straus S, Thorlund K, Jansen JP, Mulrow C, Catalá-López F, Gøtzsche PC, Dickersin K, Boutron I, Altman DG, Moher D. The PRISMA Extension Statement for Reporting of Systematic Reviews Incorporating Network Meta-analyses of Health Care Interventions: Checklist and Explanations. Ann Intern Med. 2015;162(11):777-784. doi: 10.7326/M14-2385
2. Page MJ, Moher D, Bossuyt PM, Boutron I, Hoffmann TC, Mulrow CD, Shamseer L, Tetzlaff JM, Akl EA, Brennan SE, Chou R, Glanville J, Grimshaw JM, Hróbjartsson A, Lalu MM, Li T, Loder EW, Mayo-Wilson E, McDonald S, McGuinness LA, Stewart LA, Thomas J, Tricco AC, Welch VA, Whiting P, McKenzie JE. PRISMA 2020 explanation and elaboration: updated guidance and exemplars for reporting systematic reviews. BMJ. 2021 Mar 29;372:n160. doi: 10.1136/bmj.n160. PMID: 33781993; PMCID: PMC8005925.
3. Chaimani A, Caldwell DM, Li T, Higgins JPT, Salanti G. Chapter 11: Undertaking network meta-analyses. In: Higgins JPT, Thomas J, Chandler J, Cumpston M, Li T, Page MJ, Welch VA (editors). Cochrane Handbook for Systematic Reviews of Interventions version 6.4 (updated August 2023). Cochrane, 2023. Available from www.training.cochrane.org/handbook.

#### Appendix 8: List of 62 included studies and 10 companion reports

##### Unique studies (n = 62*)

*61 studies were included in the analysis. 1 study was not considered in the analysis as it was the original PRISMA-NMA checklist.

1. Donegan S, Williamson P, D'Alessandro U, Tudur Smith C. Assessing key assumptions of network meta-analysis: a review of methods. Res Synth Methods. 2013;4(4):291-323.
2. Tan SH, Bujkiewicz S, Sutton A, Dequen P, Cooper N. Presentational approaches used in the UK for reporting evidence synthesis using indirect and mixed treatment comparisons. J Health Serv Res Policy. 2013;18(4):224-32.
3. Bafeta A, Trinquart L, Seror R, Ravaud P. Reporting of results from network meta-analyses: methodological systematic review. BMJ. 2014;348:g1741.
4. Catala-Lopez F, Tobias A, Cameron C, Moher D, Hutton B. Network meta-analysis for comparing treatment effects of multiple interventions: an introduction. Rheumatol Int. 2014;34(11):1489-96.
5. Cope S, Zhang J, Saletan S, Smiechowski B, Jansen JP, Schmid P. A process for assessing the feasibility of a network meta-analysis: a case study of everolimus in combination with hormonal therapy versus chemotherapy for advanced breast cancer. BMC Med. 2014;12:93.
6. Jansen JP, Trikalinos T, Cappelleri JC, Daw J, Andes S, Eldessouki R, et al. Indirect treatment comparison/network meta-analysis study questionnaire to assess relevance and credibility to inform health care decision making: an ISPOR-AMCP-NPC Good Practice Task Force report. Value Health. 2014;17(2):157-73.
7. Laws A, Kendall R, Hawkins N. A comparison of national guidelines for network meta-analysis. Value Health. 2014;17(5):642-54.
8. Ortega A, Fraga MD, Alegre-del-Rey EJ, Puigventos-Latorre F, Porta A, Ventayol P, et al. A checklist for critical appraisal of indirect comparisons. Int J Clin Pract. 2014;68(10):1181-9.
9. Puhan MA, Schunemann HJ, Murad MH, Li T, Brignardello-Petersen R, Singh JA, et al. A GRADE Working Group approach for rating the quality of treatment effect estimates from network meta-analysis. BMJ. 2014;349:g5630.
10. Sullivan SM, Coyle D, Wells G. What guidance are researchers given on how to present network meta-analyses to end-users such as policymakers and clinicians? A systematic review. PLoS One. 2014;9(12):e113277.
11. Chambers JD, Naci H, Wouters OJ, Pyo J, Gunjal S, Kennedy IR, et al. An assessment of the methodological quality of published network meta-analyses: a systematic review. PLoS One. 2015;10(4):e0121715.
12. Foote CJ, Chaudhry H, Bhandari M, Thabane L, Furukawa TA, Petrisor B, et al. Network Meta-analysis: Users' Guide for Surgeons: Part I - Credibility. Clin Orthop Relat Res. 2015;473(7):2166-71.
13. Hutton B, Salanti G, Caldwell DM, Chaimani A, Schmid CH, Cameron C, et al. The PRISMA extension statement for reporting of systematic reviews incorporating network meta-analyses of health care interventions: checklist and explanations. Ann Intern Med. 2015;162(11):777-84.
14. Richter T, Lee KM. Guidance Document on Reporting Indirect Comparisons. In: CADTH, editor. 2015.
15. Fleetwood K, Glanville J, McCool R, Wood H, Wilson K, Marshall C, et al. A Review of the Use of Network Meta-Analysis In Nice Single Technology Appraisals. Value in Health. 2016;19(7):348.
16. Ge L, Tian JH, Li XX, Song F, Li L, Zhang J, et al. Epidemiology Characteristics, Methodological Assessment and Reporting of Statistical Analysis of Network Meta-Analyses in the Field of Cancer. Sci Rep. 2016;6:37208.
17. Veroniki AA, Straus SE, Soobiah C, Elliott MJ, Tricco AC. A scoping review of indirect comparison methods and applications using individual patient data. BMC Med Res Methodol. 2016;16:47.
18. Chaimani A, Caldwell DM, Li T, Higgins JPT, Salanti G. Additional considerations are required when preparing a protocol for a systematic review with multiple interventions. J Clin Epidemiol. 2017;83:65-74.
19. Chaimani A, Salanti G, Leucht S, Geddes JR, Cipriani A. Common pitfalls and mistakes in the set-up, analysis and interpretation of results in network meta-analysis: what clinicians should look for in a published article. Evid Based Ment Health. 2017;20(3):88-94.
20. Hummel N, Debray TPA, Didden EM, Efthimiou O, Egger M, Fletcher C, et al. Methodological guidance, recommendations and illustrative case studies for (network) meta-analysis and modelling to predict real-world effectiveness using individual participant and/or aggregate data. 2017.
21. Zarin W, Veroniki AA, Nincic V, Vafaei A, Reynen E, Motiwala SS, et al. Characteristics and knowledge synthesis approach for 456 network meta-analyses: a scoping review. BMC Med. 2017;15(1):3.

22. Al Khalifah R, Florez ID, Guyatt G, Thabane L. Network meta-analysis: users' guide for pediatricians. BMC Pediatr. 2018;18(1):180.
23. Brignardello-Petersen R, Bonner A, Alexander PE, Siemieniuk RA, Furukawa TA, Rochwerg B, et al. Advances in the GRADE approach to rate the certainty in estimates from a network meta-analysis. J Clin Epidemiol. 2018;93:36-44.
24. Dias S, Ades AE, Welton NJ, Jansen JP, Sutton AJ. Network Meta-Analysis for Decision Making2018.
25. James A, Yavchitz A, Ravaud P, Boutron I. Node-making process in network meta-analysis of nonpharmacological treatment are poorly reported. J Clin Epidemiol. 2018;97:95-102.
26. Lee DW, Shin IS. Critical quality evaluation of network meta-analyses in dental care. J Dent. 2018;75:7-11.
27. Morton SC, Murad MH, O'Connor E, Lee CS, Booth M, Vandermeer BW, et al. Quantitative Synthesis-An Update. Methods Guide for Effectiveness and Comparative Effectiveness Reviews. AHRQ Methods for Effective Health Care. Rockville (MD)2018.
28. Shi C, Westby M, Norman G, Dumville JC, Cullum N. Node-making processes in network meta-analysis of nonpharmacological interventions should be well planned and reported. J Clin Epidemiol. 2018;101:124-5.
29. Spineli LM, Yepes-Nunez JJ, Schunemann HJ. A systematic survey shows that reporting and handling of missing outcome data in networks of interventions is poor. BMC Med Res Methodol. 2018;18(1):115.
30. Tonin FS, Steimbach LM, Mendes AM, Borba HH, Pontarolo R, Fernandez-Llimos F. Mapping the characteristics of network meta-analyses on drug therapy: A systematic review. PLoS One. 2018;13(4):e0196644.
31. Tricco AC, Zarin W, Ghassemi M, Nincic V, Lillie E, Page MJ, et al. Same family, different species: methodological conduct and quality varies according to purpose for five types of knowledge synthesis. J Clin Epidemiol. 2018;96:133-42.
32. Williams T, Stein DJ, Ipser J. A systematic review of network meta-analyses for pharmacological treatment of common mental disorders. Evid Based Ment Health. 2018;21(1):7-11.
33. Yang F, Wang H, Zou J, Li X, Jin X, Cao Y, et al. Assessing the methodological and reporting quality of network meta-analyses in Chinese medicine. Medicine (Baltimore). 2018;97(47):e13052.
34. Chaimani A, Caldwell DM, Li T, Higgins JPT, Salanti G. Undertaking network meta-analyses. Cochrane Handbook for Systematic Reviews of Interventions. 2nd ed2019.
35. Chen Y, Zeng XY, Liu DF, Tan XY, Cai XM, Yang FW, et al. [Critical quality evaluation and application value of network Meta-analyses in traditional Chinese medicine]. Zhongguo Zhong Yao Za Zhi. 2019;44(24):5322-8.
36. Gao Y, Ge L, Ma X, Shen X, Liu M, Tian J. Improvement needed in the network geometry and inconsistency of Cochrane network meta-analyses: a cross-sectional survey. J Clin Epidemiol. 2019;113:214-27.
37. Phillippo DM, Dias S, Welton NJ, Caldwell DM, Taske N, Ades AE. Threshold Analysis as an Alternative to GRADE for Assessing Confidence in Guideline Recommendations Based on Network Meta-analyses. Ann Intern Med. 2019;170(8):538-46.
38. Tonin FS, Borba HH, Leonart LP, Mendes AM, Steimbach LM, Pontarolo R, et al. Methodological quality assessment of network meta-analysis of drug interventions: implications from a systematic review. Int J Epidemiol. 2019;48(2):620-32.
39. Brignardello-Petersen R, Florez ID, Izcovich A, Santesso N, Hazlewood G, Alhazanni W, et al. GRADE approach to drawing conclusions from a network meta-analysis using a minimally contextualised framework. BMJ. 2020;371:m3900.
40. Dwan K, Livingstone N. Editorial considerations in reviews with network meta-analysis 2020 [Available from: https://training.cochrane.org/resource/editorial-considerations-reviews-network-meta-analysis.
41. Gao Y, Shi S, Li M, Luo X, Liu M, Yang K, et al. Statistical analyses and quality of individual participant data network meta-analyses were suboptimal: a cross-sectional study. BMC Med. 2020;18(1):120.
42. Lee A. Developing critical appraisal of systematic reviews reporting network meta-analysis: University of Oxford; 2020.
43. Nikolakopoulou A, Higgins JPT, Papakonstantinou T, Chaimani A, Del Giovane C, Egger M, et al. CINeMA: An approach for assessing confidence in the results of a network meta-analysis. PLoS Med. 2020;17(4):e1003082.
44. Pratt M, Wieland S, Ahmadzai N, Butler C, Wolfe D, Pussagoda K, et al. A scoping review of network meta-analyses assessing the efficacy and safety of complementary and alternative medicine interventions. Syst Rev. 2020;9(1):97.
45. Welton NJ, Phillippo DM, Owen R, Jones HE, Dias S, Bujkiewicz S, et al. CHTE2020 SOURCES AND SYNTHESIS OF EVIDENCE; UPDATE TO EVIDENCE SYNTHESIS METHODS. 2020.

46. Bae K, Shin IS. Critical evaluation of reporting quality of network meta-analyses assessing the effectiveness of acupuncture. Complement Ther Clin Pract. 2021;45:101459.
47. Chiocchia V, Nikolakopoulou A, Higgins JPT, Page MJ, Papakonstantinou T, Cipriani A, et al. ROB-MEN: a tool to assess risk of bias due to missing evidence in network meta-analysis. BMC Med. 2021;19(1):304.
48. Veroniki AA, Tsokani S, Zevgiti S, Pagkalidou I, Kontouli KM, Ambarcioglu P, et al. Do reporting guidelines have an impact? Empirical assessment of changes in reporting before and after the PRISMA extension statement for network meta-analysis. Syst Rev. 2021;10(1):246.
49. Wright E, Yasmeen N, Malottki K, Sawyer LM, Borg E, Schwenke C, et al. Assessing the Quality and Coherence of Network Meta-Analyses of Biologics in Plaque Psoriasis: What Does All This Evidence Synthesis Tell Us? Dermatol Ther (Heidelb). 2021;11(1):181-220.
50. Yuan T, Xiong J, Wang X, Yang J, Jiang Y, Zhou X, et al. The Quality of Methodological and Reporting in Network Meta-Analysis of Acupuncture and Moxibustion: A Cross-Sectional Survey. Evid Based Complement Alternat Med. 2021;2021:2672173.
51. Cho SH, Shin IS. Evaluation of the Reporting Standard Guidelines of Network Meta-Analyses in Physical Therapy: A Systematic Review. Healthcare (Basel). 2022;10(12).
52. Wang R, Dwan K, Showell MG, van Wely M, Mol BW, Askie L, et al. Reporting of Cochrane systematic review protocols with network meta-analyses-A scoping review. Res Synth Methods. 2022;13(2):164-75.
53. de Sousa PG, Mainka FF, Tonin FS, Pontarolo R. Mapping the characteristics, methodological quality and standards of reporting of network meta-analyses on antithrombotic therapies: An overview. Int J Cardiol. 2023;386:125-33.
54. Montagner AF, Angst PDM, Raggio DP, FH VDS, Tedesco TK. Methodological quality of network meta-analysis in dentistry: a meta-research. Braz Oral Res. 2023;37:e062.
55. Nagendrababu V, Narasimhan S, Faggion CM, Jr., Dharmarajan L, Jacob PS, Gopinath VK, et al. Reporting quality of systematic reviews with network meta-analyses in Endodontics. Clin Oral Investig. 2023;27(7):3437-45.
56. Nast A, Dressler C, Schuster C, Saure D, Augustin M, Reich K. Methods used for indirect comparisons of systemic treatments for psoriasis. A systematic review. Skin Health Dis. 2023;3(1):e112.
57. Sehmbi H, Retter S, Shah UJ, Nguyen D, Martin J, Uppal V. Methodological and reporting quality assessment of network meta-analyses in anesthesiology: a systematic review and meta-epidemiological study. Can J Anaesth. 2023;70(9):1461-73.
58. Wang Y, Chen N, Guo K, Li Y, E F, Yang C, et al. Reporting and methodological quality of acupuncture network meta-analyses could be improved: an evidence mapping. J Clin Epidemiol. 2023;153:1-12.
59. Lovsletten PO, Wang X, Pitre T, Odegaard M, Veroniki AA, Lunny C, et al. A systematic survey of 200 systematic reviews with network meta-analysis (published 2020-2021) reveals that few reviews report structured evidence summaries. J Clin Epidemiol. 2024;173:111445.
60. Lunny C, Veroniki AA, Higgins JPT, Dias S, Hutton B, Wright JM, et al. Methodological review of NMA bias concepts provides groundwork for the development of a list of concepts for potential inclusion in a new risk of bias tool for network meta-analysis (RoB NMA Tool). Syst Rev. 2024;13(1):25.
61. Safety D-GfHaF. Practical Guideline for Quantitative Evidence Synthesis: Direct and Indirect Comparisons. In: Commission E, editor. 2024.
62. Spineli LM. An empirical study on 209 networks of treatments revealed intransitivity to be common and multiple statistical tests suboptimal to assess transitivity. BMC Med Res Methodol. 2024;24(1):301.

##### Companion reports (n = 10)

1. Hutton B, Salanti G, Chaimani A, Caldwell DM, Schmid C, Thorlund K, et al. The quality of reporting methods and results in network meta-analyses: an overview of reviews and suggestions for improvement. PLoS One. 2014;9(3):e92508.
2. Laws A, Kendall R, Hawkins N. A comparison of national guidelines for network meta-analysis. Value Health. 2014;17(5):642-54.
3. Salanti G, Del Giovane C, Chaimani A, Caldwell DM, Higgins JP. Evaluating the quality of evidence from a network meta-analysis. PLoS One. 2014;9(7):e99682.
4. Chambers JD, Naci H, Wouters OJ, Pyo J, Gunjal S, Kennedy IR, et al. Correction: An Assessment of the Methodological Quality of Published Network Meta-Analyses: A Systematic Review. PLoS One. 2015;10(7):e0131953.

5. Hutton B, Catala-Lopez F, Moher D. [The PRISMA statement extension for systematic reviews incorporating network meta-analysis: PRISMA-NMA]. Med Clin (Barc). 2016;147(6):262-6.
6. Brignardello-Petersen R, Izcovich A, Rochwerg B, Florez ID, Hazlewood G, Alhazanni W, et al. GRADE approach to drawing conclusions from a network meta-analysis using a partially contextualised framework. BMJ. 2020;371:m3907.
7. Papakonstantinou T, Nikolakopoulou A, Higgins JPT, Egger M, Salanti G. CINeMA: Software for semiautomated assessment of the confidence in the results of network meta-analysis. Campbell Syst Rev. 2020;16(1):e1080.
8. Nagendrababu V, Faggion CM, Jr., Pulikkotil SJ, Alatta A, Dummer PMH. Methodological assessment and overall confidence in the results of systematic reviews with network meta-analyses in Endodontics. Int Endod J. 2022;55(5):393-404.
9. Sehmbi H, Retter S, Shah UJ, Nguyen D, Martin J, Uppal V. Epidemiological, methodological, and statistical characteristics of network meta-analysis in anaesthesia: a systematic review. Br J Anaesth. 2023;130(3):272-86.
10. Veroniki AA, Wong EKC, Lunny C, Martinez Molina JC, Florez ID, Tricco AC, et al. Does type of funding affect reporting in network meta-analysis? A scoping review of network meta-analyses. Syst Rev. 2023;12(1):81.

#### Appendix 9: Full Table 2: Summary of included studies for the overviews of reviews with NMA (n=38)

**Table 2.** Summary of included studies for the overviews of reviews with NMA (n=38)

| First author, year | Summary of the objectives | Inclusion Criteria | # of NMAs included in the review | Timeframe of assessed literature | Conflicts of interest | Reporting/ methodological guideline used for appraisal (if any) | Synopsis of authors' conclusions |
|---|---|---|---|---|---|---|---|
| Donegan, 2013[4] | Review and illustrate methods to assess homogeneity and consistency. | -Used at least one of the following terms within the title or abstract 'multiple treatment(s) metaanalysis', 'NMA', 'indirect evidence', 'indirect comparison', 'mixed treatment comparison' or 'multiple treatment comparison' in terms of meta-analysis.<br>-Published articles or unpublished, externally peer reviewed articles.<br>-Written in English.<br>-Focussed on indirect comparisons or NMAs. | 116 | NR | NR | NR | Homogeneity and consistency assumptions can be assessed using established methods and expert collaboration. Routine application of these methods ensures appropriate interpretation of NMA results. |
| Tan, 2013[5] | Review guidance on the presentation of IC/MTC analyses for institutional guidelines and to assess what has previously been done in practice in UK by reviewing HTA reports. Second, to provide recommendations on how to improve future reporting of IC/MTC analyses. And, third, to identify research priorities for improving the presentation of IC/MTC analyses. | Reports published in UK by the Health Technology Assessment (HTA) programme, which utilized indirect comparison/mixed treatment comparison methods. | 19 | 1997 – 2011 (full date: NR) | NR | NR | Consistent and clear result presentation enhances interpretability, clarifies treatment relationships, and supports sensitivity, subgroup, and inconsistency analyses. |
| Bafeta, 2014[6] | Examine how network meta-analysis results are reported; To assess reporting of the amount | All network meta-analyses comparing the clinical efficacy of three or more interventions in randomised controlled trials. | 121 | Inception - July 12, 2012 | No | NR | Network meta-analyses are not consistently reported, reflecting a lack of agreement on reporting standards and the |

| First author, year | Summary of the objectives | Inclusion Criteria | # of NMAs included in the review | Timeframe of assessed literature | Conflicts of interest | Reporting/ methodological guideline used for appraisal (if any) | Synopsis of authors' conclusions |
|---|---|---|---|---|---|---|---|
| | of evidence in the network of trials or the reporting of effect estimates with uncertainty from direct, indirect, or mixed evidence. | | | | | | need for guidelines. Key aspects like network geometry, trial numbers, patient data, and grouping of similar interventions must be clearly reported to ensure reliable interpretation and assess evidence quality. |
| Laws, 2014[7] | Examine both published and draft guidelines from reimbursement and health technology appraisal bodies, and considered their recommendations using appropriate methodology for the conduct of indirect comparisons and the assessments of their validity. | NR | NR | Inception - July 22, 2013 | No | ISPOR | No two recommendations from the multiple national guidelines are mutually exclusive. It is possible to perform one network meta-analysis for submission to multiple national jurisdictions. |
| Ortega, 2014[8] | Develop a user-friendly checklist for evaluating indirect comparisons to determine whether the results of an IC can be used to facilitate the decision-making process of drug evaluation and selection in clinical practice. | All studies that include indirect comparisons of treatments, divided into on the one hand, articles on methodology, including methods that allow for comparing treatments when direct comparisons and ECAs and RCTs are not available, methods for identifying possible bias and methodological problems with indirect comparisons, advantages/disadvantages of indirect vs. direct comparisons, and on the other hand, RCTs, meta-analyses, and RS that include indirect comparisons of drugs or a combination of | 60 | Inception - October, 2011 (full date: NR) | NR | NR | A user-friendly checklist for critically appraising indirect comparisons (ICs) was developed to assist in drug evaluation and selection. Focused on early drug assessment with limited RCTs, it includes fewer NMA-specific items, as some aspects overlap with standard meta-analyses and other IC types. |

| First author, year | Summary of the objectives | Inclusion Criteria | # of NMAs included in the review | Timeframe of assessed literature | Conflicts of interest | Reporting/ methodological guideline used for appraisal (if any) | Synopsis of authors' conclusions |
|---|---|---|---|---|---|---|---|
| | | indirect and direct comparisons, Articles in English, Spanish, or French. | | | | | |
| Sullivan, 2014[9] | (1) Determine if researchers are provided any guidance on how to present a NMA (2) determine if this guidance is targeted toward non-technical end-users such as policymakers or clinicians and (3) interpret these findings in the context of non-technical end-users' needs, who must apply the results of NMAs to policy or practice decisions. | -Guidelines for conducting or reporting on NMAs that primarily targeted statisticians, researchers and others who produce NMAs (e.g., indirect comparisons, mixed treatment comparisons, multiple treatment comparisons) for the purpose of informing healthcare policy and practice decisions.<br>-Formal NMA guidelines as well as interim guidance documents or working group documents that provided recommendations and were developed by collaborative groups or organizations with the intent of informing formal NMA guidelines. | 7 | 1996 - June 2014; 1980 - June 2014. (full date: NR) | No | National/international guidelines (e.g., ISPOR, CDA, NICE, PBAC, HAS, AHRQ and EUnetHTA, AGREE-II) | There is limited guidance available on presenting NMAs in accessible formats for non-technical users, such as policymakers and clinicians. Existing NMA guidelines focus on conducting analyses but lack reporting templates, tailored presentations, or extensive glossaries for diverse end-users. Improving presentation formats and integrating end-user needs can enhance the transparency, accessibility, and legitimacy of NMA-informed decisions. |
| Chambers, 2015[10] | Assess the methodological quality of published network meta-analysis. | -All published network meta-analyses in the Ovid-MEDLINE database using the following search terms: network meta-analysis; indirect treatment comparison; mixed treatment comparison; and, multiple treatments meta-analysis.<br>-Studies including randomized controlled trials with human participants.<br>-Articles published in English-language journals. | 318 | Inception - July 30, 2014 | No | ISPOR | Higher-impact journals publish NMAs with higher quality, and recent studies show negligible improvements in risk of bias assessments. To enhance NMA quality and consistency, consensus among guidelines is needed, along with efforts by journal editors to ensure transparent, reproducible, and comprehensive reporting. |
| Fleetwood, 2016[11] | Evaluate the use of NMA within Single Technology Appraisals (STAs) with respect to | NICE STAs | 25 | May 1, 2015 - April 30, 2016 | NR | NICE | STAs often include NMAs but may not fully align with NICE guidelines. Manufacturers should follow NICE Decision |

| First author, year | Summary of the objectives | Inclusion Criteria | # of NMAs included in the review | Timeframe of assessed literature | Conflicts of interest | Reporting/ methodological guideline used for appraisal (if any) | Synopsis of authors' conclusions |
|---|---|---|---|---|---|---|---|
| | the National Institute for Health and Care Excellence (NICE) guidance. | | | | | | Support Unit (DSU) Technical Support Document (TSD) 7, justify key assumptions, and ensure methods are transparent and reproducible. |
| Ge, 2016[12] | Conduct a methodological review of published NMAs in the field of cancer, summarise their characteristics, methodological quality, and reporting of key statistical analysis process. Compare the methodological quality and reporting of statistical analysis by selected general characteristics. | NMAs in the field of cancer in the English and Chinese languages, regardless of interventions. | 102 | Inception - July 9, 2015 | No | AMSTAR & PRISMA-NMA | Methodological quality of NMAs within cancer field is generally acceptable but shows flaws in literature search, quality assessment, combining findings, and addressing publication bias. Challenges like defining interventions, handling multi-group trials, and assessing similarity and consistency highlight the need for guidelines on reporting NMA statistical analyses. |
| Veroniki, 2016[13] | Conduct a comprehensive scoping review of the methods used to perform indirect comparisons with IPD or IPD combined with aggregated data. To review applications of indirect comparisons with IPD and summarize network, methods and reporting characteristics. | -Published papers, protocols, and abstracts, as well as unpublished studies that reported on a method, application, or review of IPD indirect comparison methods involving studies of any design.<br>-Application studies that compared the clinical effectiveness or safety of three or more interventions and applied any type of indirect comparison, including adjusted indirect comparison, unadjusted indirect comparison, matching adjusted indirect comparison (MAIC), simulated treatment comparison (STC), mixed comparison, and NMA. | 37 | Inception - October, 2014 (full date: NR) | Yes | PRISMA-NMA; PRISMA-IPD; ISPOR | Key methodological elements, like consistency evaluation and study protocols, are often missing in IPD-NMAs, even in high-impact journals, risking erroneous clinical decisions. Clear reporting aligned with ISPOR, PRISMA-IPD, and PRISMA-NMA guidelines is essential to improve transparency and reliability. |

| First author, year | Summary of the objectives | Inclusion Criteria | # of NMAs included in the review | Timeframe of assessed literature | Conflicts of interest | Reporting/ methodological guideline used for appraisal (if any) | Synopsis of authors' conclusions |
|---|---|---|---|---|---|---|---|
| Zarin, 2017[14] | Explore the characteristics and methodological quality of knowledge synthesis approaches underlying the NMA process. To assess the statistical methods applied using the Analysis subdomain of the ISPOR checklist. | -NMAs that compared at least four different interventions from RCTs using a valid statistical method for indirect comparisons (e.g., adjusted or anchored indirect comparison method) or NMAs (e.g., hierarchical models).<br>-Both published and unpublished reports in all languages of publication. | 456 | Inception - April 14, 2015 | Yes | ISPOR & AMSTAR | To make NMAs valuable for healthcare decision-makers, improving their reporting and conduct is essential for transparency, reproducibility, and quality. Education for researchers, training for editors and peer reviewers, and adopting guidelines like PRISMA-NMA can enhance methodological rigor and reporting completeness. |
| James, 2018[15] | Identify methods for building nodes in NMAs assessing nonpharmacological interventions. | -All published reports of NMAs assessing nonpharmacological treatments in humans.<br>-All reports of NMAs and indirect comparisons including at least three nodes and assessing at least one nonpharmacological treatment.<br>-No language, publication status, or date restrictions. | 116 | Inception – March, 2016 (full date: NR) | No | PRISMA-NMA | Despite the importance of NMAs, guidance on reporting the node-making process is limited. Authors should ensure transparency by understanding intervention content, clearly explaining nodes, and justifying the node-making process for replication. |
| Lee, 2018[16] | Examine whether the reports (SRs with NMA in dental care) adequately followed the key reporting components of PRISMA NMA. | All NMAs published in dental journals that compared the clinical efficacy of three or more interventions based on randomized controlled trials (RCT) or non-RCTs. | 21 | Inception - May 1, 2017 | No | PRISMA-NMA | The assessment revealed low-quality reporting and conduct of current NMAs, similar to other medical fields. Key issues include network geometry, inconsistency assessment, risk of bias, protocol registration, and additional analyses. |
| Spineli, 2018[17] | Bridge this knowledge gap by providing evidence about the impact of MOD on the credibility of a systematic review with NMA, including the reporting and handling of MOD as well as the | -Considered whether RCTs with 'non-standard' design, such as quasi, crossover, factorial, cluster, split-mouth, contralateral and split-face/body RCTs, were among the eligible trials<br>-Selected only one primary outcome From each eligible systematic review. | 387 | 2012-2017 (full date: NR) | No | NR | The reporting and handling of missing outcome data (MOD) in NMAs are inadequate, reflecting reviewers' limited awareness and knowledge of relevant methodologies. Improved methods, reporting guidelines, and education for reviewers, peer reviewers, and |

| First author, year | Summary of the objectives | Inclusion Criteria | # of NMAs included in the review | Timeframe of assessed literature | Conflicts of interest | Reporting/ methodological guideline used for appraisal (if any) | Synopsis of authors' conclusions |
|---|---|---|---|---|---|---|---|
| | acknowledgment of their implications. | -Gave priority to binary outcomes when the authors described more than one primary outcomes.<br>-Selected a patient-important outcome in the presence of many binary primary outcomes. | | | | | journal editors are essential to enhance transparency and the quality of healthcare inferences. |
| Tonin, 2018a[18] | Map the characteristics of the published NMAs on drug therapy comparisons. | -Studies using NMAs to compare any drug therapy intervention (defined as a pharmacological intervention including an active substance) alone or in combination with other pharmacological intervention, regardless of regimen or dosage.<br>-Any type of network (with open or closed loops) of experimental, quasi-experimental, or observational trials that assessed at least three or more treatments, comparing head to head or against placebo/no control in patients (no restriction of gender, age, or clinical/medical condition). | 365 | Inception – March, 2016 (full date: NR) | No | PRISMA-NMA | The use of online supplementary material in NMAs has increased and should include systematic review data and, if possible, raw data sets. Guidelines like PRISMA-NMA (2015) extension can improve reporting standards, enhancing evidence quality and reproducibility. |
| Tonin, 2018b[19] | Evaluate the methodological quality (R-AMSTAR) and the compliance with reporting guidelines (PRISMA statements) of the existing NMAs of drug interventions published until December 2016, and identify time trends and associations with the journal impact factor | -NMAs that had evaluated pharmacological interventions in any regimen or dosage.<br>-Any type of network of experimental, quasi-experimental or observational trials that assessed at least three or more treatments, compared head-to-head or against the placebo or controls in patients (with no restriction on gender or age) in any clinical condition. | 477 | Inception - April 25, 2017 | No | PRISMA, PRISMA-NMA, & R-AMSTAR | PRISMA-NMA is widely used, but compliance remains low with weak ties to journal impact factors. Enforcing guidelines, mandating raw data sharing, and using tools like GRADE can improve NMA transparency and quality. |

| First author, year | Summary of the objectives | Inclusion Criteria | # of NMAs included in the review | Timeframe of assessed literature | Conflicts of interest | Reporting/ methodological guideline used for appraisal (if any) | Synopsis of authors' conclusions |
|---|---|---|---|---|---|---|---|
| | and the country or countries of authors' affiliations. | | | | | | |
| Tricco, 2018[20] | Characterize methodological conduct, reporting, and quality of five knowledge synthesis (KS) approaches. | -NMAs of randomized controlled trials with a valid statistical method for comparisons (e.g., adjusted indirect comparisons).<br>-All pharmacological and non-pharmacological interventions.<br>-At least four different treatments or interventions being compared (including placebo or a no treatment group).<br>-Quality and reporting characteristics (outcome).<br>-English and other languages. | 456 | Inception – April, 2015 (full date: NR) | Yes | AMSTAR | NMAs consistently scored the highest on the AMSTAR tool likely because the purpose is to estimate treatment effects statistically. |
| Williams, 2018[21] | (A) Conduct a systematic review of published NMAs that have assessed the efficacy of pharmacological treatment for common mental disorders. (B) Review the quality of the methods of the published NMAs using a quality rating approach that has been designed specifically for NMAs. (C) Discuss differences in rankings of the efficacy and tolerability of pharmacological treatment in terms of methodological and clinical characteristics | -NMAs including RCTs of pharmacotherapy in treating adult participants (18–65 years) with common mental disorders diagnosed according to the Diagnostic Statistical Manual for Mental Disorders (DSM-III and later) or the International Classification of Diseases (ICD-10).<br>-NMAs containing RCTs that included participants with comorbid secondary mental disorders. | 20 | 2008 - March 22, 2017 | Yes | ISPOR | Most NMAs underutilized reporting tools like inconsistency or ranking plots and rarely provided full matrices of effect estimates or treatment rankings. This lack of visuals and rankings highlights critiques that NMAs are complex and primarily suited for researchers with advanced statistical skills. |

| First author, year | Summary of the objectives | Inclusion Criteria | # of NMAs included in the review | Timeframe of assessed literature | Conflicts of interest | Reporting/ methodological guideline used for appraisal (if any) | Synopsis of authors' conclusions |
|---|---|---|---|---|---|---|---|
| | of the NMAs by disorder. | | | | | | |
| Yang, 2018[22] | Assess the methodological and reporting quality of NMAs in Traditional Chinese Medicine. | -NMAs based on RCTs with the treatments of Traditional Chinese Medicine (TCM), which included Chinese herbal medicine and patent medicine.<br>-Other treatments like Western medicine with at least 1 TCM treatment in each NMA. | 40 | Inception – January, 2018 (full date: NR) | No | PRISMA-NMA & AMSTAR | The methodological and reporting quality of NMAs was moderate, with flaws in areas like publication status, study lists, bias assessment, protocols, conflicts of interest, and funding. To improve reporting, authors should follow the PRISMA-NMA checklist. |
| Chen, 2019[23] | To review the systematic reviews applying NMA in traditional Chinese medicine | -Interventions: any used in TCM but not restricted to medications via different routes and acupuncture; or related treatment complications and diseases<br>-Systematic reviews using NMA<br>-Languages restricted to English and Chinese | 122 | 2012-2019 | NR | AMSTAR; PRISMA-NMA | The assessment by methodological quality was higher after 2015 than in and before 2015, which was statistically significant. |
| Gao, 2019[24] | Investigate the general characteristics, methodological quality, and reporting quality of NMAs published in the Cochrane library. | NMAs published in the Cochrane library - Both the NMAs of randomized controlled trials and NMAs with observational studies. | 42 | Inception - April 3, 2018 | No | AMSTAR 2 & PRISMA-NMA | The quality of NMAs in the Cochrane Library, although has improved over time, still requires enhancements, particularly in publication bias assessment, network geometry, structure presentation, and inconsistency exploration. Addressing these deficiencies should be a focus for future work. |
| Gao, 2020[25] | Explore the general characteristics, statistical analysis methods used, reporting quality, and methodological quality of IPD-NMAs and to identify study-level | -IPD-NMAs of RCTs that evaluated the clinical effects of three or more interventions for patients in any clinical conditions.<br>-There were no restrictions on publication year and language. | 21 | Inception - June 3, 2019 | No | PRISMA-IPD & PRISMA-NMA | Published IPD-NMAs often lack key statistical details and have suboptimal compliance with reporting standards, particularly in handling missing data, publication bias, IPD integrity, network geometry, and consistency. Developing a |

| First author, year | Summary of the objectives | Inclusion Criteria | # of NMAs included in the review | Timeframe of assessed literature | Conflicts of interest | Reporting/ methodological guideline used for appraisal (if any) | Synopsis of authors' conclusions |
|---|---|---|---|---|---|---|---|
| | variables that were associated with the methodological and reporting quality. | | | | | | dedicated reporting tool combining PRISMA-IPD and PRISMA-NMA elements is essential to improve IPD-NMA quality. |
| Lee, 2020[26] | Construct and validate a critical appraisal tool for systematic reviews using NMA methodology for synthesis. | -Systematic reviews of healthcare interventions in humans, in which a meta-analysis combined direct and indirect comparisons among more than two interventions.<br>-Reviews of any interventions that report any outcome of benefit or harm.<br>-Conference abstracts if the comparisons and outcomes of meta-analysis were reported and the review had not been separately reported as a journal article.<br>-Methodological articles and cost effectiveness studies if they reported a new NMA using trials identified by a systematic review. | 201 | Inception - September 21, 2012 | No | PRISMA-NMA, NICE | A new critical appraisal tool was constructed for appraising systematic reviews using NMA methods for synthesis; The reporting of NMAs was inconsistent. The specific methodology used, and the assumptions made were often unclear or not stated in the published reports. |
| Pratt, 2020[27] | Establish the extent of published NMAs involving complementary and alternative medicine (CAM) interventions in the literature, to assess their objectives as well as clinical and methodologic characteristics, and to judge the current level of reporting transparency based | Systematic reviews incorporating NMAs involving one or more CAM interventions that (a) have used a valid comparison method (such as adjusted indirect comparison, Bayesian model, meta-regression, multivariate meta-analysis, graph theoretical approach); (b) included a minimum of 4 interventions in the network of evidence studied; (c) included a greater number of studies than there were nodes in | 90 | 2010 - May 29, 2018 | Yes | PRISMA-NMA | Both PRISMA and PRISMA-NMA has shortcomings, with methodological challenges in NMA assumptions and transitivity for CAM trials. As NMA use for CAM interventions grows, identifying priority topics and improving methods for comparing CAM with conventional medicine is essential. |

| First author, year | Summary of the objectives | Inclusion Criteria | # of NMAs included in the review | Timeframe of assessed literature | Conflicts of interest | Reporting/ methodological guideline used for appraisal (if any) | Synopsis of authors' conclusions |
|---|---|---|---|---|---|---|---|
| | upon criteria of the PRISMA NMA. | the network; and (d) included data from RCTs only. | | | | | |
| Bae, 2021[28] | Assess the completeness of reporting for published acupuncture-NMAs based on the Preferred Reporting Items for Systematic Reviews and Meta-analyses (PRISMA) Extension Statement for Reporting of Systematic Reviews Incorporating Network Meta-analyses of Health Care Interventions (PRISMA-NMA). | -Reports of NMAs evaluating the clinical efficacy of acupuncture. When an exact NMA terminology was not used, studies that used the analytical method to combine direct and indirect comparisons between multiple treatments.<br>-Studies involving human subjects with any medical conditions and no age limitations.<br>-Studies involving any type of acupuncture as treatment nodes.<br>-Studies that aimed to assess the clinical efficacy with any outcome measurements. | 42 | Inception - April 26, 2020 | No | PRISMA & PRISMA-NMA | The reporting quality of 42 acupuncture NMAs was low, particularly for new checklist items. Future NMAs should explicitly address network structure, transitivity, and consistency, with journals endorsing PRISMA-NMA guidelines to improve adherence and validity. |
| Veroniki, 2021[29] | Empirically assess whether the PRISMA-NMA statement had an important impact on the completeness of reporting by comparing NMA articles of randomised controlled trials (RCTs) between two time periods (2013–2015 and 2016–2018).<br>To investigate publication features and network characteristics that might modify the completeness of reporting.<br>To highlight key items that require further | -NMAs that compared at least four interventions including placebo, no treatment, waiting list or other control interventions.<br>-NMAs that conducted a valid statistical method for indirect comparisons (e.g., adjusted indirect comparison method) or NMAs (e.g., hierarchical models).<br>-Published NMAs written in English. | 1144 | Inception - July, 2018 (full date: NR) | No | PRISMA NMA | Expanded the PRISMA-NMA checklist from 32 to 49 items to optimally evaluate reporting practices, highlighting underreported aspects despite slight yearly improvements since 2015. To enhance reporting quality and minimize biases, particularly in industry-sponsored NMAs, the updated checklist should be widely adopted by authors, reviewers, editors, and journals. |

| First author, year | Summary of the objectives | Inclusion Criteria | # of NMAs included in the review | Timeframe of assessed literature | Conflicts of interest | Reporting/ methodological guideline used for appraisal (if any) | Synopsis of authors' conclusions |
|---|---|---|---|---|---|---|---|
| | attention and potential improvement moving forward. | | | | | | |
| Wang, 2021[30] | Evaluate how Cochrane NMA protocols were reported in 2018 and 2019, and to compare the protocol reporting across the 2 years to identify potential improvement over time. | -All protocols of systematic reviews of interventions with NMA published in the Cochrane Library between 2018 and 2019.<br>-Both protocols of systematic reviews of interventions and protocols of overviews of interventions. | 45 | January 1, 2018 - December 31, 2019 | NR | NR | Cochrane NMA protocols require improvement, particularly in addressing transitivity assumptions. Providing detailed, up-to-date methodological information and enhancing inclusion criteria, search strategies, data extraction, and statistical analysis are required for reproducibility and quality. |
| Wright, 2021[31] | Assess the methodological quality of these analyses and explore the differences in their methods and results. | -All NMAs assessing biologic treatments for moderate-to severe plaque psoriasis.<br>-NMAs that evaluated two or more currently licensed biologic treatments in patients with moderate-to-severe psoriasis.<br>-Analyses evaluating any efficacy or safety outcome.<br>-NMAs using aggregate level data and a Bayesian or frequentist approach. | 25 | Inception - February 19, 2020 | Yes | ISPOR | Overall, the NMAs reviewed met at least half of the ISPOR checklist criteria, showing moderate to good methodological quality. Robust NMAs should include all relevant trials and comparators, assess heterogeneity and inconsistency, and report comparative effects with uncertainty, ensuring relevance by reflecting the applicability and considering outcomes like efficacy, quality of life, and safety. |
| Yuan, 2021[32] | Evaluate the methodological and reporting quality of NMAs and summarize the effects of different treatments of acupuncture and moxibustion. | -All relevant NMAs treated with acupuncture and moxibustion.<br>-Other therapies such as traditional Chinese medicine and Western medicine may also be included, but at least three types fall under the category of acupuncture in each NMA. | 29 | Inception - January 22, 2020 | No | NR | Considering the importance of PRISMA-NMA checklist to NMA, it is advised that the researchers should strictly follow the PRISMA-NMA checklist when writing a NMA. |

| First author, year | Summary of the objectives | Inclusion Criteria | # of NMAs included in the review | Timeframe of assessed literature | Conflicts of interest | Reporting/ methodological guideline used for appraisal (if any) | Synopsis of authors' conclusions |
|---|---|---|---|---|---|---|---|
| | | -Outcome indicators were not limited, while language limited in Chinese and English. | | | | | |
| Cho, 2022[33] | Examine whether the reports adequately followed the key reporting components of the systematic review process, based on the preferred reporting items for systematic reviews and meta-analysis (PRISMA)-NMA Extension guidelines. | -All NMAs published in physical therapy journals comparing the clinical efficacy of three or more interventions based on RCTs.<br>-NMAs with at least one physical therapy in the set of treatments examined and the data from at least three clinical trials.<br>-NMAs that compared the efficacy of the treatments. | 19 | Inception - June 30, 2022 | NR | NR | The evaluation of NMA reporting standards and practices is low to moderate, with key issues in network geometry, inconsistency assessment, risk of bias, and protocol registration. Adopting and endorsing PRISMA-NMA guidelines can improve reporting and reduce the risk of invalid results in physical therapy. |
| Nast, 2022[34] | Determine whether we could develop guidance to help the clinical community better understand indirect comparison and NMA methodologies in order to assist them with interpreting the results of these types of analyses. | -Study design: Adjusted indirect comparison, matching-adjusted indirect comparison, network metaanalysis.<br>-Patient population: Adult patients with moderate to severe psoriasis.<br>-Intervention: Any biologic treatment for psoriasis.<br>-Comparator: Placebo or any systemic treatment for psoriasis.<br>-Outcomes: Efficacy, safety<br>-Publication type: Journal article<br>-Language: English | 22 | 2012 - March 26, 2020 | Yes | NICE DSU, 2017 AMSTAR 2 | Highlights the need to improve methods and reporting in SLRs and NMAs for psoriasis, as only 2 of 26 indirect comparisons were of 'high' quality, with most rated 'low' or 'critically low.' Key learnings include differences in study characteristics, misleading presentation of results, methodological inadequacies, and the need for quality improvement using tools like AMSTAR 2, NICE TSD7, and PRISMA-NMA. |
| Wang, 2022[35] | Assess and compare the reporting and methodological quality of Chinese and English NMAs on acupuncture, providing a reference for the development of high-quality | -All relevant NMAs on acupuncture.<br>-NMA on both acupressure and laser acupuncture. | 113 | Inception – February, 2022 (full date: NR) | No | NR | NMAs within acupuncture have increased, with significant quality variations between studies in Chinese and English. To improve quality, research should follow PICOS guidelines, present network structures, and use tools like PRISMA-NMA and |

| First author, year | Summary of the objectives | Inclusion Criteria | # of NMAs included in the review | Timeframe of assessed literature | Conflicts of interest | Reporting/ methodological guideline used for appraisal (if any) | Synopsis of authors' conclusions |
|---|---|---|---|---|---|---|---|
| | acupuncture studies in future. | | | | | | AMSTAR-2, while developing specific assessment tools and training authors and reviewers on internationally recognized standards. |
| de Sousa, 2023[36] | Map the characteristics and critically appraised the standards of conduct and evidence reporting of NMAs assessing antithrombotic therapies for the treatment or prophylaxis of heart diseases and cardiac surgical procedures. | -NMAs comparing any antithrombotic therapy for the treatment or prophylaxis of heart diseases or during cardiac surgical procedures.<br>-Any type of network of experimental, quasi-experimental or observational trials that assessed at least three or more treatments, comparing head-to-head or against placebo/no control in adult patients.<br>-Studies with reported data on interventions' efficacy or safety outcomes. | 86 | Inception - March, 2022 (full date: NR) | No | NR | Journal editors, peer reviewers, researchers, and funding bodies should enforce adherence to conduct and reporting guidelines before publication. While reporting ranking measures like SUCRA is optional, it aids technology selection, and researchers should always evaluate intervention uncertainties, as differences may be minor and clinically insignificant. |
| Montagner, 2023[37] | Assess and provide an overview of the methodological quality and risk of bias of network meta-analyses in dentistry. | Systematic reviews with network meta-analysis (NMAs) that compared three or more interventions in randomized clinical trials (RCTs), with clinical objectives and measurable outcomes in dental sciences. | 62 | Inception - January 28, 2022 | No | NR | NMA publications have grown significantly since 2010, offering a comprehensive approach to evaluating dental interventions and potentially shifting the systematic review paradigm. However, NMAs should only be conducted with sufficient high-quality evidence and a clear protocol, as studies often show low certainty; otherwise, conducting primary studies is recommended. |
| Nagendrababu, 2023[38] | Evaluate the reporting quality of systematic reviews with network meta-analyses (NMAs) in Endodontics using | Systematic reviews with NMAs in the specialty of endodontics published in English. | 12 | Inception – July, 2021 (full date: NR) | No | PRISMA NMA | None of the NMAs fully complied with the PRISMA-NMA checklist, indicating the need for better reporting in Endodontics, especially in |

| First author, year | Summary of the objectives | Inclusion Criteria | # of NMAs included in the review | Timeframe of assessed literature | Conflicts of interest | Reporting/ methodological guideline used for appraisal (if any) | Synopsis of authors' conclusions |
|---|---|---|---|---|---|---|---|
| | the Preferred Reporting Items for Systematic reviews and Meta-analyses (PRISMA) for NMA checklist. | | | | | | areas like network geometry, bias assessment, and additional analysis. Endorsing PRISMA-NMA guidelines in journals' author instructions can help improve reporting quality and reduce the risk of invalid results. |
| Sehmbi, 2023[39] | Evaluate the methodological and reporting quality of NMAs in anesthesiology. | -NMAs on patients undergoing any form of anesthesia (general, local, or sedation) for any surgical procedure that compared any anesthetic or analgesic interventions with an active or placebo.<br>-NMAs published in full text in English that analyzed RCTs conducted in human patients undergoing any form of anesthesia for any surgical procedure that compared any anesthetic or analgesic interventions with an active or placebo. | 62 | 2013 - October 25, 2020 | Yes | AMSTAR-2, PRISMA, and PRISMA-NMA | Most NMAs within anesthesiology fields were rated "critically low" on the AMSTAR-2 tool, reflecting modest methodological and reporting quality. Dedicated tools and common standards, along with broader adoption of the PRISMA-NMA checklist by journals, are urgently needed to improve quality. |
| Lovsletten, 2024[40] | Map whether and how systematic reviews (SRs) with network meta-analysis (NMA) use presentation formats to report (a) structured evidence summaries and (b) NMA results in general. | -English peer-reviewed SRs with NMA.<br>-NMAs evaluating at least three healthcare interventions in humans.<br>-NMAs with IPD and/or AD, and RCTs and/or observational studies.<br>-The term SR should have been included in title, abstract, or used as keyword/label.<br>-SRs with clear eligibility criteria and searching at least 2 databases. | 200 | January 1, 2020 - December 31, 2021 | Yes | NR | User-friendly, visually compelling presentation formats for NMAs that report structured evidence summaries are underused. Innovations to enhance efficiency of GRADEing processes for network meta-analysis are necessary to greatly improve their usefulness. |

| First author, year | Summary of the objectives | Inclusion Criteria | # of NMAs included in the review | Timeframe of assessed literature | Conflicts of interest | Reporting/ methodological guideline used for appraisal (if any) | Synopsis of authors' conclusions |
|---|---|---|---|---|---|---|---|
| | | -SRs assessing ROB in all primary studies.<br>-NMAs with number of studies larger than the number of nodes in the network. | | | | | |
| Spineli, 2024[41] | Uncover the extent of comparison dissimilarity and commonness of likely intransitivity in several connected networks and demonstrate the disadvantages of using multiple statistical tests to assess the transitivity assumption. | -Networks found in the nmadb database<br>-NMAs extracted from systematic reviews with at least four treatments | 209 | 1999 - April 14, 2015 | No | NICE DSU | There is lack of guidance on dealing with missing data in the context of transitivity assessment. Measuring the overall study dissimilarity between observed comparisons and comparing it with a proper threshold can aid in determining whether concerns of likely intransitivity are warranted. |

**Abbreviations –** AGREE-II: Appraisal of Guidelines, Research and Evaluation – II; AHRQ: Agency for Healthcare Research and Quality; AMSTAR: A Measurement Tool to Assess Systematic Reviews; CADTH: Canada's Drug Agency; CAM: Complementary and Alternative Medicine; DSM: Diagnostic Statistical Manual; DSU: Decision Support Unit; ECA: External Control Arm; EUnetHTA: European network for Health Technology Assessment; GRADE: Grading of Recommendations, Assessment, Development, and Evaluations; HAS: Haute Autorité de Santé; HTA: Health Technology Assessment; IC: Indirect Comparisons; ICD: International Classification of Diseases: IPD: Individual Participant Data; KS: Knowledge Synthesis; MAIC: Matching Adjusted Indirect Comparison; MOD: Missing Outcome Data; MTC: Mixed Treatment Comparisons; ISPOR: International Society for Pharmacoeconomics and Outcomes Research; NICE: National Institute for Health and Care Excellence; NMA: Network Meta-Analysis; NR: No Response; RCT: Randomized Control Trial; RS: Research Studies; PICOS: Population, Intervention, Comparison, Outcomes and Study; PBAC: Pharmaceutical Benefits Advisory Committee; PRISMA: Preferred Reporting Items for Systematic reviews and Meta-Analyses; SLR: Systematic Literature Reviews; SUCRA: Surface Under the Cumulative RAnking curve; SR: Systematic Reviews; STA: Single Technology Appraisal; STC: Simulated Treatment Comparison; TCM: Traditional Chinese Medicine; TSD: Technical Support Document

#### Appendix 10: Table 3: Summary of included studies for the guidance documents (n=23)

**Table 3.** Summary of included studies for the guidance documents (n=23)

| First author, year | Title | Summary of the objectives | Conflicts of interest | Synopsis of authors' conclusions |
|---|---|---|---|---|
| Catala-Lopez, 2014[42] | Network meta-analysis for comparing treatment effects of multiple interventions: an introduction | Provide education to ensure that core methodological considerations underlying network meta-analyses are well understood by readers and researchers to maximize their ability to appropriately interpret findings and appraise validity. | No | Educating readers and researchers on NMA concepts and assumptions is vital for proper interpretation. NMAs should follow rigorous systematic review processes, ensuring transparency, complete reporting, and awareness of limitations like transitivity, consistency, and potential biases in evidence. |
| Cope, 2014[3] | A process for assessing the feasibility of a network meta-analysis: a case study of everolimus in combination with hormonal therapy versus chemotherapy for advanced breast cancer | Outline a general process for assessing the feasibility of performing a valid NMA of RCT. | NR | If a network is not feasible, broadening the evidence base by including additional comparators or integrating non-randomized evidence or individual patient data may help but risks introducing bias from treatment effect modifiers. While these methods are evolving and require further research, the current process offers a foundation for identifying when more complex approaches are needed. |
| Jansen, 2014[43] | Indirect Treatment Comparison/Network Meta-Analysis Study Questionnaire to Assess Relevance and Credibility to Inform Health Care Decision Making: An ISPOR-AMCP-NPC Good Practice Task Force Report | Develop a questionnaire to help evidence evaluators form their opinions on the relevance and credibility of a network meta-analysis to help inform health care decision making. | NR | The Task Force created a consensus-based questionnaire to help decision-makers systematically evaluate the relevance and credibility of NMAs. Feedback will guide updates to improve its utility and support more informed healthcare decisions. |
| Puhan, 2014[44] | A GRADE Working Group approach for rating the quality of treatment effect estimates from network meta-analysis | Present a four-step approach to rate the quality of evidence in each of the direct, indirect, and NMA estimates based on methods developed by the GRADE working group. | No | The GRADE Working Group approach following four steps highlights the necessity for authors of NMA to present direct, indirect, and NMA estimates as well as quality ratings for all direct comparisons. If authors do not present these estimates, scepticism regarding any inferences from the NMA is warranted. |
| CADTH, 2015[45] | Guidance document on reporting indirect comparisons | Provide guidance on reporting indirect comparisons (IDC). | NR | NR |
| Foote, 2015[46] | Network Meta-analysis: Users' Guide for Surgeons: Part I—Credibility | Show the application of evaluation criteria for determining the credibility of a NMA through an example pertinent to clinical orthopaedics. | NR | Assessing the credibility of the methodology is an important first step in critically appraising a NMA. As with conventional systematic reviews, assessing credibility involves evaluating the article for a sensible research question, an exhaustive search, reproducible selection and assessment of articles, |

| First author, year | Title | Summary of the objectives | Conflicts of interest | Synopsis of authors' conclusions |
|---|---|---|---|---|
| | | | | presentation of clinically applicable results, and addressing certainty in effect estimates. |
| Chaimani, 2017[47] | Additional considerations are required when preparing a protocol for a systematic review with multiple interventions | Highlight aspects of a standard systematic review protocol that may need modification when multiple interventions are to be compared. | NR | NMA authors are encouraged to share protocols to reduce post hoc decisions and foster open science. The guidance in this tutorial is broadly applicable and can enhance NMA quality. |
| Chaimani, 2017[48] | Common pitfalls and mistakes in the set-up, analysis and interpretation of results in network meta-analysis: what clinicians should look for in a published article | Propose a practical framework to assess the methodological robustness and reliability of results from network meta-analysis. | No | The validity of NMA results associated with the plausibility of the transitivity assumption, requiring careful consideration of study limitations. While inconsistency can be statistically assessed and should be reported, rankings based on "best probability" can be misleading; clinicians should focus on effect sizes rather than naive rankings. |
| Hummel, 2017[49] | Methodological guidance, recommendations and illustrative case studies for (network) meta-analysis and modelling to predict real-world effectiveness using individual participant and/or aggregate data | Summarise state-of-the-art methods in NMA, IPD meta-analysis and mathematical modelling to predict drug effectiveness based on RCT data and related software, and discuss their advantages and limitations. | NR | Various methods exist to synthesize randomized and non-randomized evidence for regulatory and reimbursement decisions. This report's recommendations, best practices, and tools support researchers in implementing robust evidence synthesis and modeling strategies. |
| Al Khalifah, 2018[50] | Network meta-analysis: users' guide for pediatricians | Discuss how clinicians can evaluate the credibility of NMA methods, and how they can make judgments regarding the quality (certainty) of the evidence. | NR | NMA is a powerful tool but can be misleading due to issues like poor adherence to meta-analysis standards, trial limitations, and NMA-specific challenges like intransitivity and incoherence. This guide helps clinicians evaluate NMA credibility and evidence quality. |
| Brignardello-Petersen, 2018[51] | Advances in the GRADE approach to rate the certainty in estimates from a network meta-analysis | Focus on guidance for systematic reviewers who aim to rate the certainty of the evidence of all the pairwise comparisons from an NMA. | No | As the GRADE approach continues to be used to rate the certainty of estimates from NMA, it is anticipated that further developments will arise from the challenges that will be encountered. All conceptual advances have been and will be discussed in GRADE working group meetings, and only after full discussion will represent GRADE guidance. |
| Dias, 2018[52] | Chapter 12 Validity of Network Meta-Analysis | Introduce and discuss validity of NMAs. | NR | Network meta-analysis simplifies evidence networks by characterizing inputs as weighted averages of contrast estimates, aiding analyses of inconsistency and information flow. These methods could be adapted for sensitivity analyses or guiding data collection to reduce uncertainty, while systematic |

| First author, year | Title | Summary of the objectives | Conflicts of interest | Synopsis of authors' conclusions |
|---|---|---|---|---|
| | | | | reviews should utilize checklists like PRISMA and ISPOR guidance for NMAs. |
| Morton, 2018[53] | Methods Guide for Comparative Effectiveness Reviews: Quantitative Synthesis—An Update | Provide practical recommendations on conducting synthesis. Provide a consistent approach for situations and decisions that are commonly faced by AHRQ Evidence-based Practice Centers (EPCs)" | NR | NR |
| Shi, 2018[54] | Node-making processes in network meta-analysis of non-pharmacological interventions should be well planned and reported | Describe four ways to create a network of nodes based on NMA objectives. | NR | Recommend using additional sources of information to support node-making judgements and that NMAs should report the sources of information. |
| Brignardello-Petersen, 2019[55] | GRADE approach to rate the certainty from a network meta-analysis: avoiding spurious judgments of imprecision in sparse networks | Describe the cause of this phenomenon and provide guidance on how to avoid making spurious assessments of the certainty of the evidence of network estimates in NMAs in which it occurs. | No | Reviewers of sparse networks should address potential issues by planning sensitivity analyses to ensure reliable effect estimates. Neglecting this can lower the certainty of network estimates due to methodological flaws, reducing their value for decision-making. |
| Chaimani, 2019[56] | Undertaking network meta-analyses | Introduce/provide an overview of concepts, assumptions and methods of NMAs. | NR | Much care should be taken when interpreting the results and drawing conclusions from network meta-analysis, especially in the presence of incoherence or other potential biases. |
| Phillippo, 2019[57] | Threshold Analysis as an Alternative to GRADE for Assessing Confidence in Guideline Recommendations Based on Network Meta-Analyses | Argue that GRADE approaches proposed for NMA are insufficient for the purposes of guideline development, as the influence of the evidence on the final recommendation is not accounted for. Outline threshold analysis as an alternative approach. | NR | Threshold analysis is integrated into guideline development by assessing study risk of bias, performing (network) meta-analysis, and analyzing sensitivity to individual comparisons and studies. Identified sensitivities should be explored for plausible evidence changes, enabling decision-makers to assess robustness and incorporate findings into recommendations. |
| Dwan, 2020[58] | Editorial decisions in reviews with network meta-analysis | Present editorial considerations in reviews with NMA. | NR | NR |
| Nikolakopoulou, 2020[59] | CINeMA: An approach for assessing confidence in the results of a network metaanalysis | Present a methodological framework to evaluate confidence in the results from network meta-analyses, Confidence in Network Meta-Analysis (CINeMA), when multiple interventions are compared. | Yes | CINeMA enhances transparency and reduces subjectivity by limiting selective evidence use. Easy to apply even in complex networks, CINeMA provides a rigorous and comprehensive framework for evaluating confidence in treatment effect estimates from NMAs. |
| Welton, 2020[60] | CHTE2020 Sources and Synthesis of Evidence; Update to Evidence Synthesis Methods | Review existing and emerging methods for synthesising evidence on clinical effectiveness for decision- making in Health Technology Appraisals (HTA), including NMA. | NR | NR |

| First author, year | Title | Summary of the objectives | Conflicts of interest | Synopsis of authors' conclusions |
|---|---|---|---|---|
| Chiocchia, 2021[61] | ROB-MEN: a tool to assess risk of bias due to missing evidence in network metaanalysis | Present a tool to assess the Risk Of Bias due to Missing Evidence in Network meta-analysis (ROB-MEN). | Yes | For the comparisons observed for the outcome of interest or other outcomes, the overall judgement considered qualitative assessments for both the within-study and the across study assessment of bias. The assessment of selective outcome reporting bias is likely to be the most valuable because its impact can be quantified more easily than that of publication bias. |
| Lunny, 2023[62] | RoB NMA Explanation and Elaboration: guidance for using a new tool to assess risk of bias (RoB) in network meta-analysis (NMA) | Facilitate and promote the use of the RoB NMA tool. Describe each of the items, which are presented as 'signalling statements'. | Yes | The Risk of Bias in Network Meta-Analysis was developed because no tool existed to assess the risk of bias in this type of evidence synthesis. |
| HTA CG, 2024[63] | Practical Guideline for Quantitative Evidence Synthesis: Direct and Indirect Comparisons | Describe the available methods for direct and indirect comparisons, their underlying assumptions, strengths, and weaknesses, and specify the appropriateness of methods to the data situation. | NR | NR |

**Abbreviations** – AHRQ: Agency for Healthcare Research and Quality; AMCP: Academy of Managed Care Pharmacy; CINeMA: Confidence in Network Meta-Analysis; EPC: Evidence-based Practice Centers; GRADE: Grading of Recommendations, Assessment, Development, and Evaluations; HTA: Health Technology Assessment; IDC: Indirect Comparisons; IPD: Individual Participant Data; ISPOR: International Society for Pharmacoeconomics and Outcomes Research; NCP: National Pharmaceutical Council; NMA: Network Meta-Analysis; NR: No Response; PRISMA: Preferred Reporting Items for Systematic reviews and Meta-Analyses; RCT: Randomized Control Trial; ROB-MEN: Risk Of Bias-Missing Evidence in Network meta-analysis

#### Appendix 11: Full Table 4: Results of additional considerations for PRISMA-NMA and PRISMA 2020 items across all included studies (n=61)

**Table 4.** Results of additional considerations for PRISMA-NMA and PRISMA 2020 items across all included studies (n=61)

| PRISMA 2020 and PRISMA-NMA Item | No. of papers reporting evidence (%) | Additional NMA-related considerations |
|---|---|---|
| (1) Title | 14 (23.0%) | - Report the exact term 'network meta-analysis'[28] (instead of other related terms).<br>- The title should clearly include all elements of the PICOS framework to reflect the scope of the review.[38] |
| (2) Abstract | 15 (24.6%) | - Report the role that interest-holders had in the conduct of the review.[58] |
| **Introduction** | | |
| (3) Rationale | 26 (42.6%) | - Report if the decision to compare interventions is based on clinical judgement or other factors.[3] |
| (4) Objectives | 25 (41.0%) | - Report if an objective was to investigate factors that may modify the treatment effects.[50] |
| **Methods** | | |
| (5) Eligibility Criteria | 33 (54.1%) | - Define the pre-specified intervention nodes, justify the choice of nodes, and describe the method used to establish the nodes.[15, 62] Explain the nodes' inclusion criteria (e.g., expert consensus, previous classification, international standards or conceptual frameworks).[54]<br>- Clarify if all patients included in the trials could be randomized to any of the treatments in the network.[52]<br>- Report the clinically meaningful target population.[52]<br>- Clarify if the status of publication (i.e., grey literature) was used as an inclusion criterion.[19, 20, 22] |
| (6) Information Sources | 26 (32.6%) | N/A |
| (7) Search Strategy | 30 (49.2%) | - Use PRISMA-S guidelines as a template for transparent and reproducible reporting for literature search.[39] |
| (8) Selection Process | 20 (32.8%) | - Report a priori objective criteria for selecting interventions for consideration in the network structure.[39, 62] |
| (9) Data Collection Process | 24 (39.3%) | N/A |
| (10) Data Items | 25 (41.0%) | - List all potential effect modifiers that can impact the transitivity assumption, and any prognostic variables, and how these were identified (e.g. a literature review, expert input, findings from prior subgroup analyses).[30, 58, 63] |
| (S1) Geometry of the Network | 34 (55.7%) | - Report the node-making process – e.g., pre-specified eligibility criteria for primary studies and nodes,[54] node formulation, pre-specified lumping and/or splitting approach, and any assumptions made[15] along with rationale (e.g., based on empirical evidence and/or clinical expertise),[62] and node definition.[54] Report if grouping of treatment classes, treatment doses or treatments in general was performed.[52]<br>- Specify if nodes and/or edges were weighted according to the number and/or size of studies or other characteristics.[64]<br>- Specify whether all compared interventions were connected through RCTs of within-study comparisons or whether steps were taken to connect disconnected interventions using approaches that involved use of observational data, expert opinion, borrowing of data from related conditions or other methods.[62]<br>- Report what the chosen reference intervention is along with rationale (e.g., the intervention most connected to the other interventions in the network).[65] |
| (11) Study Risk of Bias Assessment | 33 (54.1%) | N/A |
| (12) Effect Measures | 25 (41.0%) | - Indicate ways used to examine the uncertainty in the ranking of interventions (e.g. by reporting CIs/CrIs for ranks or all probabilities for each intervention at each possible rank or cumulative ranking curves).[6] |

| | | |
|---|---|---|
| | | - Specify outcome measures: 1. Justify outcome measures selected for analysis. 2. If efficacy-based outcome measures, justify omission of safety/adverse event outcomes in analysis.[45]<br>- Specify if studies share the same outcome measure (defined in the same manner, including same time horizon and reported in the same way).[8]<br>- Specify any thresholds for effects used, review authors must be explicit about the thresholds and should establish them using absolute estimates of effect.[65] Report any decision rules regarding the interpretation of treatment effect estimates and their uncertainty.[3] |
| (13) Synthesis Methods | 48 (78.7%) | - Indicate whether statistical methods were used that preserve within-study randomization (e.g., naïve indirect comparison against adjusted indirect comparison).[11, 18, 21, 31, 49, 66]<br>- In the presence of inconsistency, describe if both direct and indirect evidence were included in NMA[11, 18, 21, 31, 49]<br>- Describe the process that was followed to determine the appropriateness of performing NMAs using the included studies. Indicate if alternative methods to NMA were used (e.g., matching adjusted indirect comparison, simulated treatment comparison).[6, 45, 47, 63]<br>- Describe statistical methods used to model different doses, different co-therapies, treatments, classes, dose or treatment combination models, and trials with multiple outcomes, where appropriate (e.g., dose-effect models, component NMA, multi-outcome models, population adjustment and multilevel network meta-regression models).[13, 52, 63] Describe methods used to compare the different models (e.g., DIC and residual deviance).[13]<br>- Report any sensitivity analyses performed to examine the effects of alternative node-making approaches on findings from NMAs and impact of funding and the associated RoB (e.g., funding bias)[39]on study results and conclusions with rationale.[54]<br>- In subgroup analysis, explain the differences between direct and indirect evidence based upon study characteristics.[53, 63]<br>- Describe the reasons for missing data, the methods and assumptions used for handling missing outcome data (e.g., use of LOCF).[17, 25, 63] |
| (S2) Assessment of Inconsistency (or Incoherence) | 45 (73.8%) | - Describe the analyses performed to assess the hypothesis of consistency and/or clinical and methodological transitivity assumption (e.g., using local and global approaches)[56, 59] and present any graphics generated.[7, 9, 25, 28, 41, 63] Report estimation of heterogeneity and discuss the extent of heterogeneity in each model used.[6, 45, 47]<br>- Describe the RoB introduced by limitations of individual studies and judgement used to infer about the plausibility of intransitivity. Possible effect modifiers could be clinical and methodological.[48, 63] |
| (14) Reporting Bias Assessment | 29 (47.5%) | - Describe any methods used to assess RoB due to missing evidence in NMA (e.g., ROB-MEN).[12, 22, 58, 61, 67] |
| (15) Certainty Assessment | 16 (26.2%) | - Report methods to assess the overall certainty of evidence and the use of any related tools (e.g., CINeMA, GRADE-NMA, GRADE).[12, 17, 21, 30, 39, 51, 59, 61, 64] Report any modifications to reflect specific issues in NMA from standard GRADE, such as contributions of direct evidence to NMA effect estimates, transitivity and consistency.[67] If selected NMA comparisons are to be reported in the Summary of Findings, provide a clear rationale.[58] |
| **Results** | | |
| (16) Study Selection | 27 (44.3%) | - Clarify which studies were identified in the systematic review and which studies were included in each NMA.[9]<br>- Report studies excluded from the analysis due to heterogeneity of populations, study design, etc.[45]<br>- Present evidence that the selection and assessment is reproducible. Duplicate review eligibility and RoB assessment, including a measure of agreement.[46] |
| (S3) Presentation of Network Structure | 32 (52.5%) | N/A |
| (S4) Summary of Network Geometry | 24 (39.3%) | - Report the percentage information that direct evidence contributes to each relative effect estimated in a NMA (e.g., by presenting a contribution matrix).[56]<br>- Report any reasons for observed inconsistencies in NMAs based on the geometry and evidence analysed.[38] Where inconsistency cannot be explained, describe any sensitivity analyses conducted.[60] |

| | | |
|---|---|---|
| (17) Study Characteristics | 27 (44.3%) | - Report the included studies in adequate detail,[24, 25, 32, 35, 36] including information on design and methodology of included studies.[7] |
| (18) Risk of Bias in Studies | 25 (41.0%) | - Provide study contributions with the RoB judgments in a contribution matrix to evaluate within-study bias for each NMA estimate.[59]<br>- Present RoB assessments for each study and pairwise comparison.[61] |
| (19) Results of Individual Studies | 23 (37.7%) | - Presentation of results: When NMAs involve many studies and treatments, present the results of individual studies in online supplementary materials.[28] |
| (20) Results of Synthesis | 40 (64.5%) | - If interventions are classified in different groups, consider reporting the magnitude of effect, certainty of the evidence, and rankings (e.g., rank probabilities, SUCRA values or P-scores), if available, to draw conclusions.[63, 65] If ranking of interventions is provided, consider presenting it along with NMA treatment effects and its uncertainty by outcome.[43, 48]<br>- Present the results from traditional meta-analysis and NMA.[10]<br>- Specify whether results were robust to sensitivity analyses related to NMA assumptions and potential biases.[42]<br>- Report results as per both their clinical and statistical significance.[37]<br>- Report the agreement between confidence and prediction intervals of each treatment effect.[64]<br>- Report and interpret findings alongside available head-to-head trial data.[34] |
| (S5) Exploration of Inconsistency (or Incoherence) | 32 (52.5%) | - In NMAs with at least one close loop, report findings from global and local inconsistency assessments and potential reasons for the observed inconsistencies in the NMAs, if applicable.[10, 33, 38, 45, 63] Clarify when no formal assessment of consistency is possible (e.g., no closed loops in the network).[63]<br>- Report findings of transitivity assessment, and any systematic differences in treatment effect modifiers across treatment comparisons in NMA.[31] |
| (21) Reporting Biases | 24 (39.3%) | - Present the results of the RoB assessment due to missing evidence in NMA (e.g., ROB-MEN).[24, 25, 32, 35-37, 61] |
| (22) Certainty of Evidence | 15 (25.6%) | - Report findings of assessment on certainty of evidence (e.g., in summary of findings tables) for each outcome and comparison separately.[30, 37, 58] Report reasons for rating down certainty in the footnotes of the table that presents the direct, indirect, and network estimates of effect.[44, 50]<br>- Present a table with point estimates, CIs/CrIs, and ratings for all of the direct, network and indirect estimates.[51] |
| **(23) Discussion** | | |
| a) Summary of Evidence | 18 (29.5%) | - Discuss if the content of nodes differs from that in other NMAs assessing the same clinical question.[54]<br>- Discuss the uncertainty of classification of intervention nodes in NMA.[36] |
| b) & c) Limitations | 18 (29.5%) | - Report the impact of RoB and publication bias on findings and conclusions of the review.[19, 20, 22, 37] |
| d) Conclusions | 21 (34.4%) | N/A |
| **Other Information** | | |
| (24) Registration and Protocol | 30 (49.2%) | N/A |
| (25) Support (Funding) | 24 (39.3%) | N/A |
| (26) Competing Interests | 15 (24.6%) | N/A |
| (27) Availability of Data, Code, and Other Materials | 4 (6.6%) | - Describe the statistical NMA model used for combination of results across multiple interventions with associated algebra together with analysis code (including data with a data code sheet explaining the data structure) either in the main text or as an appendix.[5, 32, 35, 36] |

**Abbreviations –** CI: Confidence Interval; CINeMA: Confidence in Network Meta-Analysis; CrI: Credible Interval; DIC: Deviance Information Criterion; GRADE: Grading of Recommendations, Assessment, Development, and Evaluations; LOCF: Last-Observation-Carried-Forward; MID: Minimally Important Difference; N/A: Not Applicable; NMA: Network Meta-Analysis; PICOS: Population, Intervention, Comparison, Outcomes and Study; PRISMA-S: Preferred Reporting Items for Systematic reviews and Meta-Analyses – Statement; RoB: Risk of Bias; ROB-MEN: RoB assessment due to missing evidence in NMA; SUCRA: Surface Under the Cumulative RAnking curve

#### References

1. Hutton B, Salanti G, Chaimani A, Caldwell DM, Schmid C, Thorlund K, et al. The quality of reporting methods and results in network meta-analyses: an overview of reviews and suggestions for improvement. PLoS One. 2014;9(3):e92508. doi: 10.1371/journal.pone.0092508.
2. Hutton B, Catala-Lopez F, Moher D. [The PRISMA statement extension for systematic reviews incorporating network meta-analysis: PRISMA-NMA]. Med Clin (Barc). 2016;147(6):262-6. doi: 10.1016/j.medcli.2016.02.025.
3. Cope S, Zhang J, Saletan S, Smiechowski B, Jansen JP, Schmid P. A process for assessing the feasibility of a network meta-analysis: a case study of everolimus in combination with hormonal therapy versus chemotherapy for advanced breast cancer. BMC Med. 2014;12:93. doi: 10.1186/1741-7015-12-93.
4. Donegan S, Williamson P, D'Alessandro U, Tudur Smith C. Assessing key assumptions of network meta-analysis: a review of methods. Res Synth Methods. 2013;4(4):291-323. doi: 10.1002/jrsm.1085.
5. Tan SH, Bujkiewicz S, Sutton A, Dequen P, Cooper N. Presentational approaches used in the UK for reporting evidence synthesis using indirect and mixed treatment comparisons. J Health Serv Res Policy. 2013;18(4):224-32. doi: 10.1177/1355819613498379.
6. Bafeta A, Trinquart L, Seror R, Ravaud P. Reporting of results from network meta-analyses: methodological systematic review. BMJ. 2014;348:g1741. doi: 10.1136/bmj.g1741.
7. Laws A, Kendall R, Hawkins N. A comparison of national guidelines for network meta-analysis. Value Health. 2014;17(5):642-54. doi: 10.1016/j.jval.2014.06.001.
8. Ortega A, Fraga MD, Alegre-del-Rey EJ, Puigventos-Latorre F, Porta A, Ventayol P, et al. A checklist for critical appraisal of indirect comparisons. Int J Clin Pract. 2014;68(10):1181-9. doi: 10.1111/ijcp.12487.
9. Sullivan SM, Coyle D, Wells G. What guidance are researchers given on how to present network meta-analyses to end-users such as policymakers and clinicians? A systematic review. PLoS One. 2014;9(12):e113277. doi: 10.1371/journal.pone.0113277.
10. Chambers JD, Naci H, Wouters OJ, Pyo J, Gunjal S, Kennedy IR, et al. An assessment of the methodological quality of published network meta-analyses: a systematic review. PLoS One. 2015;10(4):e0121715. doi: 10.1371/journal.pone.0121715.
11. Fleetwood K, Glanville J, McCool R, Wood H, Wilson K, Marshall C, et al. A Review of the Use of Network Meta-Analysis In Nice Single Technology Appraisals. Value in Health. 2016;19(7):348. doi: 10.1016/j.jval.2016.09.009.
12. Ge L, Tian JH, Li XX, Song F, Li L, Zhang J, et al. Epidemiology Characteristics, Methodological Assessment and Reporting of Statistical Analysis of Network Meta-Analyses in the Field of Cancer. Sci Rep. 2016;6:37208. doi: 10.1038/srep37208.
13. Veroniki AA, Straus SE, Soobiah C, Elliott MJ, Tricco AC. A scoping review of indirect comparison methods and applications using individual patient data. BMC Med Res Methodol. 2016;16:47. doi: 10.1186/s12874-016-0146-y.
14. Zarin W, Veroniki AA, Nincic V, Vafaei A, Reynen E, Motiwala SS, et al. Characteristics and knowledge synthesis approach for 456 network meta-analyses: a scoping review. BMC Med. 2017;15(1):3. doi: 10.1186/s12916-016-0764-6.
15. James A, Yavchitz A, Ravaud P, Boutron I. Node-making process in network meta-analysis of nonpharmacological treatment are poorly reported. J Clin Epidemiol. 2018;97:95-102. doi: 10.1016/j.jclinepi.2017.11.018.
16. Lee DW, Shin IS. Critical quality evaluation of network meta-analyses in dental care. J Dent. 2018;75:7-11. doi: 10.1016/j.jdent.2018.05.010.
17. Spineli LM, Yepes-Nunez JJ, Schunemann HJ. A systematic survey shows that reporting and handling of missing outcome data in networks of interventions is poor. BMC Med Res Methodol. 2018;18(1):115. doi: 10.1186/s12874-018-0576-9.

18. Tonin FS, Steimbach LM, Mendes AM, Borba HH, Pontarolo R, Fernandez-Llimos F. Mapping the characteristics of network meta-analyses on drug therapy: A systematic review. PLoS One. 2018;13(4):e0196644. doi: 10.1371/journal.pone.0196644.
19. Tonin FS, Borba HH, Leonart LP, Mendes AM, Steimbach LM, Pontarolo R, et al. Methodological quality assessment of network meta-analysis of drug interventions: implications from a systematic review. Int J Epidemiol. 2019;48(2):620-32. doi: 10.1093/ije/dyy197.
20. Tricco AC, Zarin W, Ghassemi M, Nincic V, Lillie E, Page MJ, et al. Same family, different species: methodological conduct and quality varies according to purpose for five types of knowledge synthesis. J Clin Epidemiol. 2018;96:133-42. doi: 10.1016/j.jclinepi.2017.10.014.
21. Williams T, Stein DJ, Ipser J. A systematic review of network meta-analyses for pharmacological treatment of common mental disorders. Evid Based Ment Health. 2018;21(1):7-11. doi: 10.1136/eb-2017-102718.
22. Yang F, Wang H, Zou J, Li X, Jin X, Cao Y, et al. Assessing the methodological and reporting quality of network meta-analyses in Chinese medicine. Medicine (Baltimore). 2018;97(47):e13052. doi: 10.1097/MD.0000000000013052.
23. Chen Y, Zeng XY, Liu DF, Tan XY, Cai XM, Yang FW, et al. [Critical quality evaluation and application value of network Meta-analyses in traditional Chinese medicine]. Zhongguo Zhong Yao Za Zhi. 2019;44(24):5322-8. doi: 10.19540/j.cnki.cjcmm.20191022.501.
24. Gao Y, Ge L, Ma X, Shen X, Liu M, Tian J. Improvement needed in the network geometry and inconsistency of Cochrane network meta-analyses: a cross-sectional survey. J Clin Epidemiol. 2019;113:214-27. doi: 10.1016/j.jclinepi.2019.05.022.
25. Gao Y, Shi S, Li M, Luo X, Liu M, Yang K, et al. Statistical analyses and quality of individual participant data network meta-analyses were suboptimal: a cross-sectional study. BMC Med. 2020;18(1):120. doi: 10.1186/s12916-020-01591-0.
26. Lee A. Developing critical appraisal of systematic reviews reporting network meta-analysis: University of Oxford; 2020.
27. Pratt M, Wieland S, Ahmadzai N, Butler C, Wolfe D, Pussagoda K, et al. A scoping review of network meta-analyses assessing the efficacy and safety of complementary and alternative medicine interventions. Syst Rev. 2020;9(1):97. doi: 10.1186/s13643-020-01328-3.
28. Bae K, Shin IS. Critical evaluation of reporting quality of network meta-analyses assessing the effectiveness of acupuncture. Complement Ther Clin Pract. 2021;45:101459. doi: 10.1016/j.ctcp.2021.101459.
29. Veroniki AA, Tsokani S, Zevgiti S, Pagkalidou I, Kontouli KM, Ambarcioglu P, et al. Do reporting guidelines have an impact? Empirical assessment of changes in reporting before and after the PRISMA extension statement for network meta-analysis. Syst Rev. 2021;10(1):246. doi: 10.1186/s13643-021-01780-9.
30. Wang R, Dwan K, Showell MG, van Wely M, Mol BW, Askie L, et al. Reporting of Cochrane systematic review protocols with network meta-analyses-A scoping review. Res Synth Methods. 2022;13(2):164-75. doi: 10.1002/jrsm.1531.
31. Wright E, Yasmeen N, Malottki K, Sawyer LM, Borg E, Schwenke C, et al. Assessing the Quality and Coherence of Network Meta-Analyses of Biologics in Plaque Psoriasis: What Does All This Evidence Synthesis Tell Us? Dermatol Ther (Heidelb). 2021;11(1):181-220. doi: 10.1007/s13555-020-00463-y.
32. Yuan T, Xiong J, Wang X, Yang J, Jiang Y, Zhou X, et al. The Quality of Methodological and Reporting in Network Meta-Analysis of Acupuncture and Moxibustion: A Cross-Sectional Survey. Evid Based Complement Alternat Med. 2021;2021:2672173. doi: 10.1155/2021/2672173.
33. Cho SH, Shin IS. Evaluation of the Reporting Standard Guidelines of Network Meta-Analyses in Physical Therapy: A Systematic Review. Healthcare (Basel). 2022;10(12) doi: 10.3390/healthcare10122371.

34. Nast A, Dressler C, Schuster C, Saure D, Augustin M, Reich K. Methods used for indirect comparisons of systemic treatments for psoriasis. A systematic review. Skin Health Dis. 2023;3(1):e112. doi: 10.1002/ski2.112.
35. Wang Y, Chen N, Guo K, Li Y, E F, Yang C, et al. Reporting and methodological quality of acupuncture network meta-analyses could be improved: an evidence mapping. J Clin Epidemiol. 2023;153:1-12. doi: 10.1016/j.jclinepi.2022.11.004.
36. de Sousa PG, Mainka FF, Tonin FS, Pontarolo R. Mapping the characteristics, methodological quality and standards of reporting of network meta-analyses on antithrombotic therapies: An overview. Int J Cardiol. 2023;386:125-33. doi: 10.1016/j.ijcard.2023.05.036.
37. Montagner AF, Angst PDM, Raggio DP, FH VDS, Tedesco TK. Methodological quality of network meta-analysis in dentistry: a meta-research. Braz Oral Res. 2023;37:e062. doi: 10.1590/1807-3107bor-2023.vol37.0062.
38. Nagendrababu V, Narasimhan S, Faggion CM, Jr., Dharmarajan L, Jacob PS, Gopinath VK, et al. Reporting quality of systematic reviews with network meta-analyses in Endodontics. Clin Oral Investig. 2023;27(7):3437-45. doi: 10.1007/s00784-023-04948-w.
39. Sehmbi H, Retter S, Shah UJ, Nguyen D, Martin J, Uppal V. Methodological and reporting quality assessment of network meta-analyses in anesthesiology: a systematic review and meta-epidemiological study. Can J Anaesth. 2023;70(9):1461-73. doi: 10.1007/s12630-023-02510-6.
40. Lovsletten PO, Wang X, Pitre T, Odegaard M, Veroniki AA, Lunny C, et al. A systematic survey of 200 systematic reviews with network meta-analysis (published 2020-2021) reveals that few reviews report structured evidence summaries. J Clin Epidemiol. 2024;173:111445. doi: 10.1016/j.jclinepi.2024.111445.
41. Spineli LM. An empirical study on 209 networks of treatments revealed intransitivity to be common and multiple statistical tests suboptimal to assess transitivity. BMC Med Res Methodol. 2024;24(1):301. doi: 10.1186/s12874-024-02436-7.
42. Catala-Lopez F, Tobias A, Cameron C, Moher D, Hutton B. Network meta-analysis for comparing treatment effects of multiple interventions: an introduction. Rheumatol Int. 2014;34(11):1489-96. doi: 10.1007/s00296-014-2994-2.
43. Jansen JP, Trikalinos T, Cappelleri JC, Daw J, Andes S, Eldessouki R, et al. Indirect treatment comparison/network meta-analysis study questionnaire to assess relevance and credibility to inform health care decision making: an ISPOR-AMCP-NPC Good Practice Task Force report. Value Health. 2014;17(2):157-73. doi: 10.1016/j.jval.2014.01.004.
44. Puhan MA, Schunemann HJ, Murad MH, Li T, Brignardello-Petersen R, Singh JA, et al. A GRADE Working Group approach for rating the quality of treatment effect estimates from network meta-analysis. BMJ. 2014;349:g5630. doi: 10.1136/bmj.g5630.
45. Richter T, Lee KM. Guidance Document on Reporting Indirect Comparisons. In: CADTH, editor. 2015.
46. Foote CJ, Chaudhry H, Bhandari M, Thabane L, Furukawa TA, Petrisor B, et al. Network Meta-analysis: Users' Guide for Surgeons: Part I - Credibility. Clin Orthop Relat Res. 2015;473(7):2166-71. doi: 10.1007/s11999-015-4286-x.
47. Chaimani A, Caldwell DM, Li T, Higgins JPT, Salanti G. Additional considerations are required when preparing a protocol for a systematic review with multiple interventions. J Clin Epidemiol. 2017;83:65-74. doi: 10.1016/j.jclinepi.2016.11.015.
48. Chaimani A, Salanti G, Leucht S, Geddes JR, Cipriani A. Common pitfalls and mistakes in the set-up, analysis and interpretation of results in network meta-analysis: what clinicians should look for in a published article. Evid Based Ment Health. 2017;20(3):88-94. doi: 10.1136/eb-2017-102753.
49. Hummel N, Debray TPA, Didden EM, Efthimiou O, Egger M, Fletcher C, et al. Methodological guidance, recommendations and illustrative case studies for (network) meta-analysis and modelling to predict real-world effectiveness using individual participant and/or aggregate data. 2017 doi: 10.13140/RG.2.2.36349.36327.

50. Al Khalifah R, Florez ID, Guyatt G, Thabane L. Network meta-analysis: users' guide for pediatricians. BMC Pediatr. 2018;18(1):180. doi: 10.1186/s12887-018-1132-9.
51. Brignardello-Petersen R, Bonner A, Alexander PE, Siemieniuk RA, Furukawa TA, Rochwerg B, et al. Advances in the GRADE approach to rate the certainty in estimates from a network meta-analysis. J Clin Epidemiol. 2018;93:36-44. doi: 10.1016/j.jclinepi.2017.10.005.
52. Dias S, Ades AE, Welton NJ, Jansen JP, Sutton AJ. Network Meta-Analysis for Decision Making: Wiley; 2018.
53. Morton SC, Murad MH, O'Connor E, Lee CS, Booth M, Vandermeer BW, et al. Quantitative Synthesis-An Update. Methods Guide for Effectiveness and Comparative Effectiveness Reviews. AHRQ Methods for Effective Health Care. Rockville (MD)2018.
54. Shi C, Westby M, Norman G, Dumville JC, Cullum N. Node-making processes in network meta-analysis of nonpharmacological interventions should be well planned and reported. J Clin Epidemiol. 2018;101:124-5. doi: 10.1016/j.jclinepi.2018.04.009.
55. Brignardello-Petersen R, Murad MH, Walter SD, McLeod S, Carrasco-Labra A, Rochwerg B, et al. GRADE approach to rate the certainty from a network meta-analysis: avoiding spurious judgments of imprecision in sparse networks. J Clin Epidemiol. 2019;105:60-7. doi: 10.1016/j.jclinepi.2018.08.022.
56. Chaimani A, Caldwell DM, Li T, Higgins JPT, Salanti G. Undertaking network meta-analyses. Cochrane Handbook for Systematic Reviews of Interventions. 2nd ed2019.
57. Phillippo DM, Dias S, Welton NJ, Caldwell DM, Taske N, Ades AE. Threshold Analysis as an Alternative to GRADE for Assessing Confidence in Guideline Recommendations Based on Network Meta-analyses. Ann Intern Med. 2019;170(8):538-46. doi: 10.7326/M18-3542.
58. Dwan K, Livingstone N. Editorial considerations in reviews with network meta-analysis 2020. Available from: [https://training.cochrane.org/resource/editorial-considerations-reviews-network-meta-analysis].
59. Nikolakopoulou A, Higgins JPT, Papakonstantinou T, Chaimani A, Del Giovane C, Egger M, et al. CINeMA: An approach for assessing confidence in the results of a network meta-analysis. PLoS Med. 2020;17(4):e1003082. doi: 10.1371/journal.pmed.1003082.
60. Welton NJ, Phillippo DM, Owen R, Jones HE, Dias S, Bujkiewicz S, et al. CHTE2020 SOURCES AND SYNTHESIS OF EVIDENCE; UPDATE TO EVIDENCE SYNTHESIS METHODS. 2020.
61. Chiocchia V, Nikolakopoulou A, Higgins JPT, Page MJ, Papakonstantinou T, Cipriani A, et al. ROB-MEN: a tool to assess risk of bias due to missing evidence in network meta-analysis. BMC Med. 2021;19(1):304. doi: 10.1186/s12916-021-02166-3.
62. Lunny C, Veroniki AA, Higgins JPT, Dias S, Hutton B, Wright JM, et al. Methodological review of NMA bias concepts provides groundwork for the development of a list of concepts for potential inclusion in a new risk of bias tool for network meta-analysis (RoB NMA Tool). Syst Rev. 2024;13(1):25. doi: 10.1186/s13643-023-02388-x.
63. Practical Guideline for Quantitative Evidence Synthesis: Direct and Indirect Comparisons. In: Commission E, editor. 2024.
64. Papakonstantinou T, Nikolakopoulou A, Higgins JPT, Egger M, Salanti G. CINeMA: Software for semiautomated assessment of the confidence in the results of network meta-analysis. Campbell Syst Rev. 2020;16(1):e1080. doi: 10.1002/cl2.1080.
65. Brignardello-Petersen R, Izcovich A, Rochwerg B, Florez ID, Hazlewood G, Alhazanni W, et al. GRADE approach to drawing conclusions from a network meta-analysis using a partially contextualised framework. BMJ. 2020;371:m3907. doi: 10.1136/bmj.m3907.
66. Lunny C, Veroniki AA, Hutton B, White I, Higgins J, Wright JM, et al. Knowledge user survey and Delphi process to inform development of a new risk of bias tool to assess systematic reviews with network meta-analysis (RoB NMA tool). BMJ Evid Based Med. 2022 doi: 10.1136/bmjebm-2022-111944.

67. Salanti G, Del Giovane C, Chaimani A, Caldwell DM, Higgins JP. Evaluating the quality of evidence from a network meta-analysis. PLoS One. 2014;9(7):e99682. doi: 10.1371/journal.pone.0099682.
